# Blood cell differential count discretization methods to predict survival in acutely ill adults reporting to the emergency room: a retrospective cohort study in 2020

**DOI:** 10.1101/2023.01.13.22283538

**Authors:** Riccardo M. Fumagalli, Marco Chiarelli, Mauro P. Zago, Massimo Cazzaniga, Luciano D’angelo, Mario Cerino, Sabina Terragni, Elisa Lainu, Cristina Lorini, Claudio Scarazzati, Sara Tazzari, Francesca Porro, Simone Aldé, Morena Burati, William Brambilla, Daria Valsecchi, Paolo Spreafico, Valter Tantardini, Gianpaolo Schiavo, Claudio Bonato, Luca Cavalieri d’oro, Stefano Nattino, Matteo Locatelli, Luca A. M. Fumagalli

**Affiliations:** Dip.Emergenza Accettazione (DEA), Pronto Soccorso, Ospedale “A.Manzoni”, Lecco, Italy; Klinik für Angiologie, Universitätsspital Zürich, Zurich, Switzerland; Dip.Chirurgico, Chirurgia Urgenza, Ospedale “A.Manzoni”, Lecco, Italy; DEA, Pronto Soccorso, Ospedale “A.Manzoni”, Lecco, Italy; Dip. Servizi Clinici, Ospedale “A.Manzoni”, Lecco, Italy; UOC Epidemiologia, ATS Brianza, Monza, Italy; Università degli studi di Milano, scuola di specializzazione in Medicina d’Emergenza-Urgenza, Milano, Italy; Polo formativo ATS Brianza, Monza, Italy; Ospedale “A.Manzoni”, Lecco, Italy

**Keywords:** Hospital emergency services, Survival, Mortality, Complete blood counts

## Abstract

**Aims:** To assess survival predictivity of baseline blood cell differential count (BCDC) discretization methods in acutely ill adults visiting the emergency room over one-year.

**Methods:** Retrospective cohort study on one-year survival of adults reporting to the emergency room of the A. Manzoni Hospital (Italy) during 2020. Automated BCDC analysis performed at baseline, assessed hemoglobin, red cell mean volume and distribution width (RDW), platelet distribution width (PDW), platelet-hematocrit, absolute red blood cells, white blood cells, neutrophils, lymphocytes, monocytes, eosinophils, basophils, and platelets. Discretization cutoffs were defined by: Benchmark laboratory reference values and Tailored (maximally selected rank statistics for linear and sigmoid-shaped distributed variables; optimal-equal hazard ratio (HR) method for U-shaped distributed variables. Explanatory variables (age, gender, inward admission) were analyzed using Cox multivariable regression. Receiver operating characteristic curves used the sum of Cox-significant variables in each method.

**Results:** Of 11052 patients (median age 67 years, interquartile range (IQR) 51–81, 48% female), 59% (n=6489) were discharged and 41% (n=4563) were admitted. After a 306-day median follow up (IQR 208–417 days), 9455 (86%) patients were alive and 1597 (14%) deceased. Increased HRs were associated with age > 73 years (HR=4.29 CI 3.78–4.87), and hospital admission (HR=2.05, CI 1.83–2.29). Age, sex, hemoglobin, mean corpuscular volume, RDW, PDW, neutrophils, lymphocytes and eosinophils were significant in overall. Benchmark included basophils and platelet count (area under the ROC curve (AUROC) 0.78). Tailored included monocyte counts and PCT (AUROC of 0.82).

**Conclusions:** Tailored discretization of BCDC provided meaningful insight regarding acute patient survival.

**Key messages:** *What is already known on this topic:* Information on survival predictivity of BCDC is scarce, particularly in acutely ill patients considering that reference values are based on the general population.

*What this study adds:* Laboratory reference interval values predicting survival were hemoglobin, RDW, MCV, neutrophil, lymphocyte, eosinophil, and basophils counts, PLT, and PDW, independently of sex, age, and acute inward admission. Survival predictivity was improved by discretization of hemoglobin, RDW, MCV, neutrophil, lymphocyte, eosinophil, and monocyte counts, and PDW, according to the maximally selected rank statistics and optimal-equal HR method.

*How this study might affect research, practice, or policy:* Baseline BCDC discretized by tailored methods may be a useful biomarker for hazard warning in acute illness.

## Introduction

Immune response to chronic and acute diseases may drive disease outcomes, but assessments are rarely standardized in clinical practice. Therefore, it is often restricted to measuring total white cell count in peripheral blood, a low-cost easily performed laboratory test. Few studies have reported the predictivity of blood cell count on patient survival under acute and non-acute conditions.[1,2,3]

Reference values of blood cell differential counts are calculated based on the general population, whereas less details are known for acutely ill patients. In this population subset, the identification of differential blood cell cutoff values may be challenging considering nonlinear effects of continuous variables on survival.[4,5] Among non-linear relationships, U-shaped associations between continuous biological variables and outcomes are commonly observed in clinical and epidemiological studies, and several statistical methods have been tested to solve the issue.[6] The severe acute respiratory syndrome-coronavirus 2 (SARS-CoV2) epidemic affected emergency healthcare facilities worldwide and new variants are expected to spread, although with varying morbidity and mortality. Therefore, including SARS-CoV2 patients may aid the identification of survival biomarkers for acute patients[7].

To assess predictivity of peripheral blood cell differential count on survival, we retrospectively discretized automated peripheral blood cell differential counts. These were performed at the initial visit of acutely ill adult or trauma patients to our Emergency Room (ER) during 2020. Patients were assessed using laboratory reference benchmark values (Benchmark method) and selected optimal cutoff finding methods (Tailored methods), namely optimal equal hazard ratio (OEHR)[6] and maximally selected rank statistic method (MSRS)[8]. We constructed two models selecting variables among differential counts independently associated with long term overall survival. Finally, we analyzed and compared the area under the receiver operating characteristic (ROC) curve (AUROC) of both models originated from the sum of variables obtained by both methods.

## Methods

This retrospective cohort analysis was conducted using data recorded at the Emergency Room of the A. Manzoni Hospital, Lecco, Italy (600 beds, national health services (NHS) hospital) between January 1^st^ and December 31^st^, 2020. Eligible patients were aged > 18 years, consecutively admitted to the Emergency Room (ER) for acute illness or trauma, and for whom a complete blood differential count collection was indicated by ER medical staff at first presentation. The recorded patient characteristics were age, sex, and outcome after evaluation by the ER medical staff, including discharge or admission to any hospital ward. Complete blood cell differential count (BCDC) was performed using the automated Sysmex XN-9000 analyzer on peripheral blood samples taken at baseline.

Survival was the referral outcome for explorative model development and was assessed on June 30th, 2021, by a population registry office query through the NHS territorial service. Predictors were searched among the BCDC automated analysis assessment of hemoglobin (Hb), mean red cell volume (MCV), red cell distribution width (RDW), platelet distribution width (PDW), platelet hematocrit (PCT) and absolute count of red blood cells (RBC), white blood cells (WBC), neutrophils (Ne), lymphocytes (Ly), monocytes (Mo), eosinophils (Eo), basophils (Ba), and platelets (PLT). Missing data were excluded, as only patients having BCDC records were evaluated.

Statistical analysis was carried out using descriptive methods for distribution and dispersion (skewness and kurtosis). Explorative analyses of continuous variables and differences between live and dead patients was performed by Independent Samples T-Test (nonparametric) Mann-Whitney U test.

The “Benchmark” reference model was set by discretization of BCDC values on our laboratory reference interval, established according to the C28-A3 guideline by the Clinical and Laboratory Standards Institute (CLSI).[9]

The “Tailored” discretization was set as follows. The relationship between each continuous variable and log relative hazard were plotted using the penalized B-splines (psplines) technique[10] for fitting the nonlinear effect of covariate in Cox models[11] by minimizing pitfalls associated with dichotomization of biological variables.[4]

Variables were treated differently according to their respective distribution profile.

Linear and sigmoid-shaped variables were dichotomized by the maximally selected rank statistic method (MSRS).[8] U-shaped variables were univariately discretized by cutoff point determination using the optimal-equal hazard ratio method (OEHR).[6]

Discretized explanatory variables were defined as favorable and unfavorable. There variables were then analyzed using a Cox multivariable regression to build two models (one for each discretization method) containing independently significant BCDC subsets and demographic characteristics among the study participants. For each of the two discretization methods, we counted how many factors presented value classified as unfavourable, to calculate a score sum.

Actuarial curves of the whole population, stratified into five risk groups according to quintiles were calculated using the Kaplan–Meier method and compared by log–rank test in either method. Finally, a receiver operating curve (ROC curve) for both discretization methods was drawn using the sum of factors classified as unfavourable. ROC curves were then compared using the DeLong test.[12]

Ethical obligations were fulfilled by the Hospital Board, in compliance with national regulations regarding retrospective observational studies.

Analyses were performed using R and Jamovi (R-based free software).[13,14]

## Results

In this study, 11635 complete blood cell counts were registered in the Emergency General Department during 2020. Patients younger than 18 years and pregnant women in labor were excluded as listed in the Pediatric and Obstetric Emergency Laboratory Services subsets. After further removal of duplicated tests and repeated admissions, 11133 patients remained. Outliers exceeding 99% of BCDC element values were removed (81 patients) to exclude extreme outliers, most likely affected by hematologic diseases. Finally, 11052 patients were available for analysis. Median age was 67 years (IQR 51–81) and 48% of participants were female (Fig,1).

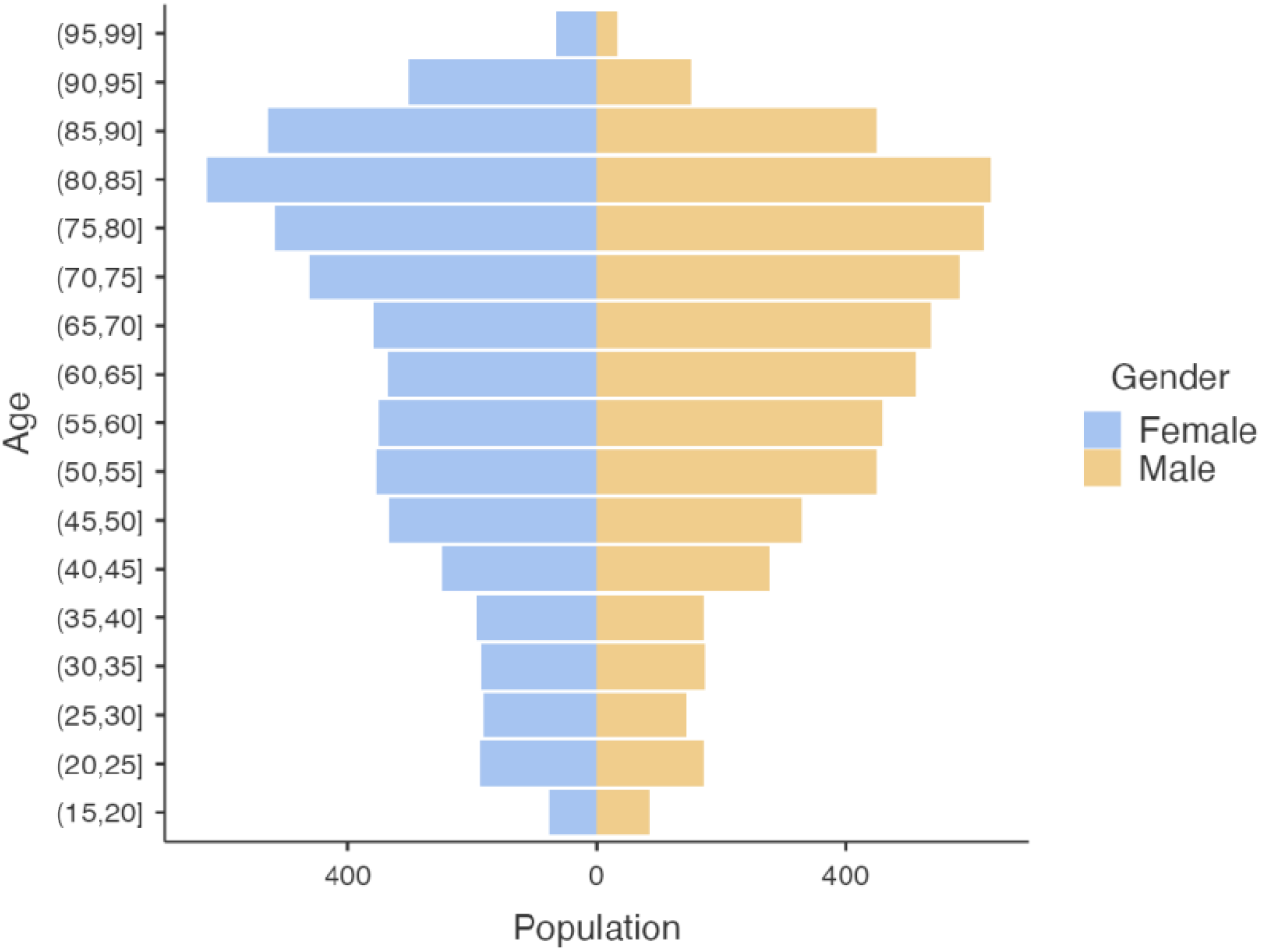

Fifty-nine percent of patients (n= 6489) were discharged and 41% (n=4563) were acutely admitted to hospital wards.

After a median follow up of 306 days (IQR 208–417 dd), 9455 patients (86%) were alive and 1597 (14%) were deceased (Tab.1).

**Table 1.** Patient Characteristics.

| Patients Characteristics |  | N = 11,052 |
| --- | --- | --- |
| Sex n (%) |  |  |
| Female |  | 5,284 (48%) |
| Male |  | 5,768 (52%) |

| **Patients Characteristics** | **N = 11,052** |
| --- | --- |
| Age median years (IQR) | 67 (51, 81) |
| Discharged/ Inward n (%) |  |
| Discharged at home | 6,489 (59%) |
| Inward Admitted | 4,563 (41%) |
| Alive/deceased n (%) |  |
| Alive | 9,455 (86%) |
| Dead | 1,597 (14%) |

<sup>1</sup> Statistics presented: n (%); median, interquartile range (IQR).

Out of 4563 patients acutely admitted to the hospital, 543 (12%) were deceased within 30 days of admission, 599 (13%) were deceased more than 30 days of admission whereas 3421 (75%) were alive at a median time of 10 months.

Out of 6489 patients Discharged from the Emergency Department, 204 (3%) were deceased within 30 days, 251 (4%) died later, whereas 6034 (93%) were alive after a median time of 10 months (Tab.2).

**Table 2.** Discharged and inward admitted patients

|  | **Discharged (N=6489)** | **Inward (N=4563)** | **Total (N=11052)** | **p value** |
| --- | --- | --- | --- | --- |
|  |  |  |  | < 0.001<sup>1</sup> |
| alive | 6034 (93.0%) | 3421 (75.0%) | 9455 (85.6%) |  |
| deceased within 30 days | 204 (3.1%) | 543 (11.9%) | 747 (6.8%) |  |
| deceased after 30 days | 251 (3.9%) | 599 (13.1%) | 850 (7.7%) |  |

<sup>1</sup>Pearson's Chi-squared test

The flow of discharged and inward-admitted patients on deceased or alive status is depicted in Fig.2.

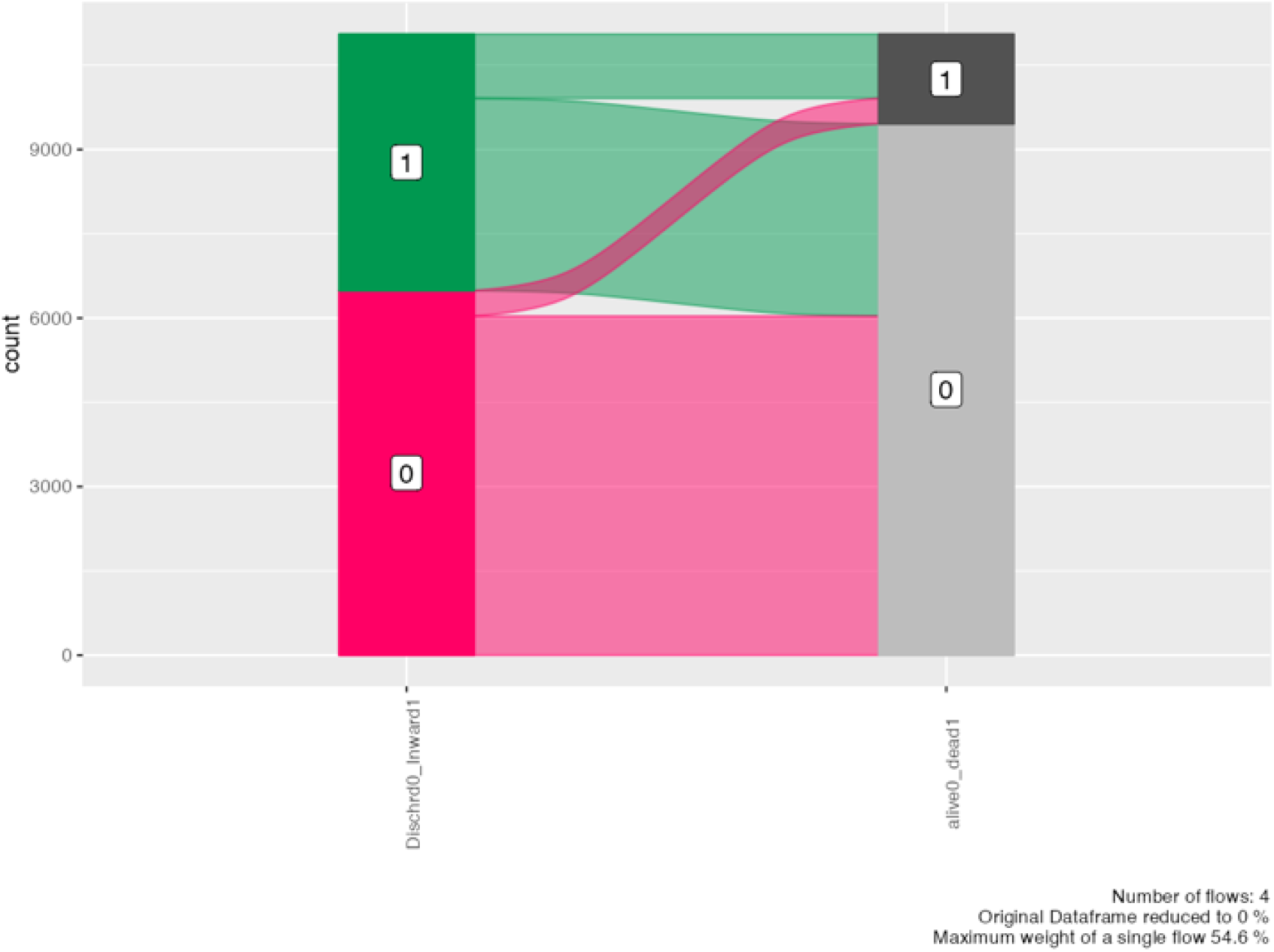

Descriptive statistics, skewness, and kurtosis of the BCDC variables are reported in supplemental material (Tab.S/1). Most of the BCDC subsets were nonparametric. Variable density plots by alive and deceased groups are depicted in supplemental material (FigS/1).

Differences between alive and deceased groups were explored using the continuous variables analysis and compared using the Mann-Whitney U test. As described in supplemental material (Tabl.S/2 Fig.S/2), all variables, except for monocyte count, significantly differed between dead and alive groups. Overall survival is plotted in Fig.3.

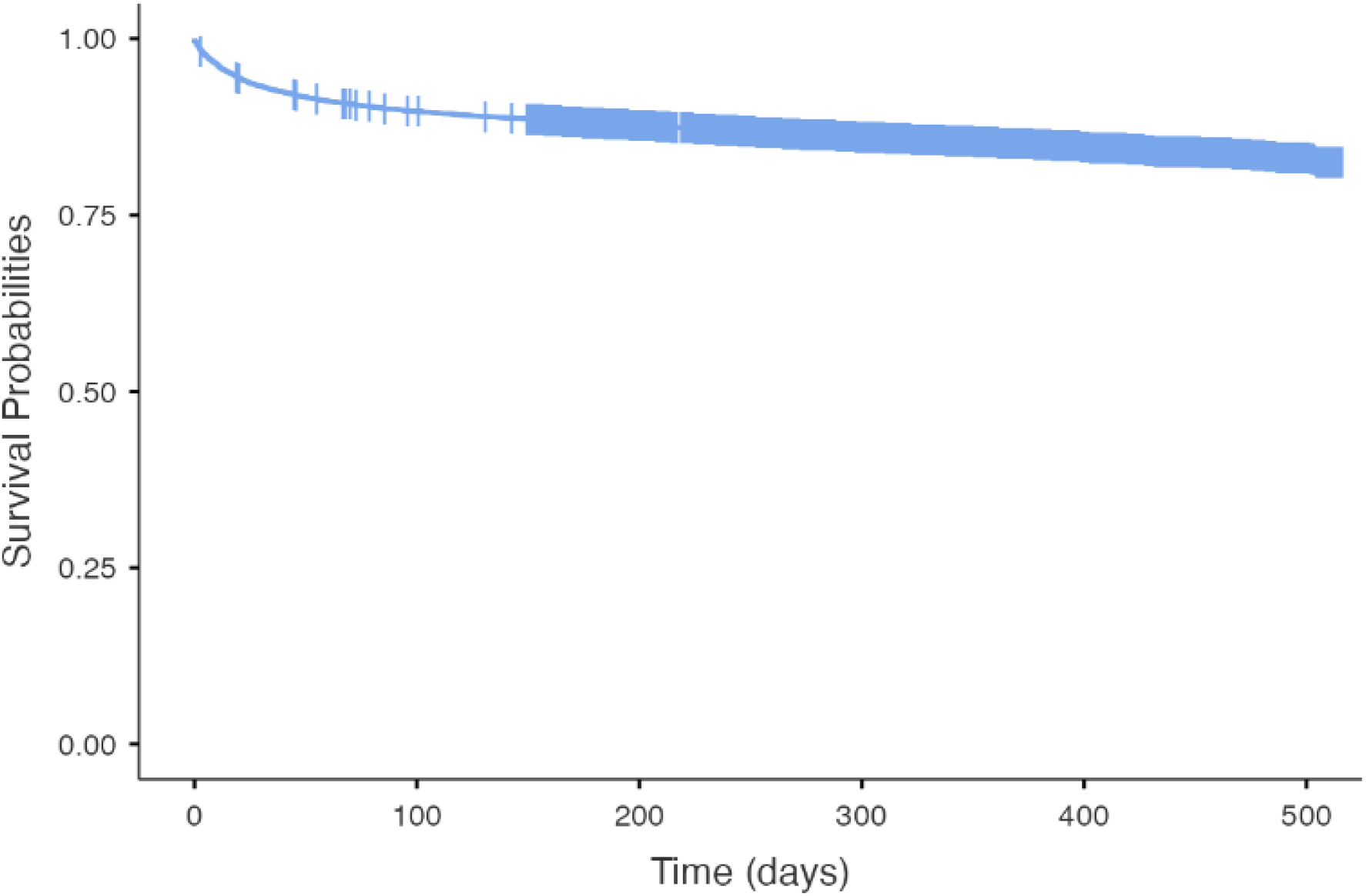

Relationship between each continuous variable and log relative hazard were plotted using the penalized B splines technique[10] (Fig 4).

**Fig 4.**
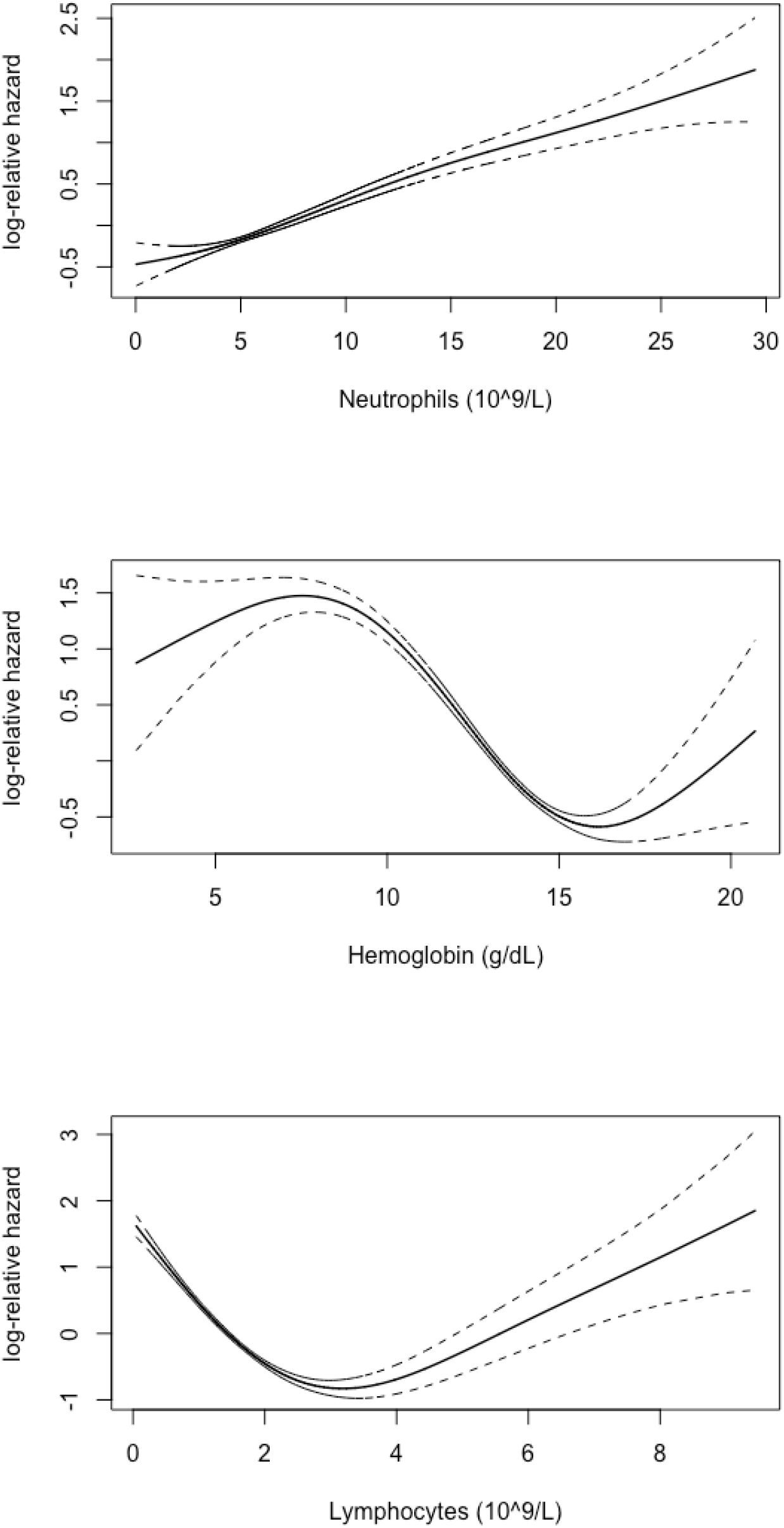
shape exemplary of covariates Penalized-B splines (psplines) top =: linear; middle=sigmoid; bottom= asymmetric U

Age, Ne count and Hb, MCV, RDW shown linear and sigmoid shape, respectively, so were dichotomized by the maximally selected rank statistic method (MSRS).[8] WBC, Ly, Mo, Eo, Ba count, PLT, PCT, and PDW shown nonlinear U-shape and were univariately discretized by optimal-equal hazard ratio method (OEHR).[6]

Results of variables discretization into favorable and unfavorable value intervals, according to selected methods are detailed in Table 4.

**Table 4.** BCDC Laboratory Reference intervals (Benchmark) and unfavourable intervals by MSRS/OEHR (Tailored) discretization.

|  |  | Benchmark<br>discretization | Tailored<br>discretization |  |
| --- | --- | --- | --- | --- |
| Covariate | Distribution<br>profile | Laboratory reference<br>interval (CLSI) | univariate MSRS<br>Unfavorable<br>interval | univariate OEHR<br>Unfavorable<br>interval |
|  |  | Lower -upper | Single Cutoff | L-R Cutoff |
| Age (years) | Linear |  | >73 |  |
| Hemoglobin (g/dL) | Linear | 13.5- 17 | <11.7 |  |
| RBC (10<sup>9</sup>/L) | Sigmoid | 4.50- 5.90 | <3.79 |  |
| MCV (fl) | Sigmoid | 80- 96 | >97 |  |
| RDW (%) | Sigmoid | 12.2- 15 | >13.8 |  |
| WBC (10<sup>9</sup>/L) | Asym-U | 4.0- 9.0 |  | <0.36 >15.43 |
| Neutrophils (10<sup>9</sup>/L) | Linear | 1.60- 6.80 | >9.65 |  |
| Lymphocytes (10<sup>9</sup>/L) | Asym-U | 0.8- 4.5 |  | <1.04 >6.42 |
| Monocytes (10<sup>9</sup>/L) | Asym-U | 0.12- 1.08 |  | <0.25 >1.32 |
| Eosinophils (10<sup>9</sup>/L) | Asym-U | 0.05- 0.65 |  | <0.01 >1.41 |
| Basophils (10<sup>9</sup>/L) | Asym-U | 0.01- 0.20 |  | <0.01 >0.12 |
| PLT (10<sup>9</sup>/L) | Asym-U | 150- 400 |  | <131 >426 |
| PCT (%) | Asym-U | 0.19- 0.40 |  | <0.14 >0.39 |
| PDW (%) | Symm-U | 9.6- 15.2 |  | <8.9 >16 |

Abbreviations: Hemoglobin (Hb), mean red cell volume (MCV), red cell distribution width (RDW), platelet distribution width (PDW), platelet hematocrit (PCT) and absolute count of red blood cells (RBC), white blood cells (WBC), platelets (PLT), maximally selected rank statistic (MSRS), optimal-equal hazard ratio (OEHR), asymmetrical (Asym), symmetrical (Symm).

Details and graphics on variable discretization for both OEHR and MSRS techniques are reported in supplemental material (Section OEHR MSRS Techniques).

Concerning the Benchmark discretization method, the Cox multivariable survival analysis regression of variables was discretized into on-reference and off-reference intervals, according to the reference laboratory value (supplemental material Table S/3,S/4 Fig S/3, S/4).

Sex, age and emergency room medical staff indication for discharge or inward admission were included in the model. As expected, the strongest predictors were age >73 years HR= 4.49(CI 3.96– 5.09) and inward admission, HR= 2.17 (CI 1.94–2.43), whereas male sex HR was = 1.29(1.16–1.43). Benchmark BCDC independently significant unfavorable values are detailed in Tab.5 and plotted in Fig.5.

**Table 5.** Hazards Regression -Benchmark BCDC, age, gender, inward/discharged

| **Dependent: Surv** | **Count (%)** | **HR (univariable)** | **HR (multivariable)** |
| --- | --- | --- | --- |
| sex_OF_1M Female 0 | 5284 (100.0) | - | - |
| Male 1 | 5768 (100.0) | 1.16 (1.05-1.28, p=0.003) | 1.29 (1.16-1.43, p<0.001) |
| Age_0fav (<73 yrs) 0 | 6693 (100.0) | - | - |
| 1unfavour (>73 yrs) 1 | 4359 (100.0) | 6.72 (5.95-7.58, p<0.001) | 4.49 (3.96-5.09, p<0.001) |
| Discharged 0 | 6489 (100.0) | - | - |
| Inward 1 | 4563 (100.0) | 4.03 (3.61-4.49, p<0.001) | 2.17 (1.94-2.43, p<0.001) |
| HB_ref (13.5-17) 0 | 5806 (100.0) | - | - |
| Off ref 1 | 5246 (100.0) | 2.79 (2.51-3.11, p<0.001) | 1.56 (1.39-1.74, p<0.001) |
| MCV_ref (80-96) 0 | 8872 (100.0) | - | - |
| Off ref 1 | 2180 (100.0) | 2.26 (2.03-2.50, p<0.001) | 1.26 (1.13-1.40, p<0.001) |
| RDW_ref (12.2-15) 0 | 8015 (100.0) | - | - |
| Off ref 1 | 3037 (100.0) | 1.99 (1.80-2.20, p<0.001) | 1.57 (1.42-1.74, p<0.001) |
| Ne_ref (1.60-6.80) 0 | 6698 (100.0) | - | - |
| Off ref 1 | 4354 (100.0) | 1.83 (1.66-2.01, p<0.001) | 1.36 (1.23-1.51, p<0.001) |
| Ly_ref (0.8-4.5) 0 | 9197 (100.0) | - | - |
| Off ref 1 | 1855 (100.0) | 3.03 (2.73-3.35, p<0.001) | 1.45 (1.30-1.63, p<0.001) |
| Eo_ref (0.05-0.65) 0 | 6384 (100.0) | - | - |
| Off ref 1 | 4668 (100.0) | 2.10 (1.90-2.32, p<0.001) | 1.34 (1.20-1.49, p<0.001) |
| Ba_ref (0.01-0.20) 0 | 10497 (100.0) | - | - |
| Off ref 1 | 555 (100.0) | 2.85 (2.44-3.32, p<0.001) | 1.44 (1.22-1.70, p<0.001) |
| PLT_ref (150-400) 0 | 9423 (100.0) | - | - |
| Off ref 1 | 1629 (100.0) | 2.42 (2.17-2.70, p<0.001) | 1.33 (1.18-1.49, p<0.001) |
| PDW_ref (9.6-15.2) 0 | 9167 (100.0) | - | - |
| Off ref 1 | 1885 (100.0) | 1.58 (1.41-1.78, p<0.001) | 1.17 (1.04-1.32, p=0.009) |

**Fig 5.**
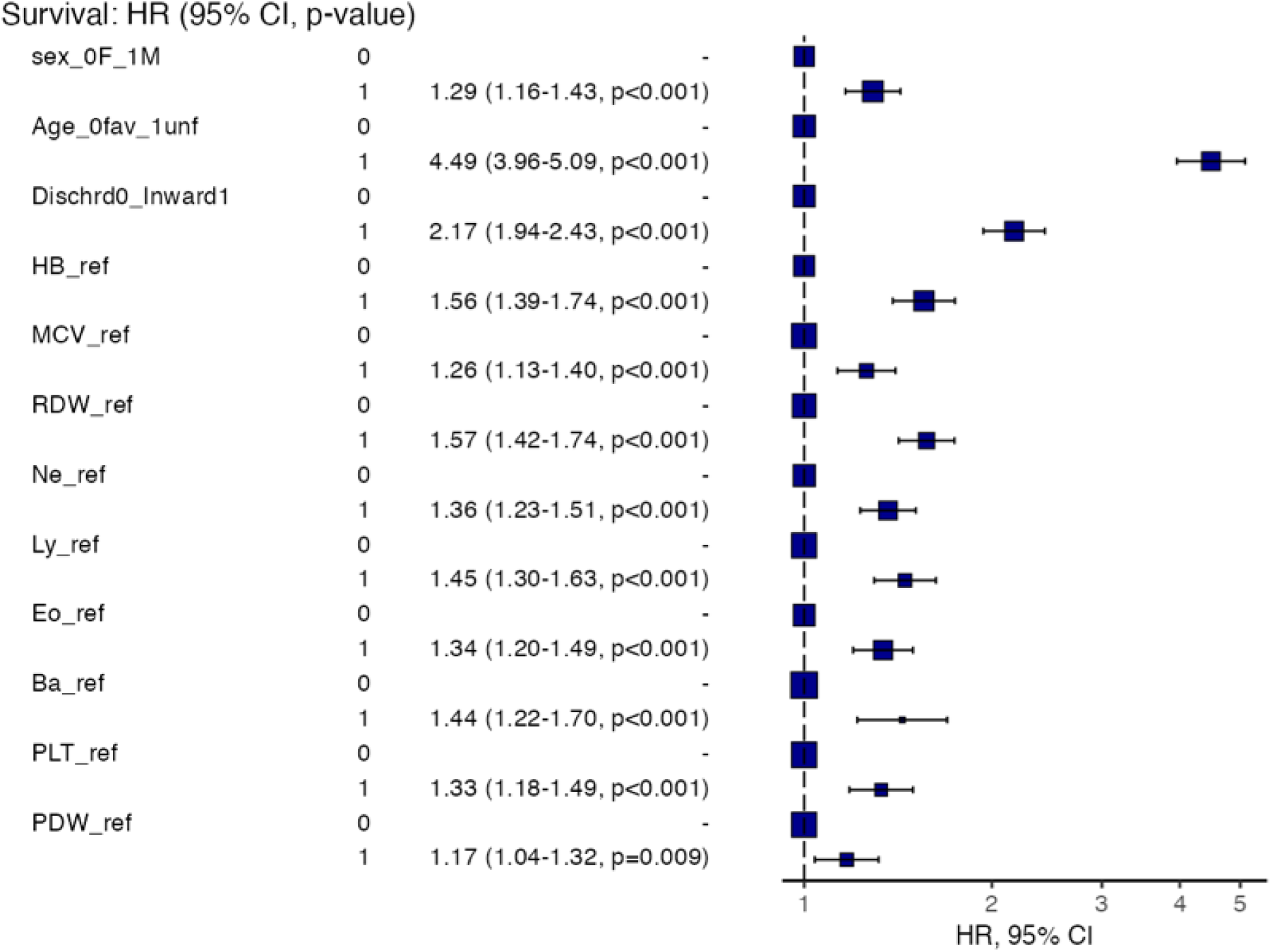
Hazards Regression Plot -Benchmark BCDC

Regarding the Tailored discretization method, by Cox multivariable survival analysis regression strong predictors were age >73 years HR= 3.77(CI 3.32–4.28) and inward admission HR= 1.93(CI 1.72–2.17). Male sex HR was 1.26(CI 1.14–1.39). Tailored BCDC independently significant unfavorable values are detailed in Tab.6 and depicted in Fig.6

**Table 6.** Hazards Regression -Tailored BCDC, age, gender, inward/discharge

| **Dependent: Surv** |  | **Count (%)** | **HR (univariable)** | **HR (multivariable)** |
| --- | --- | --- | --- | --- |
| sex_OF_1M. Female | 0 | 5284 (100.0) | - | - |
| Male | 1 | 5768 (100.0) | 1.16 (1.05-1.28, p=0.003) | 1.26 (1.14-1.39, p<0.001) |
| Age_0fav (<73 yrs) | 0 | 6693 (100.0) | - | - |
| 1unfavour (>73) | 1 | 4359 (100.0) | 6.72 (5.95-7.58, p<0.001) | 3.77 (3.32-4.28, p<0.001) |
| Discharged | 0 | 6489 (100.0) | - | - |
| Inward | 1 | 4563 (100.0) | 4.03 (3.61-4.49, p<0.001) | 1.93 (1.72-2.17, p<0.001) |
| Hb_DIC (>11.7) fav | 0 | 9297 (100.0) | - | - |
| (<11.7) unf | 1 | 1755 (100.0) | 3.59 (3.24-3.97, p<0.001) | 1.42 (1.27-1.58, p<0.001) |
| MCV_DIC (<97) fav | 0 | 9737 (100.0) | - | - |
| (>97) unf | 1 | 1315 (100.0) | 2.76 (2.46-3.10, p<0.001) | 1.41 (1.25-1.58, p<0.001) |
| RDW_DIC (<13.8) fav | 0 | 7360 (100.0) | - | - |
| (>13.8) unf | 1 | 3692 (100.0) | 4.76 (4.29-5.29, p<0.001) | 2.22 (1.98-2.49, p<0.001) |
| NeDIC (<9.65) fav | 0 | 9118 (100.0) | - | - |
| (>9.65) unf | 1 | 1934 (100.0) | 2.11 (1.89-2.35, p<0.001) | 1.35 (1.21-1.52, p<0.001) |
| Ly0fav (>1.04<6.42) | 0 | 8068 (100.0) | - | - |
| (<1.04>6.42) unf | 1 | 2984 (100.0) | 3.04 (2.75-3.35, p<0.001) | 1.37 (1.23-1.52, p<0.001) |
| Mo_0fav (>0.25<1.32) | 0 | 10167 (100.0) | - | - |
| (<0.25>1.32) unf | 1 | 885 (100.0) | 2.36 (2.06-2.71, p<0.001) | 1.21 (1.05-1.39, p=0.008) |
| Eo0fav (>0.01<1.41) | 0 | 9304 (100.0) | - | - |
| (<0.01>1.41) unf | 1 | 1748 (100.0) | 3.06 (2.76-3.40, p<0.001) | 1.69 (1.50-1.90, p<0.001) |
| PCT_0fav (>0.14<0.39) | 0 | 9839 (100.0) | - | - |
| (<0.14>0.39) unf | 1 | 1213 (100.0) | 2.87 (2.56-3.22, p<0.001) | 1.45 (1.28-1.63, p<0.001) |
| PDW_0fav (>8.9<16) | 0 | 10079 (100.0) | - | - |
| (<8.9>16) unf | 1 | 973 (100.0) | 1.84 (1.60-2.12, p<0.001) | 1.19 (1.03-1.37, p=0.017) |

**Fig 6.**
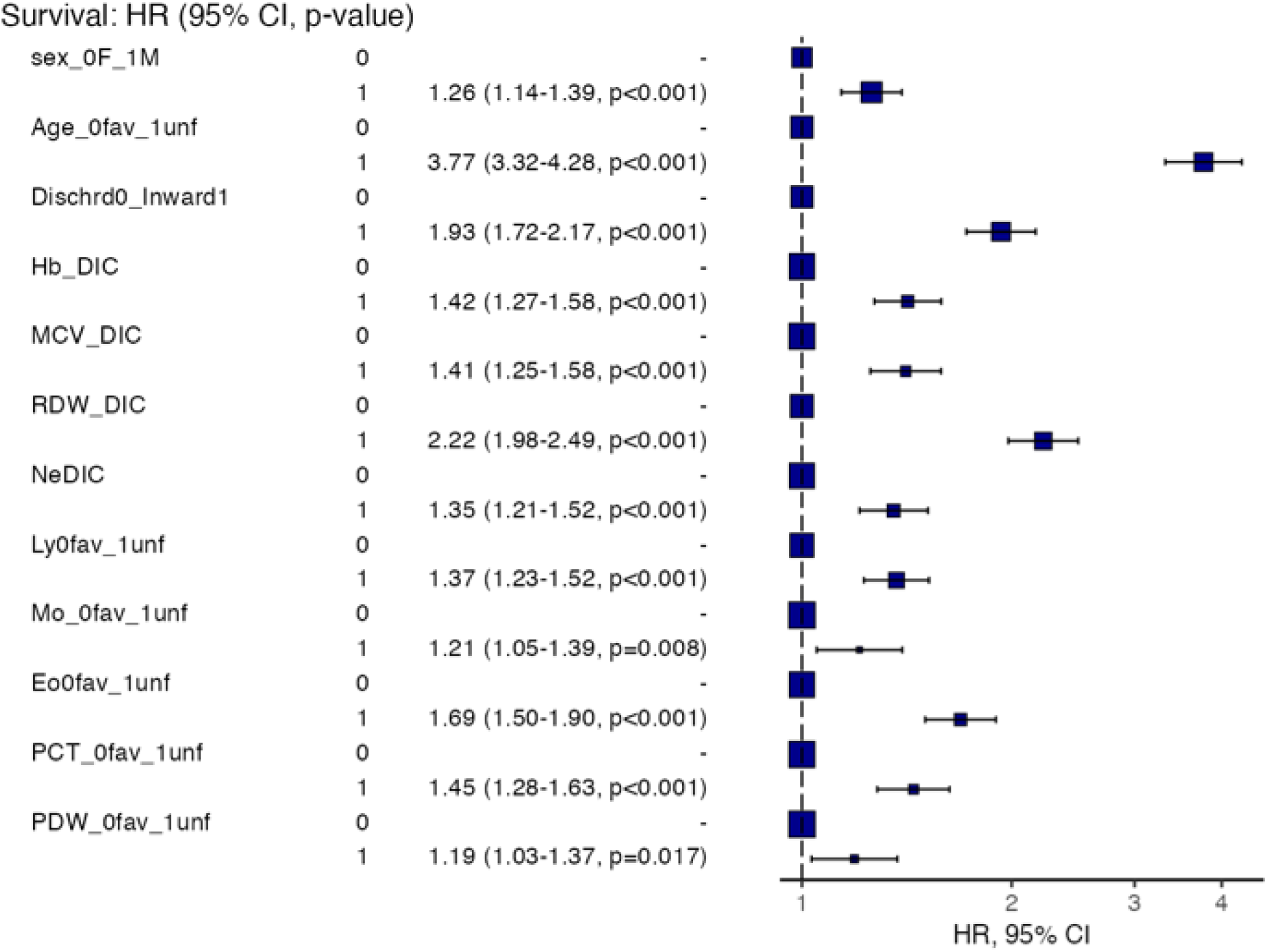
Hazards Regression Plot -Tailored BCDC

The models tested without the *Inward admission* explanatory variable were quite stable and superimposable for both the significant variables and their relative HR and CI values. However, only the age > 73 years HR increased in both models (see supplemental material Tab.S/5, Fig.S/5, Tab.S/6 Fig.S/6).

Among BCDC significant by Cox analysis elements, median number of unfavourable variables in alive and dead patients was 2 and 3 for Benchmark and 2 and 4 for Tailored discretization method, respectively. Number of unfavourable variables were classified for five equal groups and cutpoints were computed (Score Sum). The 20th, 40th, 60th and 80th percentiles were 1,2,3,4 points and 0,1,2,3 points for Benchmark and Tailored methods, respectively (Tab.7).

**Table 7.**
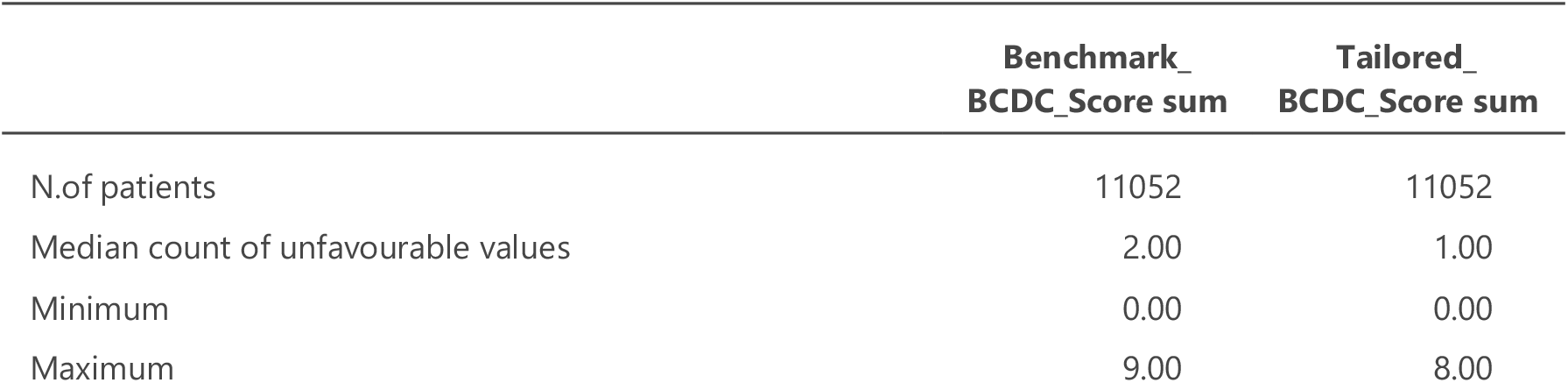
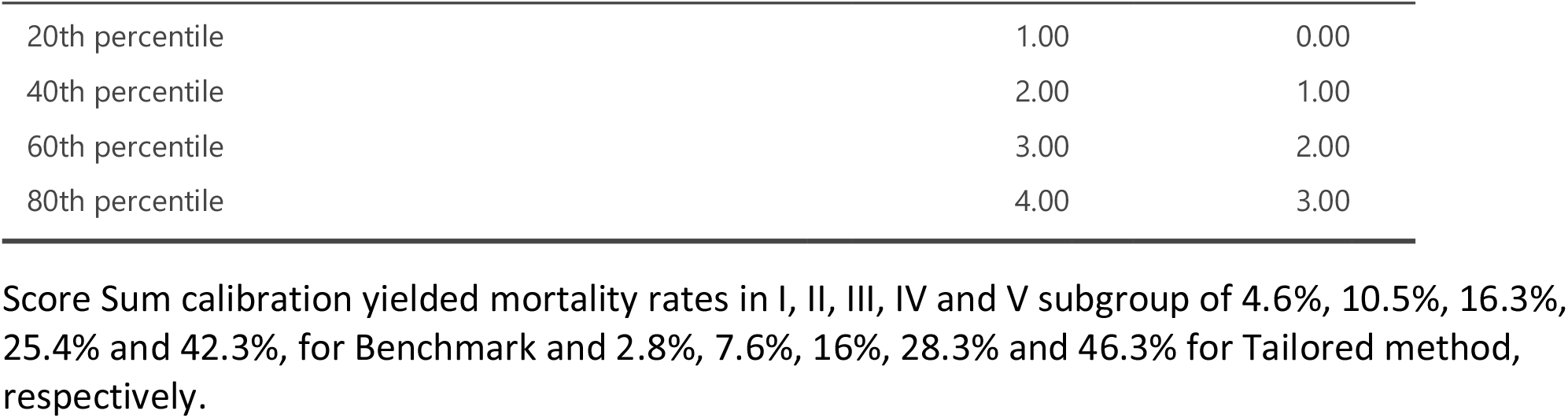
Score sum (count of unfavourable factors by quintiles) for each discretization method

The relationship between dead and alive in subgroups of patients stratified according to quintiles of Score Sum (tailored method) is shown in Fig.7.

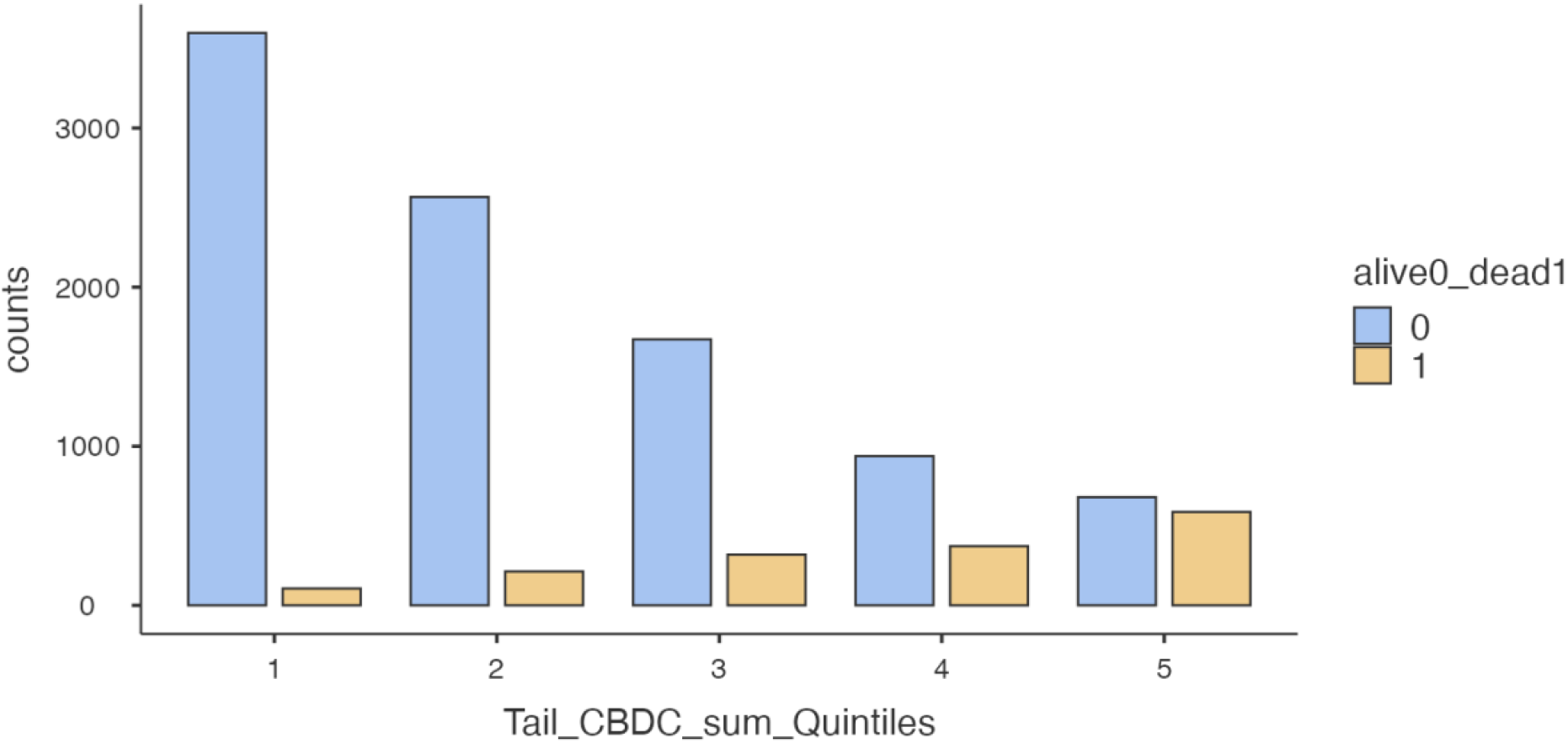

Actuarial curves of the whole population, stratified into five risk groups according to quintiles were calculated using the Kaplan–Meier method and compared by log–rank test. Events summary are detailed in Tab.8

**Table 8.** Events Summary by Tailored Score Sum (count of unfavourable BCDC values) Quintiles

| Quintiles | N, of Patients | Censored | Observed Events | Expected Events |
| --- | --- | --- | --- | --- |
| 1 | 3704 | 3598 | 106 | 577 |
| 2 | 2780 | 2567 | 213 | 418 |
| 3 | 1991 | 1672 | 319 | 286 |
| 4 | 1310 | 938 | 372 | 172 |
| 5 | 1267 | 680 | 587 | 144 |

A growing actuarial probability of death was observed from I to V quintile, as shown in Fig.8 for tailored model.

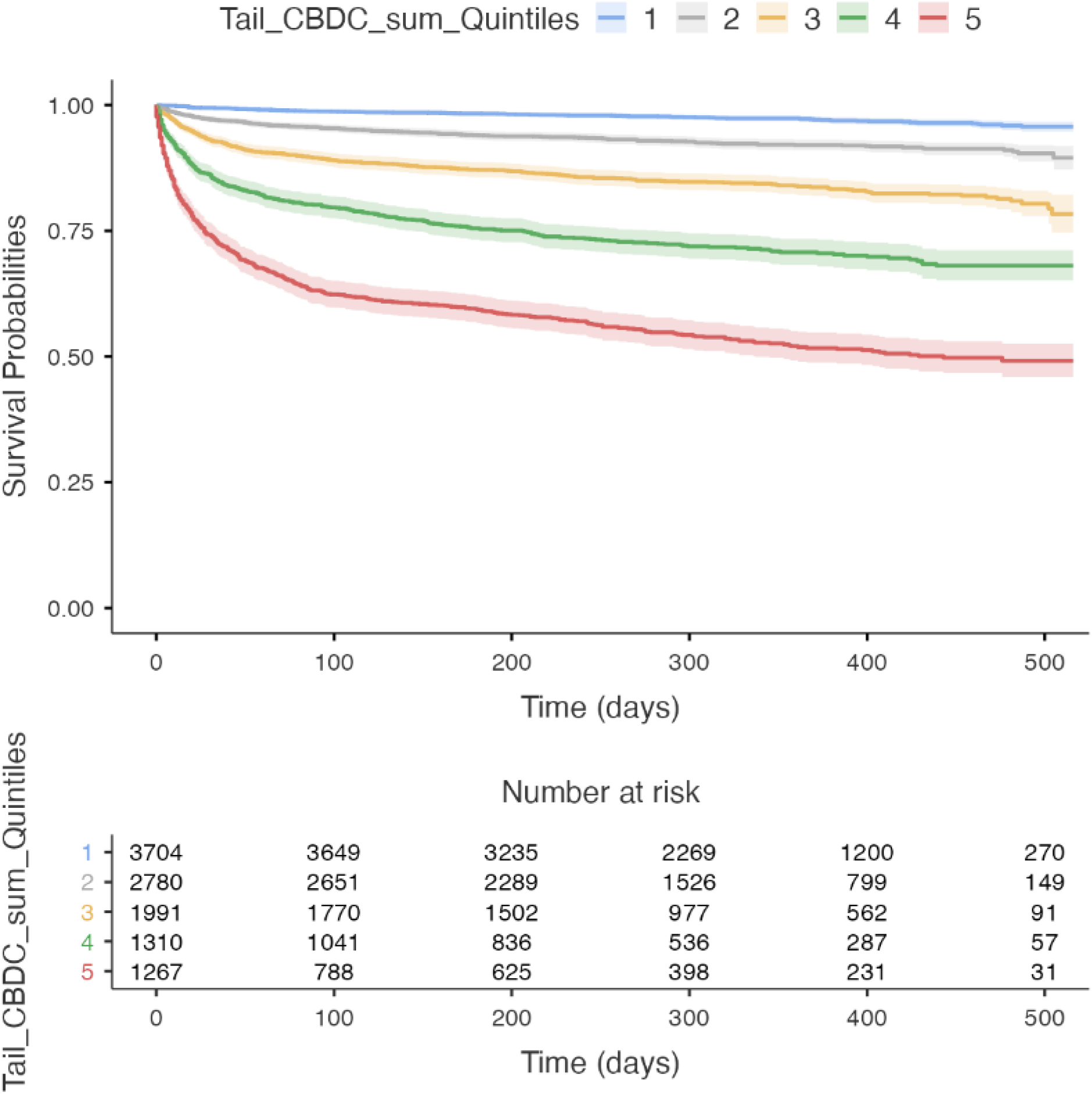

All details concerning score sum calibration in Benchmark and Tailored models are reported in supplemental material (Tab.S/7, Tab.S/8, Fig.S/7, Fig.S/8). Actuarial curves and log-rank test are reported in supplemental material (Tab.S/9, Tab.S/10, Fig.S/9, Fig.S/10).

Finally, we explored the predictive ability on survival of BCDC discretized using the Benchmark and Tailored methods. A ROC curve was generated by ranking the number of unfavourable BCDC variables for each discretization method.

Both curves demonstrated adequate predictivity on survival, with AUROCs of 74% and 79% for the Benchmark (including Hb, MCV, RDW, Ne, Ly, Eo, Ba, PLT, PDW) and Tailored model (including Hb, MCV, RDW, Ne, Ly, Eo, Mo, PCT, PDW), respectively. According to the DeLong test, the predictivity of Tailored method BCDC discretization was better than that of the Benchmark laboratory reference values (Supplemental materials, Table.S/11 Fig.S/11)

Inclusion of demographic variables (Age > 73 years, male sex) in the models increased AUROCs predictivity on survival from 74% to 79% and 79% to 82% for the Benchmark and Tailored discretization method, respectively (Table 9 and Fig.9). Further details are in Supplemental material (Tab.S/12, Fig.S/12).

**Table 9.**
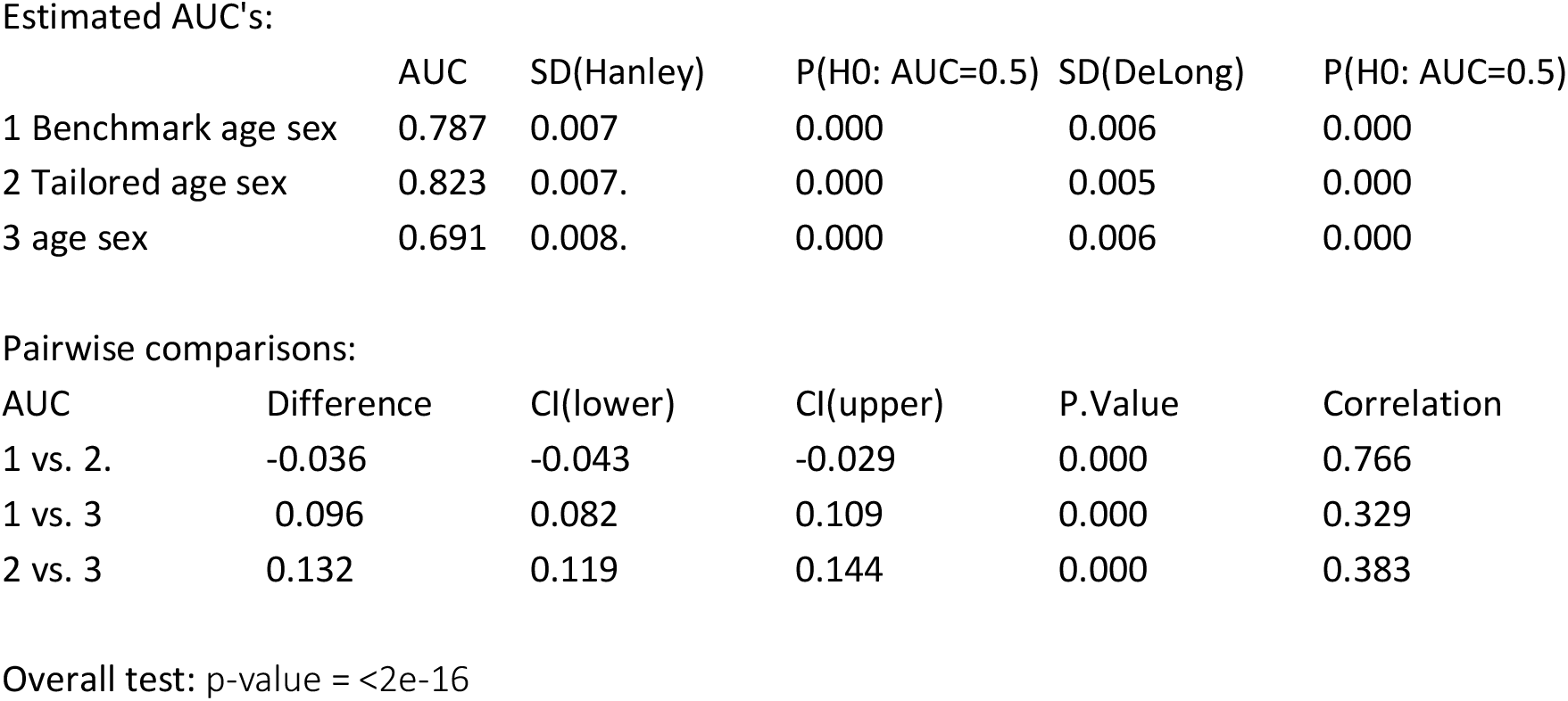
DeLong Test of Difference between AUCs

Estimated AUC's:
|  | AUC | SD(Hanley) | P(H0: AUC=0.5) | SD(DeLong) | P(H0: AUC=0.5) |
| --- | --- | --- | --- | --- | --- |
| 1 Benchmark age sex | 0.787 | 0.007 | 0.000 | 0.006 | 0.000 |
| 2 Tailored age sex | 0.823 | 0.007. | 0.000 | 0.005 | 0.000 |
| 3 age sex | 0.691 | 0.008. | 0.000 | 0.006 | 0.000 |

Pairwise comparisons:

**Fig 9.**
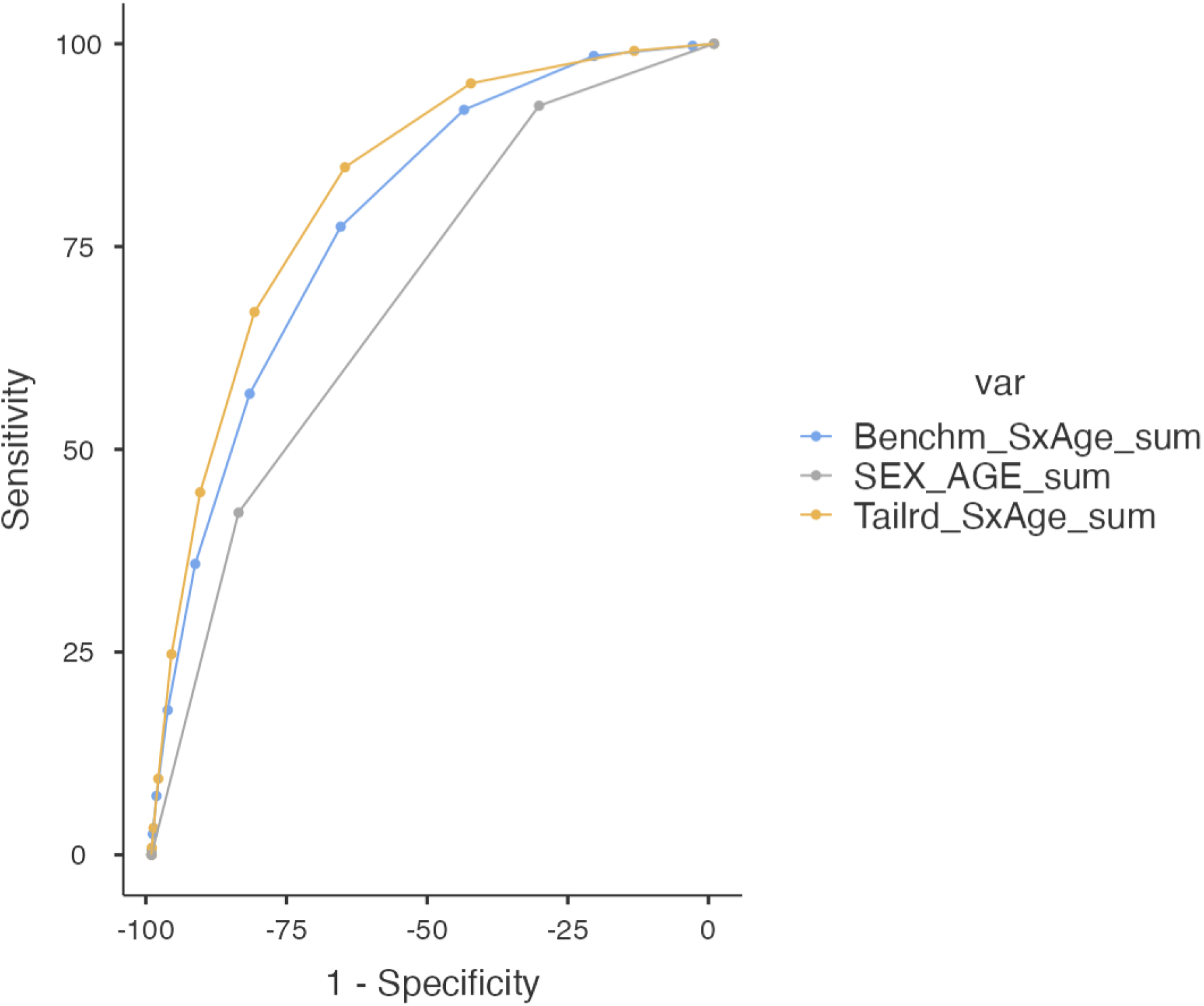
ROC Curves for: Benchmark BCDC+Age+Sex, Tailored BCDC+Age+Sex and Age+Sex

## Discussion

Automated complete blood differential count represents a simple, low cost and easily performed assessment of a patient’s general and immune status in emergency settings. However, its predictive ability on survival remains to be thoroughly assessed.

This retrospective study on patients referring to our ER during 2020 showed that selected elements from a single baseline blood sample, taken at initial presentation for most acute disease or trauma, was efficient in predicting one-year survival, independently from demographic parameters (age and sex) and from ER medical staff decision to discharge or hospitalize the patient.

The laboratory reference values defined on general population intervals was narrow compared with those computed using Tailored methods on HR. The HRs of each significant variable and AUROC overall predictivity was lower considering the interval was based on the general population instead of targeted at survival in a general population subset, such as acutely ill patients.

Concerning the Tailored discretization method, the OEHR methodology[6] was very helpful for discretization of U-shaped variables, which are biologically relevant in health and disease. This method targeted value intervals not purely based on frequency in the general population for each BCDC element. Additionally, it was more methodologically fitted and did not rely on a fixed a priori threshold but was specifically based on HR. Furthermore, the OEHR method proved useful for discretization of variables characterized by low cell count within BCDC (Eo and Ba count), which are flawed by intrinsic inaccuracy in automated count.[15,16] Clinically significant thresholds largely consisted in either absence (< 0.01 ×10^9^/L) or consistent presence of Eo or Ba in peripheral blood.

Both methods for value discretization exhibited an acceptable performance on survival predictivity, although they were had an explorative purpose.

Moreover, it is clear that the increasing number of unfavorable BCDC variables at baseline is associated with increased mortality risk, as demonstrated by both models. Beyond statistical methodology implications, this would represent biological insight into marrow disruption as well as spleen and lymphatic tissue response. Both these processes are often hurt during acute illness or trauma, and their extent and severity degrees are likely linked with poor outcomes.

A major study limitation was lack of data regarding patient performance status, symptoms, vital signs and disease diagnosis, which are required to assess the relevance of BCDC for outcome predictions in comparison with other laboratory or clinical parameters. For this reason, among explanatory variables we introduced the inward admission in addition to age and sex.

Nonetheless, this variable is usually considered an outcome separate from survival in clinical studies. In this study setting, such variable could roughly surrogate a more severe clinical condition, and/or clinically relevant instrumental findings, resulting in medical staff decision to admit the patient to the hospital. The Cox analysis repeated with and without the inward admission explanatory variable yielded superimposable results, suggesting that the model is robust with respect to the removal of this variable (Supplemental material Tab.S/5,Tab.S/6,Fig.S/5,Fig.S/6).

Another concern relates to the Coronavirus disease of 2020 (COVID-19) pandemic, which affected the number of patients referring to the ER, hospitalization rates, and the mortality rate in 2020. The italian region Lombardia, where our hospital is located, underwent two COVID-19 waves during 2020, resulting in two peaks in hospital admissions of approximately 13.000 patients in early April and 10.000 in November[17], out of a population of 10 million people (45% over 50 years-old).

The influx of COVID-19 patients in our hospital during 2020 might have skewed the results of this study. Consequently, the discretized BCDC value intervals identified as favorable or unfavorable in this population may be unsuitable if applied in a different timeframe. Nevertheless, useful insight emerged from this retrospective monocentric study. First, thoroughly investigating BCDC variables in acutely ill patients demonstrated usefulness for outcome predictions, as we found for counts of Ne, Ly, Mo, Eo, Ba, and for values of RDW, MCV, and PDW. Nonetheless, general use variables (i.e., total WBC count and PLT count) were excluded using the multivariable Cox analysis. Such a finding warrants cautious consideration and could be a signal that in acute settings, conditions frequently exist in which high RDW, lymphopenia and/or eosinopenia reflect severe suffering, leading to increased mortality risks.

In longitudinal studies lymphopenia is associated with increased mortality risk for all causes[18, 19]. Lymphocytopenia in severely ill patients[20] was a better predictor of bacteremia in comparison to the total leukocyte and Ne count in a cohort of 21.372 cases. Lymphopenia was a predictor of illness severity and short-term mortality risk in a cohort of 58.260 patients admitted to the hospital[21]. Lymphopenia in chronic diseases is an unfavorable independent variable, including in cancer[22, 23,24]. In prospective studies, lymphopenia was associated with increased risk of all cause specific mortality[19] and was additive to traditional risk factors[18]. Extreme lymphocytosis was noticed in hematologic disease and in patients with central nervous system (CNS) bleeding or head trauma with poor prognosis[1].

Concerning RDW and lymphopenia, a recent study on 1641 SARS-CoV2 hospitalized patients demonstrated that high RDW at baseline and subsequent RDW elevation increased in-hospital mortality. This finding is not restricted to COVID-19, as it was described in a retrospective study on 1.715 chronic Hepatitis C virus (HCV) patients undergoing a five-year follow up[25]. Lymphopenia and elevated RDW were associated with long-term mortality risk in 15179 patients undergoing coronary angiography in both acute and non-acute settings[26].

Eosinopenia was reported as a marker of poor outcome in lung disease[27] and critically ill patients[28]. Absolute eosinopenia was associated with clinically poor outcomes in firstwave COVID-19 pneumonia[7, 29].

In experimental rabbit models, lymphocytes are redistributed from peripheral blood to lymphatic tissue after cortisol administration, for instance, following major surgery. *Escherichia coli* endotoxemia and surgery were accompanied by lymphocytopenia and increased cortisol. Conversely, lymphocytes are redistributed from the spleen and bone marrow to peripheral blood, lungs, and liver after adrenaline infusion[30,31,32]. From a pathophysiological perspective, lymphopenia, including eosinopenia, neutrophilia and increased RDW[33] may be markers and drivers of an ineffective response to major health-disturbing events.

Our study represents a straightforward explorative attempt to accurately assess the acute patient immune status at presentation, enabling the detection of at-risk patients. Further investigation should include prognostic stratification and evenly tailored diagnostic and therapeutic pathways. This introductory model cannot be considered ready for clinical application, but a basis for further clinical research addressing whether baseline BCDC can generate reliable and specific biomarkers for premature detection of specific acute conditions. Finally, long-term therapeutic research should explore whether is it possible to actively manage host immunity by reproducing an effective response, to improve the overall outcome.

## Conclusion

Values from automated peripheral BCDC counts taken at baseline in adult patients visiting our emergency room during 2020 were discretized using laboratory reference values (Benchmark) or OEHR and MSRS (Tailored). Variables were adequately efficient and robust in predicting one-year survival, independently from demographic (age and sex) and ER medical staff decision to discharge or hospitalize patients. Tailored discretization of Hb, MCV, RDW, Ne, Ly, Mo, Eo, PCT and PDW yielded more accurate survival predictions than Benchmark laboratory reference interval in our cohort of patients.

Further studies are warranted to validate these findings and explore whether specific BCDC patterns can predict outcomes of single acute diseases or conditions.

## Data Availability

All data produced in the present study are available upon reasonable request to the authors

## Acknowledgments

We are very grateful to A.Manzoni Hospital units of “Pronto Soccorso” and “Dipartimento Servizi Clinici” graduate, nursing and technical staff; to Dr. M.Tavola, and G. Baraldo for their wise hints; to Dr. M. Biffi and Dr. C. Curioni for their factual help; to Dr. G.Nattino (Unit of Causal Inference in Epidemiology, Istituto Mario Negri IRCCS) for invaluable advice. We would like to thank Editage (www.editage.com) for English language editing.

## Competing Interests

none declared

## Funding

none

## Notes

### Competing Interest Statement

The authors have declared no competing interest.

### Funding Statement

This study did not receive any funding

### Author Declarations

IRB of ASST Lecco gave ethical approval for this work on 2021-07-14 by deliberation no 566

