## Supplemental materials for "Blood cell differential count discretization methods to predict survival in acutely ill adults reporting to the emergency room: a retrospective cohort study in 2020"

#### Table S/1 Descriptive analysis of BCDC and demographic variables

| | Age yrs | Hb | RBC | MCV | RDW | WBC | Ne | Ly | Mo | Eo | Ba | PLT | PCT | PDW |
|---|---|---|---|---|---|---|---|---|---|---|---|---|---|---|
| N | 11052 | 11052 | 11051 | 11052 | 11052 | 11052 | 11052 | 11052 | 11052 | 11052 | 11052 | 11052 | 11052 | 11052 |
| Mean | 64.2 | 13.5 | 4.53 | 90.2 | 13.7 | 9.24 | 6.74 | 1.68 | 0.673 | 0.105 | 0.034 | 232 | 0.241 | 12.4 |
| Median | 67.0 | 13.7 | 4.57 | 90.2 | 13.2 | 8.48 | 5.82 | 1.52 | 0.630 | 0.060 | 0.030 | 222 | 0.230 | 12.1 |
| SD | 19.7 | 2.04 | 0.684 | 6.62 | 1.76 | 3.92 | 3.73 | 0.944 | 0.305 | 0.134 | 0.023 | 81.8 | 0.079 | 2.27 |
| Min | 17 | 2.70 | 0.600 | 54.1 | 10.7 | 0.360 | 0.030 | 0.050 | 0.00 | 0.00 | 0.00 | 2 | 0.00 | 3.70 |
| Max | 99 | 20.7 | 8.32 | 148 | 30.1 | 48.1 | 29.5 | 9.44 | 2.49 | 1.48 | 0.240 | 1023 | 1.01 | 25.3 |
| Skewness | -0.509 | -0.750 | -0.499 | -0.103 | 2.33 | 1.50 | 1.59 | 1.45 | 1.13 | 3.19 | 1.54 | 1.59 | 1.51 | 1.11 |
| Std. error skewness | 0.023 | 0.023 | 0.023 | 0.023 | 0.023 | 0.023 | 0.023 | 0.023 | 0.023 | 0.023 | 0.023 | 0.023 | 0.023 | 0.023 |
| Kurtosis | -0.658 | 1.38 | 1.43 | 4.57 | 8.66 | 4.27 | 3.71 | 4.62 | 2.57 | 17.2 | 5.36 | 7.30 | 6.90 | 2.24 |
| Std. error kurtosis | 0.046 | 0.046 | 0.046 | 0.046 | 0.046 | 0.046 | 0.046 | 0.046 | 0.046 | 0.046 | 0.046 | 0.046 | 0.046 | 0.046 |

Abbreviations: Hemoglobin (Hb), mean red cell volume (MCV), red cell distribution width (RDW), platelet distribution width (PDW), platelet hematocrit (PCT) and absolute count of red blood cells (RBC), white blood cells (WBC), neutrophils (Ne), lymphocytes (Ly), monocytes (Mo), eosinophils (Eo), basophils (Ba), and platelets (PLT), standard deviation (SD), maximum (max), minimum (min).

### Fig S/1 Density and Q-Q Plots of BCDC and demographic variables by Alive/Dead status

Age

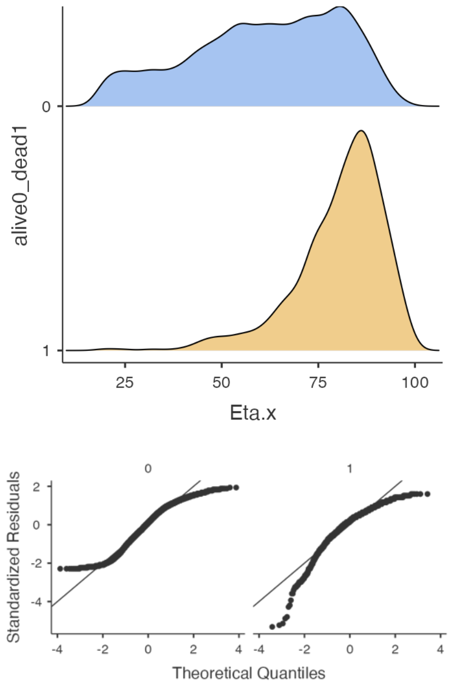


RBC

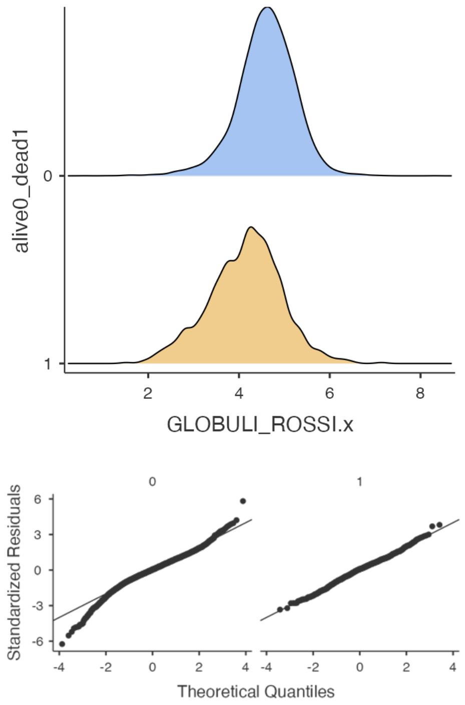


Hb

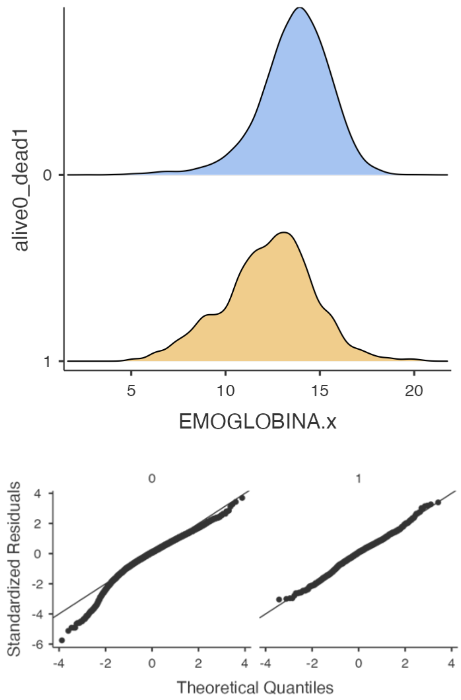


MCV

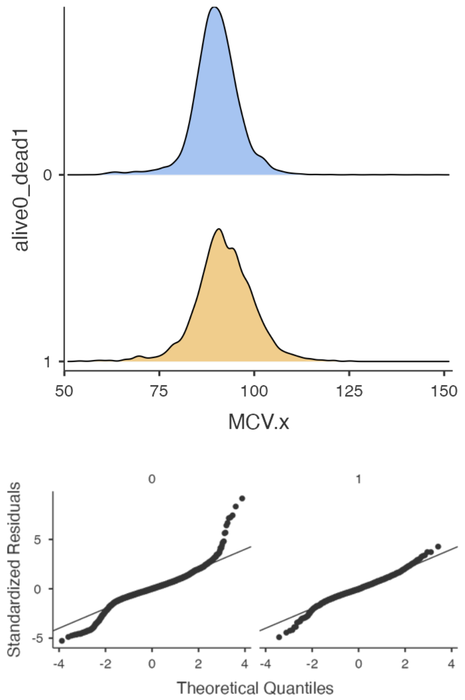

RDW

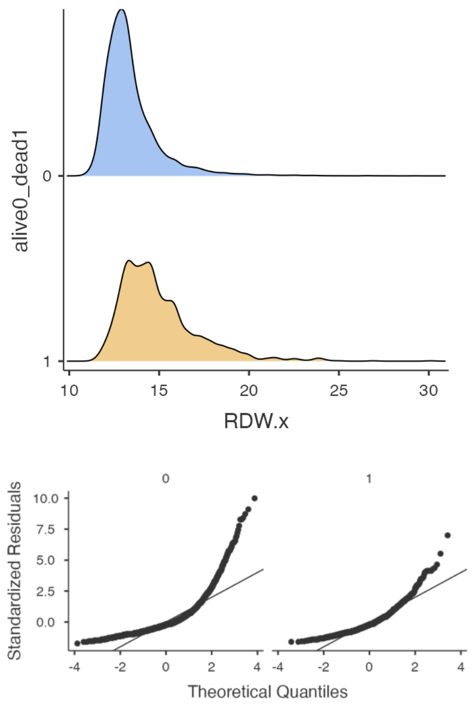


Neutrophils

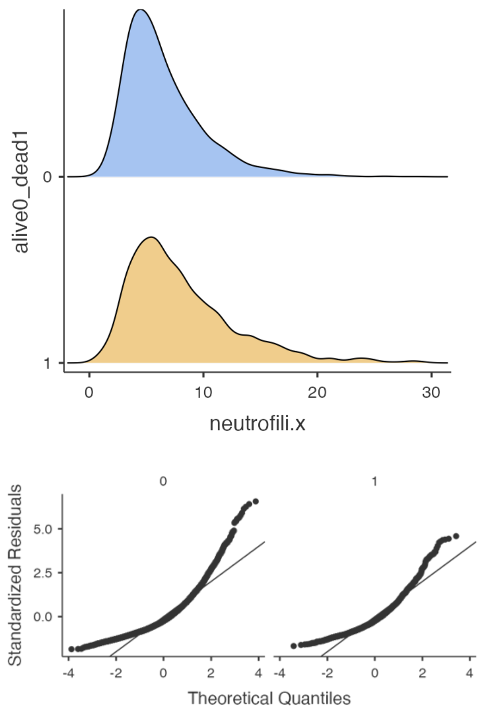


WBC

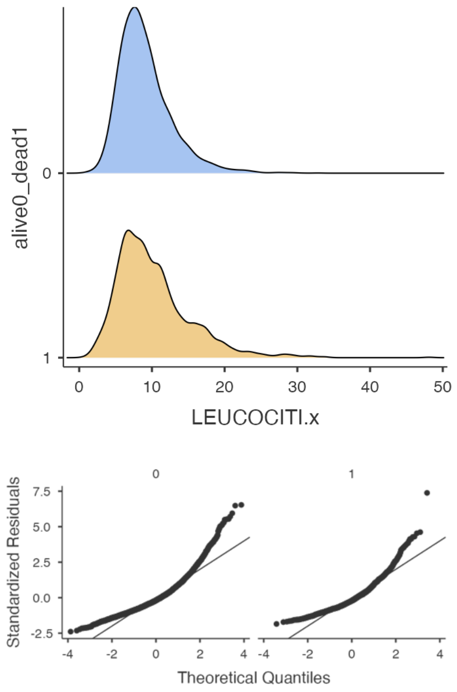


Lymphocytes

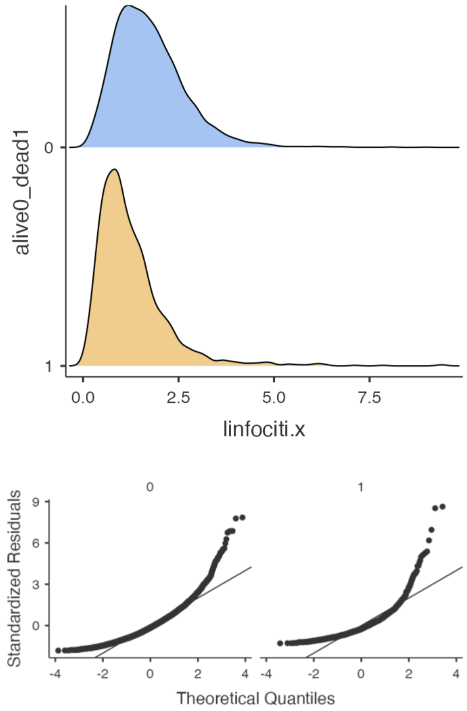

#### Monocytes

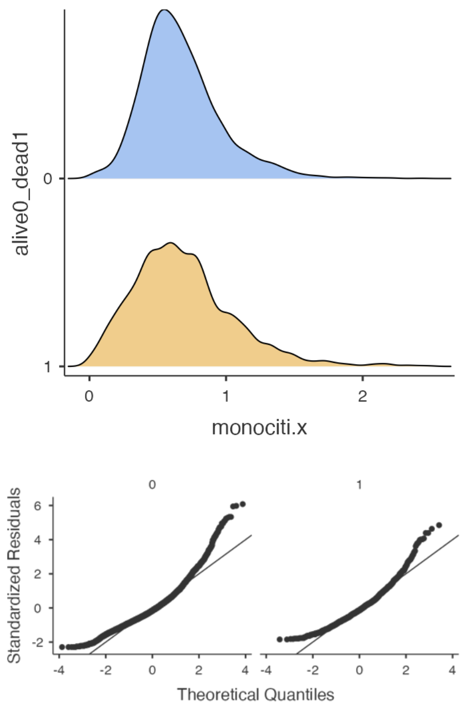


#### Basophils

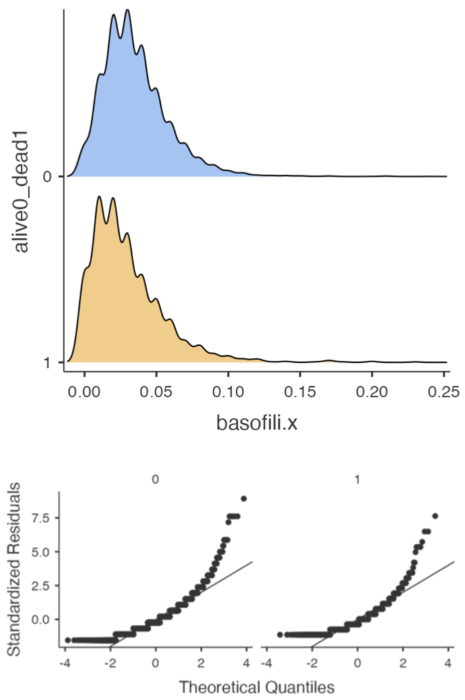


#### Eosinophils

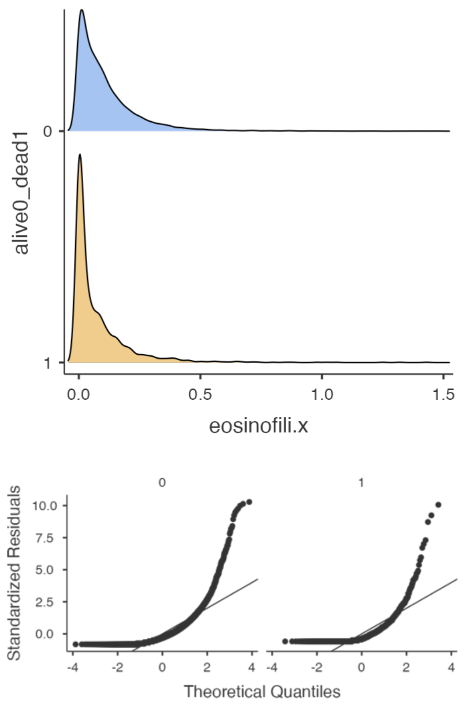


#### Platelets

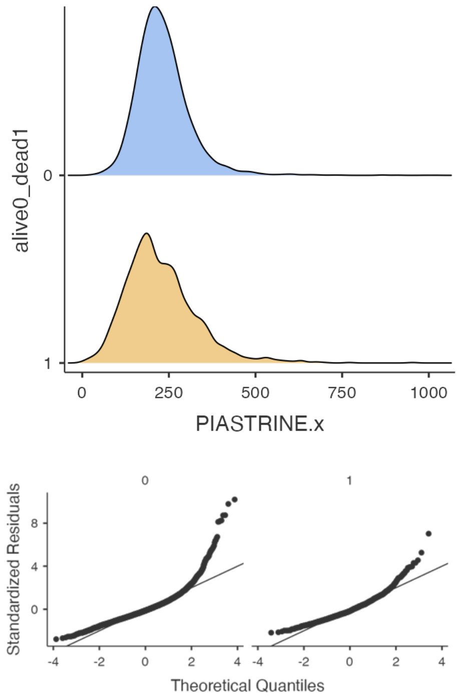

PCT

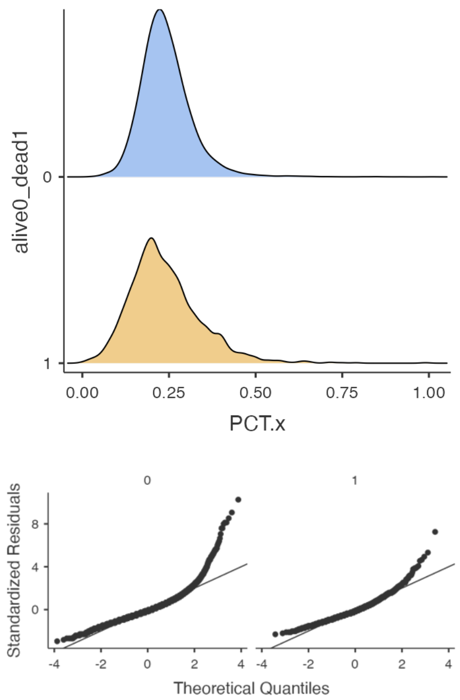
alive0_dead1
0
1
0.00
0.25
0.50
0.75
1.00
PCT.x
0
1
Standardized Residuals
8
4
0
-4
-2
0
2
4
Theoretical Quantiles

PDW

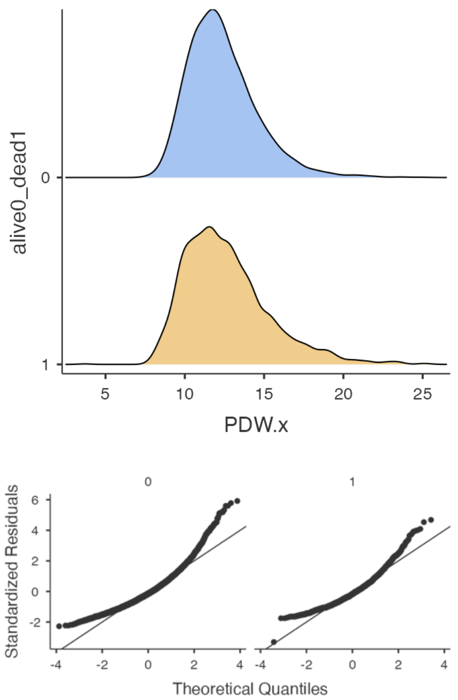
alive0_dead1
0
1
5
10
15
20
25
PDW.x
0
1
Standardized Residuals
6
4
2
0
-2
-4
-2
0
2
4
Theoretical Quantiles

**Table S/2 Independent samples T-test results (Alive vs Deceased patients)**

#### Table S/2a BCDC Independent Samples T-Test by Alive vs Dead

Independent Samples T-Test

| | | Statistic | df | p |
|---|---|---|---|---|
| Hemoglobin | Student's t | 25.77 [a] | 11050 | < .001 |
| | Mann-Whitney U | 4.76e+6 | | < .001 |
| RBC | Student's t | 25.38 [a] | 11049 | < .001 |
| | Mann-Whitney U | 4.84e+6 | | < .001 |
| MCV | Student's t | -12.30 [a] | 11050 | < .001 |
| | Mann-Whitney U | 6.08e+6 | | < .001 |
| RDW | Student's t | -33.59 [a] | 11050 | < .001 |
| | Mann-Whitney U | 3.67e+6 | | < .001 |
| WBC | Student's t | -10.62 [a] | 11050 | < .001 |
| | Mann-Whitney U | 6.78e+6 | | < .001 |
| neutrofili | Student's t | -15.53 [a] | 11050 | < .001 |
| | Mann-Whitney U | 6.05e+6 | | < .001 |
| linfociti | Student's t | 18.81 [a] | 11050 | < .001 |
| | Mann-Whitney U | 4.81e+6 | | < .001 |
| monociti | Student's t | -1.89 [a] | 11050 | 0.058 |
| | Mann-Whitney U | 7.53e+6 | | 0.881 |
| eosinofili | Student's t | 8.21 | 11050 | < .001 |
| | Mann-Whitney U | 5.66e+6 | | < .001 |
| basofili | Student's t | 8.75 [a] | 11050 | < .001 |
| | Mann-Whitney U | 6.12e+6 | | < .001 |
| PLT | Student's t | 2.45 [a] | 11050 | 0.014 |
| | Mann-Whitney U | 6.94e+6 | | < .001 |
| PCT | Student's t | 1.14 [a] | 11050 | 0.254 |
| | Mann-Whitney U | 7.07e+6 | | < .001 |
| PDW | Student's t | -4.42 [a] | 11050 | < .001 |
| | Mann-Whitney U | 7.31e+6 | | 0.045 |
| Age | Student's t | -38.12 [a] | 11050 | < .001 |
| | Mann-Whitney U | 3.03e+6 | | < .001 |

[a] Levene's test is significant ($p < .05$), suggesting a violation of the assumption of equal variances
Abbreviations: Hemoglobin (Hb), mean red cell volume (MCV), red cell distribution width (RDW), platelet distribution width (PDW), platelet hematocrit (PCT) and absolute count of red blood cells (RBC), white blood cells (WBC), neutrophils (Ne), lymphocytes (Ly), monocytes (Mo), eosinophils (Eo), basophils (Ba), and platelets (PLT), degree of freedom (df)

### Table S/2b Assumptions

Alive vs Dead

Homogeneity of Variances Tests

| | | F | df | df2 | p |
|---|---|---|---|---|---|
| EMOGLOBINA.x | Levene's | 160.72 | 1 | 11050 | < .001 |
| | Variance ratio | 0.635 | 9454 | 1596 | < .001 |
| GLOBULI_ROSSI.x | Levene's | 151.81 | 1 | 11049 | < .001 |
| | Variance ratio | 0.640 | 9453 | 1596 | < .001 |
| MCV.x | Levene's | 107.97 | 1 | 11050 | < .001 |
| | Variance ratio | 0.673 | 9454 | 1596 | < .001 |
| RDW.x | Levene's | 280.02 | 1 | 11050 | < .001 |
| | Variance ratio | 0.532 | 9454 | 1596 | < .001 |
| LEUCOCITI.x | Levene's | 260.97 | 1 | 11050 | < .001 |
| | Variance ratio | 0.504 | 9454 | 1596 | < .001 |
| neutrofili.x | Levene's | 204.01 | 1 | 11050 | < .001 |
| | Variance ratio | 0.560 | 9454 | 1596 | < .001 |
| linfociti.x | Levene's | 14.53 | 1 | 11050 | < .001 |
| | Variance ratio | 0.961 | 9454 | 1596 | 0.289 |
| monociti.x | Levene's | 139.58 | 1 | 11050 | < .001 |
| | Variance ratio | 0.619 | 9454 | 1596 | < .001 |
| eosinofili.x | Levene's | 3.53 | 1 | 11050 | 0.060 |
| | Variance ratio | 0.986 | 9454 | 1596 | 0.704 |
| basofili.x | Levene's | 12.56 | 1 | 11050 | < .001 |
| | Variance ratio | 0.766 | 9454 | 1596 | < .001 |
| PIASTRINE.x | Levene's | 219.75 | 1 | 11050 | < .001 |
| | Variance ratio | 0.562 | 9454 | 1596 | < .001 |
| PCT.x | Levene's | 266.85 | 1 | 11050 | < .001 |
| | Variance ratio | 0.523 | 9454 | 1596 | < .001 |
| PDW.x | Levene's | 106.24 | 1 | 11050 | < .001 |
| | Variance ratio | 0.653 | 9454 | 1596 | < .001 |
| Eta.x | Levene's | 743.14 | 1 | 11050 | < .001 |
| | Variance ratio | 2.815 | 9454 | 1596 | < .001 |

Note. Additional results provided by *moretests*

#### Table S/2c Tests of Normality

| | | statistic | p |
|---|---|---|---|
| EMOGLOBINA.x | Shapiro-Wilk | NaN | |
| | Kolmogorov-Smirnov | 0.0543 | < .001 |
| | Anderson-Darling | 55.7 | < .001 |
| GLOBULI_ROSSI.x | Shapiro-Wilk | NaN | |
| | Kolmogorov-Smirnov | 0.0392 | < .001 |
| | Anderson-Darling | 33.9 | < .001 |
| MCV.x | Shapiro-Wilk | NaN | |
| | Kolmogorov-Smirnov | 0.0642 | < .001 |
| | Anderson-Darling | 100.5 | < .001 |
| RDW.x | Shapiro-Wilk | NaN | |
| | Kolmogorov-Smirnov | 0.1540 | < .001 |
| | Anderson-Darling | 465.6 | < .001 |
| LEUCOCITI.x | Shapiro-Wilk | NaN | |
| | Kolmogorov-Smirnov | 0.0893 | < .001 |
| | Anderson-Darling | 197.4 | < .001 |
| neutrofili.x | Shapiro-Wilk | NaN | |
| | Kolmogorov-Smirnov | 0.1076 | < .001 |
| | Anderson-Darling | 284.1 | < .001 |
| linfociti.x | Shapiro-Wilk | NaN | |
| | Kolmogorov-Smirnov | 0.0733 | < .001 |
| | Anderson-Darling | 158.9 | < .001 |
| monociti.x | Shapiro-Wilk | NaN | |
| | Kolmogorov-Smirnov | 0.0775 | < .001 |
| | Anderson-Darling | 132.8 | < .001 |
| eosinofili.x | Shapiro-Wilk | NaN | |
| | Kolmogorov-Smirnov | 0.2069 | < .001 |
| | Anderson-Darling | 718.3 | < .001 |
| basofili.x | Shapiro-Wilk | NaN | |
| | Kolmogorov-Smirnov | 0.1534 | < .001 |
| | Anderson-Darling | 234.5 | < .001 |
| PIASTRINE.x | Shapiro-Wilk | NaN | |
| | Kolmogorov-Smirnov | 0.0736 | < .001 |
| | Anderson-Darling | 137.4 | < .001 |
| PCT.x | Shapiro-Wilk | NaN | |
| | Kolmogorov-Smirnov | 0.0916 | < .001 |
| | Anderson-Darling | 136.3 | < .001 |

#### Table S/2c Tests of Normality

| | | statistic | p |
|---|---|---|---|
| PDW.x | Shapiro-Wilk | NaN | |
| | Kolmogorov-Smirnov | 0.0762 | < .001 |
| | Anderson-Darling | 121.1 | < .001 |
| Eta.x | Shapiro-Wilk | NaN | |
| | Kolmogorov-Smirnov | 0.0592 | < .001 |
| | Anderson-Darling | 76.6 | < .001 |

Note. Additional results provided by *moretests*

#### Figure S/2 BCDC descriptive statistics plots by Alive dead

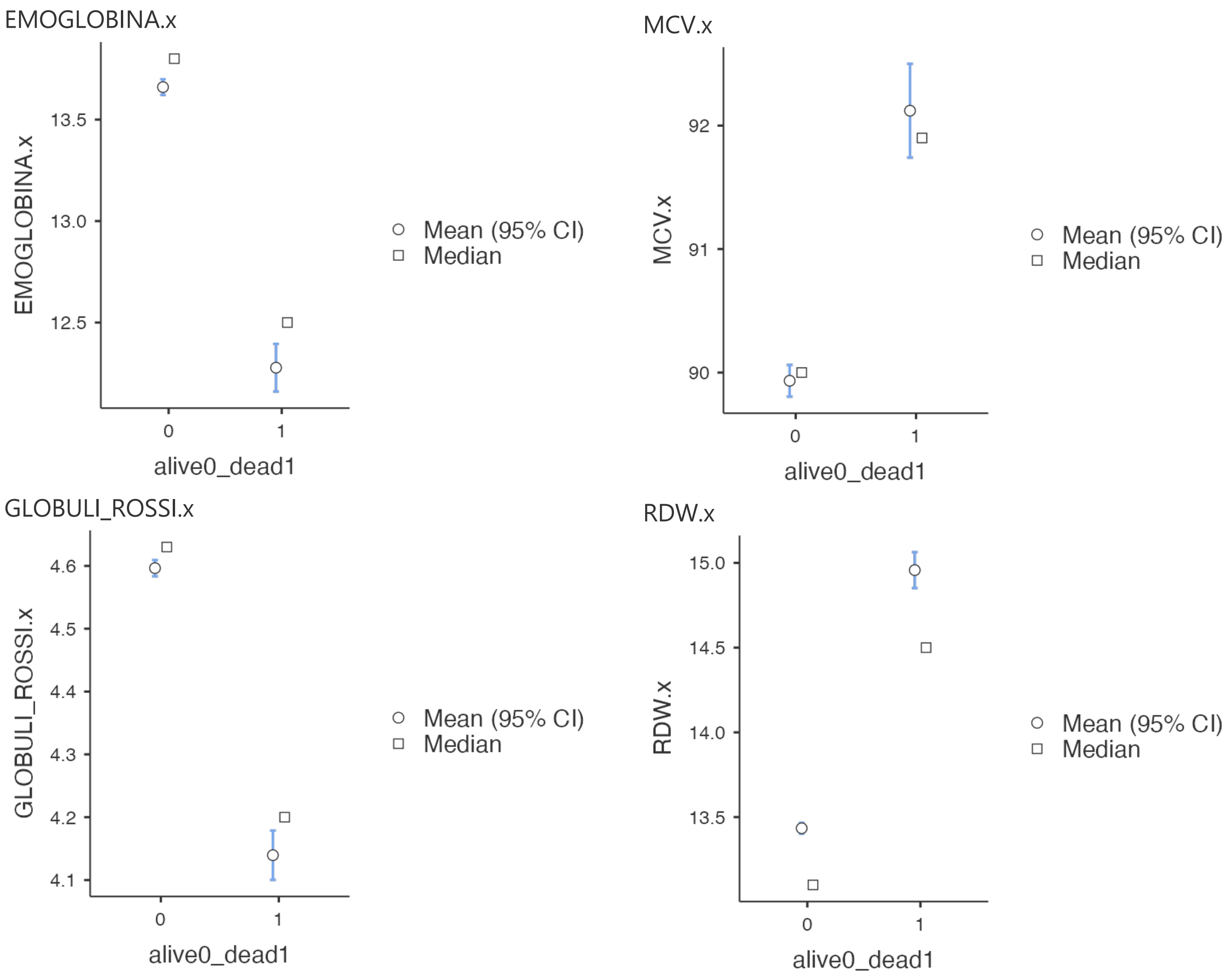

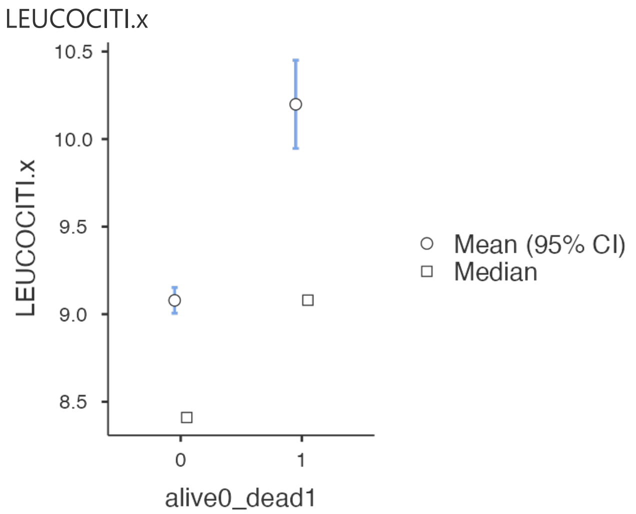
LEUCOCITI.x
10.5
10.0
9.5
9.0
8.5
LEUCOCITI.x
0
1
alive0_dead1
Mean (95% CI)
Median

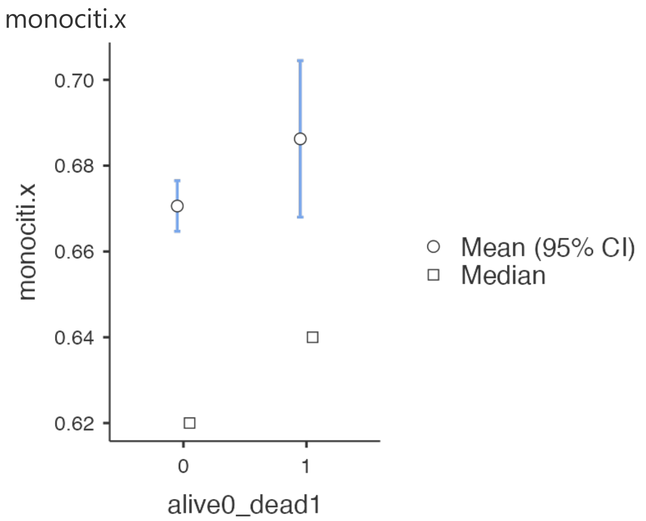
monociti.x
0.70
0.68
0.66
0.64
0.62
monociti.x
0
1
alive0_dead1
Mean (95% CI)
Median

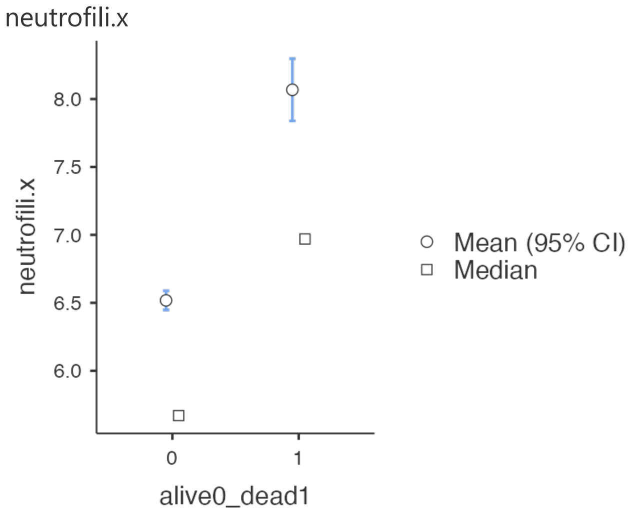
neutrofili.x
8.0
7.5
7.0
6.5
6.0
neutrofili.x
0
1
alive0_dead1
Mean (95% CI)
Median

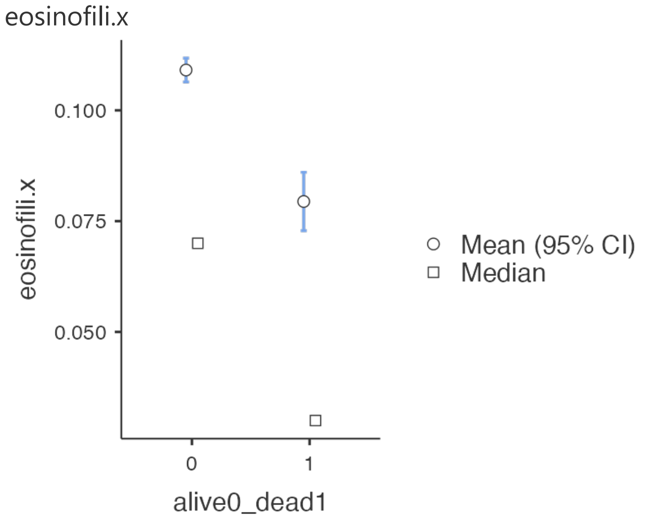
eosinofili.x
0.100
0.075
0.050
eosinofili.x
0
1
alive0_dead1
Mean (95% CI)
Median

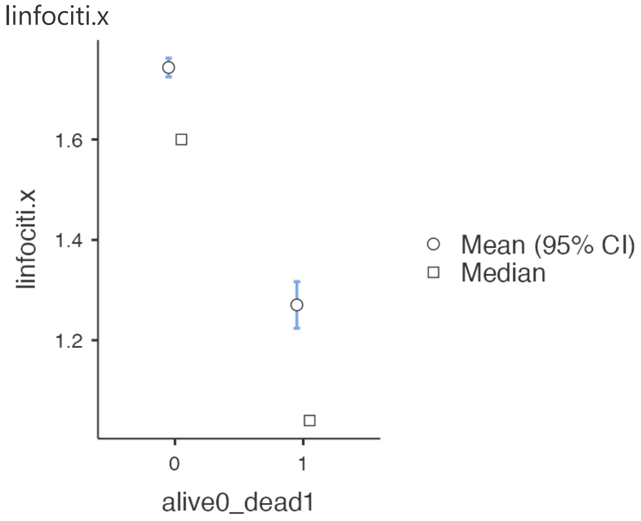
linfociti.x
1.6
1.4
1.2
linfociti.x
0
1
alive0_dead1
Mean (95% CI)
Median

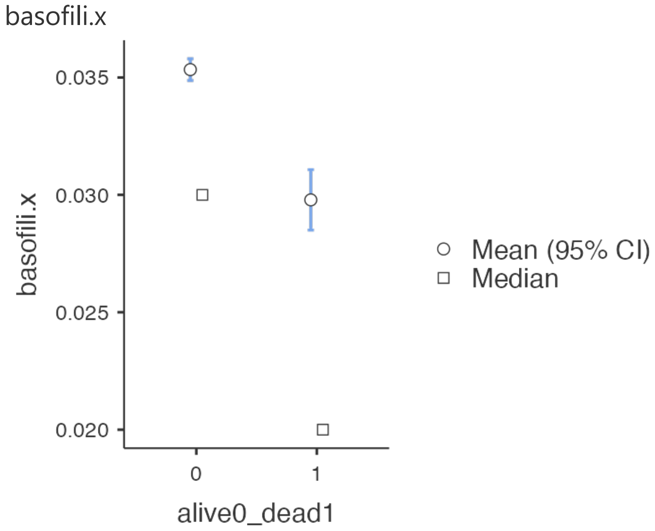
basofili.x
0.035
0.030
0.025
0.020
basofili.x
0
1
alive0_dead1
Mean (95% CI)
Median

PIASTRINE.x

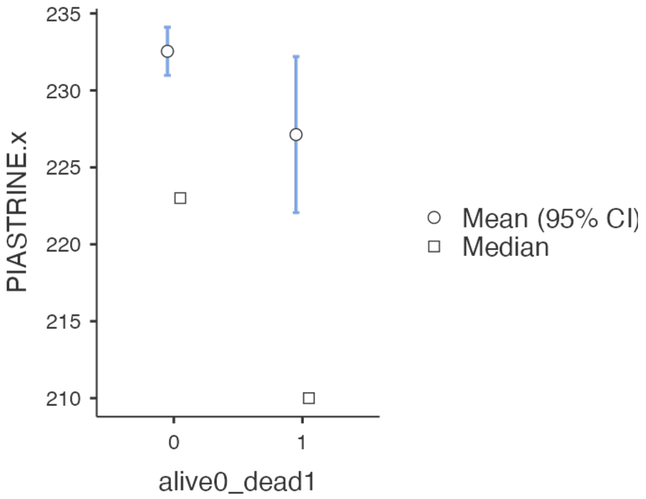


PDW.x

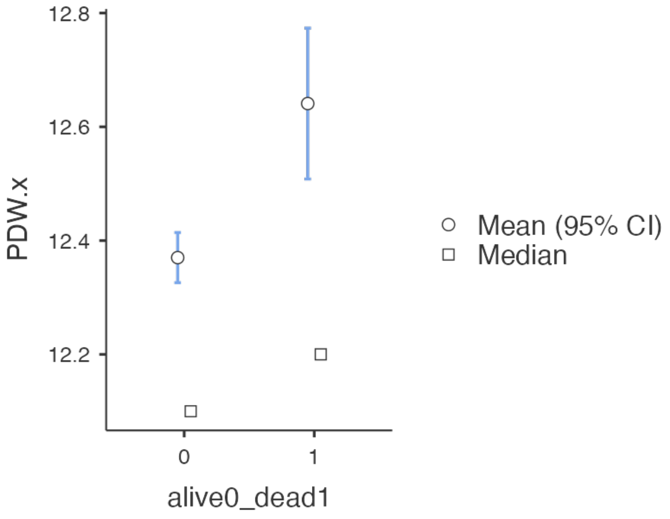


PCT.x

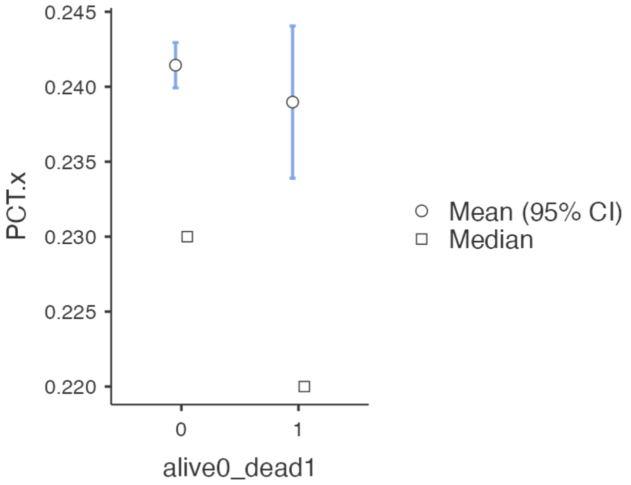


Age

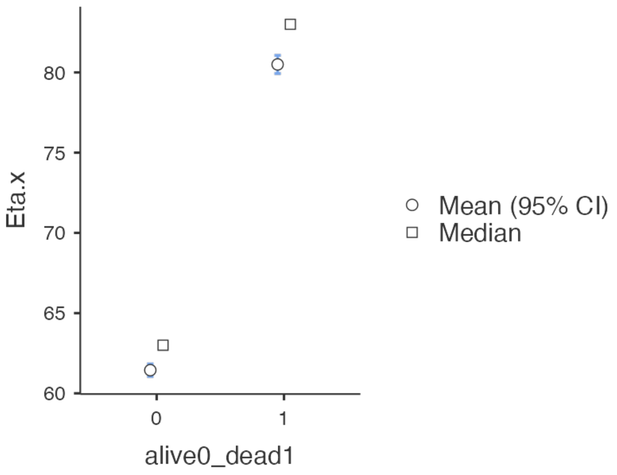

#### **Table S/3- Multivariable Survival Analysis** Benchmark BCDC, Gender, Age, Inward/Discharged

| Dependent: Surv | | all | HR (univariable) | HR (multivariable) |
|---|---|---|---|---|
| sex_0F_1M | 0 | 5284 (100.0) | - | - |
| | 1 | 5767 (100.0) | 1.16 (1.05-1.28, p=0.003) | 1.28 (1.16-1.42, p<0.001) |
| Age_0fav_1unf | 0 | 6692 (100.0) | - | - |
| | 1 | 4359 (100.0) | 6.72 (5.95-7.58, p<0.001) | 4.45 (3.92-5.05, p<0.001) |
| Dischrd0_Inward1 | 0 | 6488 (100.0) | - | - |
| | 1 | 4563 (100.0) | 4.02 (3.61-4.49, p<0.001) | 2.16 (1.93-2.42, p<0.001) |
| HB_ref | 0 | 5805 (100.0) | - | - |
| | 1 | 5246 (100.0) | 2.79 (2.51-3.11, p<0.001) | 1.51 (1.31-1.75, p<0.001) |
| RBC_ref | 0 | 5929 (100.0) | - | - |
| | 1 | 5122 (100.0) | 2.63 (2.36-2.92, p<0.001) | 1.05 (0.92-1.21, p=0.462) |
| MCV_ref | 0 | 8871 (100.0) | - | - |
| | 1 | 2180 (100.0) | 2.25 (2.03-2.50, p<0.001) | 1.25 (1.12-1.39, p<0.001) |
| RDW_ref | 0 | 8014 (100.0) | - | - |
| | 1 | 3037 (100.0) | 1.99 (1.80-2.20, p<0.001) | 1.57 (1.42-1.74, p<0.001) |
| WBC_ref | 0 | 5907 (100.0) | - | - |
| | 1 | 5144 (100.0) | 1.50 (1.36-1.66, p<0.001) | 1.10 (0.94-1.28, p=0.225) |
| Ne_ref | 0 | 6698 (100.0) | - | - |
| | 1 | 4353 (100.0) | 1.83 (1.66-2.02, p<0.001) | 1.28 (1.10-1.49, p=0.001) |
| Ly_ref | 0 | 9196 (100.0) | - | - |
| | 1 | 1855 (100.0) | 3.03 (2.73-3.35, p<0.001) | 1.45 (1.29-1.62, p<0.001) |
| Mo_ref | 0 | 7851 (100.0) | - | - |
| | 1 | 3200 (100.0) | 1.35 (1.22-1.50, p<0.001) | 1.01 (0.90-1.13, p=0.858) |
| Eo_ref | 0 | 6383 (100.0) | - | - |
| | 1 | 4668 (100.0) | 2.10 (1.90-2.31, p<0.001) | 1.33 (1.19-1.49, p<0.001) |
| Ba_ref | 0 | 10496 (100.0) | - | - |
| | 1 | 555 (100.0) | 2.85 (2.44-3.32, p<0.001) | 1.42 (1.20-1.68, p<0.001) |
| PLT_ref | 0 | 9422 (100.0) | - | - |
| | 1 | 1629 (100.0) | 2.42 (2.17-2.70, p<0.001) | 1.18 (1.01-1.39, p=0.042) |
| PCT_ref | 0 | 8282 (100.0) | - | - |
| | 1 | 2769 (100.0) | 2.07 (1.87-2.29, p<0.001) | 1.15 (1.00-1.34, p=0.055) |
| PDW_ref | 0 | 9166 (100.0) | - | - |
| | 1 | 1885 (100.0) | 1.58 (1.41-1.78, p<0.001) | 1.18 (1.05-1.32, p=0.007) |

**Model Metrics:** Number in dataframe = 11051, Number in model = 11051, Missing = 0, Number of events = 1597, Concordance = 0.821 (SE = 0.004), R-squared = 0.177( Max possible = 0.929), Likelihood ratio test = 2147.925 (df = 16, p = 0.000)

Abbreviation; value on reference interval (ref 0); value off interval (1)

Fig S/3

#### Hazards Regression Plot - Benchmark BCDC, Gender ,Age, Inw/Disch

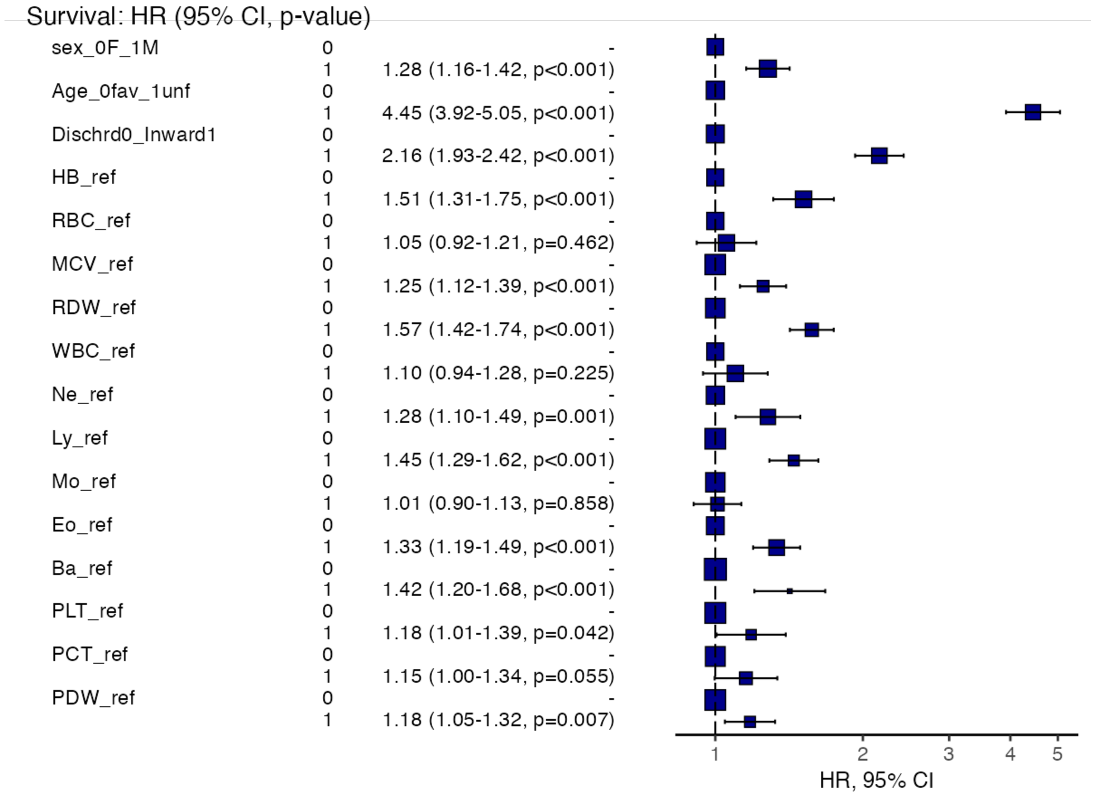

#### **Table S/4-Multivariable Survival Analysis** Tailored BCDC, Gender, Age, Inward/Discharged

| **Dependent: Surv** | | **all** | **HR (univariable)** | **HR (multivariable)** |
|---|---|---|---|---|
| sex_0F_1M | 0 | 5284 (100.0) | - | - |
| | 1 | 5767 (100.0) | 1.16 (1.05-1.28, p=0.003) | 1.26 (1.14-1.40, p<0.001) |
| Age_0fav_1unf | 0 | 6692 (100.0) | - | - |
| | 1 | 4359 (100.0) | 6.72 (5.95-7.58, p<0.001) | 3.78 (3.33-4.29, p<0.001) |
| Dischrd0_Inward1 | 0 | 6488 (100.0) | - | - |
| | 1 | 4563 (100.0) | 4.02 (3.61-4.49, p<0.001) | 1.92 (1.71-2.15, p<0.001) |
| Hb_DIC | 0 | 9296 (100.0) | - | - |
| | 1 | 1755 (100.0) | 3.59 (3.24-3.97, p<0.001) | 1.35 (1.16-1.57, p<0.001) |
| RBC_DIC | 0 | 9703 (100.0) | - | - |
| | 1 | 1348 (100.0) | 3.83 (3.45-4.26, p<0.001) | 1.07 (0.91-1.26, p=0.429) |
| MCV_DIC | 0 | 9736 (100.0) | - | - |
| | 1 | 1315 (100.0) | 2.76 (2.46-3.09, p<0.001) | 1.39 (1.22-1.57, p<0.001) |
| RDW_DIC | 0 | 7359 (100.0) | - | - |
| | 1 | 3692 (100.0) | 4.76 (4.29-5.29, p<0.001) | 2.22 (1.97-2.49, p<0.001) |
| WBC_0fav_1unf | 0 | 10266 (100.0) | - | - |
| | 1 | 785 (100.0) | 2.59 (2.25-2.97, p<0.001) | 1.12 (0.93-1.35, p=0.233) |
| NeDIC | 0 | 9117 (100.0) | - | - |
| | 1 | 1934 (100.0) | 2.11 (1.89-2.35, p<0.001) | 1.29 (1.11-1.49, p=0.001) |
| Ly0fav_1unf | 0 | 8067 (100.0) | - | - |
| | 1 | 2984 (100.0) | 3.04 (2.75-3.35, p<0.001) | 1.37 (1.23-1.53, p<0.001) |
| Mo_0fav_1unf | 0 | 10166 (100.0) | - | - |
| | 1 | 885 (100.0) | 2.36 (2.06-2.71, p<0.001) | 1.18 (1.02-1.36, p=0.030) |
| Eo0fav_1unf | 0 | 9303 (100.0) | - | - |
| | 1 | 1748 (100.0) | 3.06 (2.76-3.40, p<0.001) | 1.66 (1.47-1.87, p<0.001) |
| Ba_0fav_1unf | 0 | 10424 (100.0) | - | - |
| | 1 | 627 (100.0) | 2.74 (2.36-3.18, p<0.001) | 1.10 (0.93-1.30, p=0.272) |
| PLT_0fav_1unf | 0 | 10094 (100.0) | - | - |
| | 1 | 957 (100.0) | 2.87 (2.53-3.25, p<0.001) | 0.90 (0.73-1.11, p=0.311) |
| PCT_0fav_1unf | 0 | 9838 (100.0) | - | - |
| | 1 | 1213 (100.0) | 2.87 (2.56-3.22, p<0.001) | 1.53 (1.27-1.85, p<0.001) |
| PDW_0fav_1unf | 0 | 10078 (100.0) | - | - |
| | 1 | 973 (100.0) | 1.84 (1.60-2.12, p<0.001) | 1.20 (1.04-1.38, p=0.015) |

**Model Metrics:** Number in dataframe = 11051, Number in model = 11051, Missing = 0, Number of events = 1597, Concordance = 0.838 (SE = 0.004), R-squared = 0.195( Max possible = 0.929), Likelihood ratio test = 2401.428 (df = 16, p = 0.000)

Abbreviations: discretized value on favourable interval (0); discretized value on unfavourable interval (1); DIC = dichotomized by MSRS; 0fav_1unfav = discretized by OEHR

#### Fig S/4 Hazards Regression Plot - Tailored BCDC, Gender, Age, Inward/Discharged

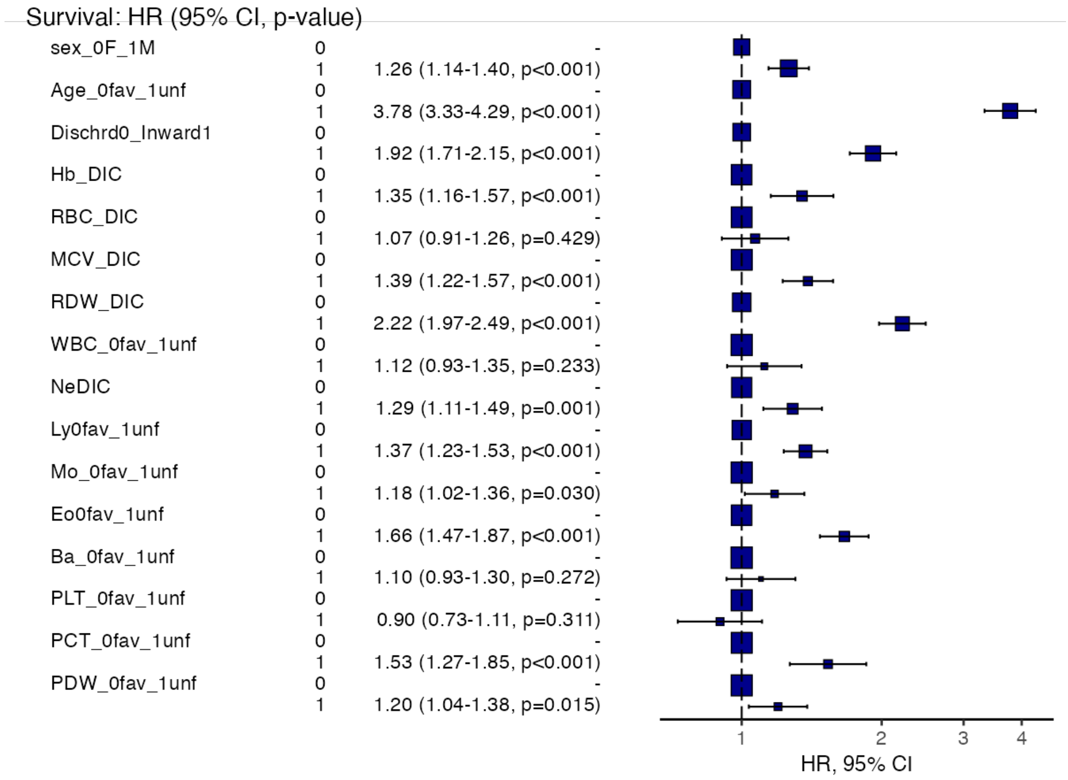

#### **Tab S/5 Multivariable Survival Analysis -** Benchmark BCDC, Gender, Age (without Inward/Discharged)

| **Dependent: Surv(mytime, myoutcome)** | | **all** | **HR (univariable)** | **HR (multivariable)** |
|---|---|---|---|---|
| sex_0F_1M | 0 | 5284 (100.0) | - | - |
| | 1 | 5768 (100.0) | 1.16 (1.05-1.28, p=0.003) | 1.37 (1.23-1.51, p<0.001) |
| Age_0fav_1unf | 0 | 6693 (100.0) | - | - |
| | 1 | 4359 (100.0) | 6.72 (5.95-7.58, p<0.001) | 5.06 (4.46-5.74, p<0.001) |
| HB_ref | 0 | 5806 (100.0) | - | - |
| | 1 | 5246 (100.0) | 2.79 (2.51-3.11, p<0.001) | 1.64 (1.46-1.84, p<0.001) |
| MCV_ref | 0 | 8872 (100.0) | - | - |
| | 1 | 2180 (100.0) | 2.26 (2.03-2.50, p<0.001) | 1.31 (1.18-1.46, p<0.001) |
| RDW_ref | 0 | 8015 (100.0) | - | - |
| | 1 | 3037 (100.0) | 1.99 (1.80-2.20, p<0.001) | 1.65 (1.49-1.83, p<0.001) |
| Ne_ref | 0 | 6698 (100.0) | - | - |
| | 1 | 4354 (100.0) | 1.83 (1.66-2.01, p<0.001) | 1.48 (1.34-1.64, p<0.001) |
| Ly_ref | 0 | 9197 (100.0) | - | - |
| | 1 | 1855 (100.0) | 3.03 (2.73-3.35, p<0.001) | 1.53 (1.36-1.72, p<0.001) |
| Eo_ref | 0 | 6384 (100.0) | - | - |
| | 1 | 4668 (100.0) | 2.10 (1.90-2.32, p<0.001) | 1.39 (1.25-1.56, p<0.001) |
| Ba_ref | 0 | 10497 (100.0) | - | - |
| | 1 | 555 (100.0) | 2.85 (2.44-3.32, p<0.001) | 1.56 (1.32-1.84, p<0.001) |
| PLT_ref | 0 | 9423 (100.0) | - | - |
| | 1 | 1629 (100.0) | 2.42 (2.17-2.70, p<0.001) | 1.35 (1.20-1.52, p<0.001) |
| PDW_ref | 0 | 9167 (100.0) | - | - |
| | 1 | 1885 (100.0) | 1.58 (1.41-1.78, p<0.001) | 1.21 (1.07-1.36, p=0.002) |

**Model Metrics:** Number in dataframe = 11052, Number in model = 11052, Missing = 0, Number of events = 1597, Concordance = 0.806 (SE = 0.005), R-squared = 0.162( Max possible = 0.929), Likelihood ratio test = 1949.794 (df = 11, p = 0.000)

Abbreviation; value on reference interval (ref 0); value off interval (1)

#### Fig S/5

#### Hazards Regression Plot - Benchmark BCDC, Gender, Age (without Inward/Discharged)

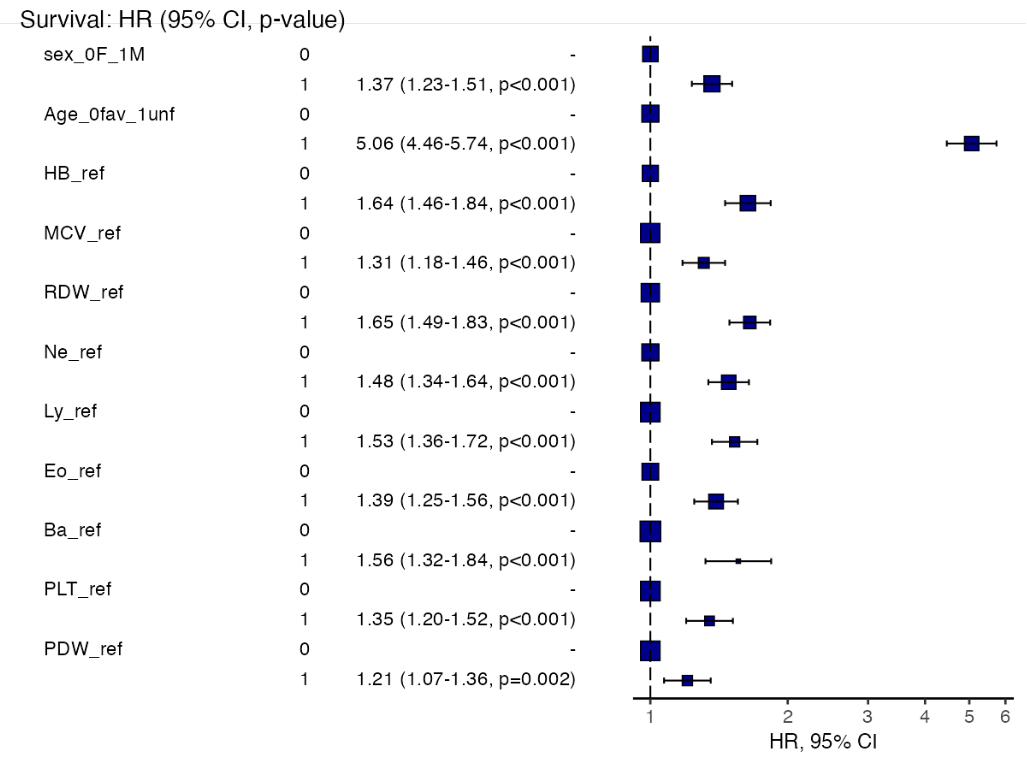

#### Tab S/6

#### **Multivariable Survival Analysis -** Tailored BCDC, Gender, Age (without Inward/Discharged)

| Dependent: Surv(mytime, myoutcome) | | all | HR (univariable) | HR (multivariable) |
|---|---|---|---|---|
| sex_0F_1M | 0 | 5284 (100.0) | - | - |
| | 1 | 5768 (100.0) | 1.16 (1.05-1.28, p=0.003) | 1.31 (1.19-1.45, p<0.001) |
| Age_0fav_1unf | 0 | 6693 (100.0) | - | - |
| | 1 | 4359 (100.0) | 6.72 (5.95-7.58, p<0.001) | 4.02 (3.54-4.56, p<0.001) |
| Hb_DIC | 0 | 9297 (100.0) | - | - |
| | 1 | 1755 (100.0) | 3.59 (3.24-3.97, p<0.001) | 1.50 (1.34-1.67, p<0.001) |
| MCV_DIC | 0 | 9737 (100.0) | - | - |
| | 1 | 1315 (100.0) | 2.76 (2.46-3.10, p<0.001) | 1.46 (1.30-1.64, p<0.001) |
| RDW_DIC | 0 | 7360 (100.0) | - | - |
| | 1 | 3692 (100.0) | 4.76 (4.29-5.29, p<0.001) | 2.39 (2.13-2.68, p<0.001) |
| NeDIC | 0 | 9118 (100.0) | - | - |
| | 1 | 1934 (100.0) | 2.11 (1.89-2.35, p<0.001) | 1.48 (1.33-1.66, p<0.001) |
| Ly0fav_1unf | 0 | 8068 (100.0) | - | - |
| | 1 | 2984 (100.0) | 3.04 (2.75-3.35, p<0.001) | 1.43 (1.28-1.59, p<0.001) |
| Mo_0fav_1unf | 0 | 10167 (100.0) | - | - |
| | 1 | 885 (100.0) | 2.36 (2.06-2.71, p<0.001) | 1.24 (1.07-1.43, p=0.003) |
| Eo0fav_1unf | 0 | 9304 (100.0) | - | - |
| | 1 | 1748 (100.0) | 3.06 (2.76-3.40, p<0.001) | 1.76 (1.57-1.98, p<0.001) |
| PCT_0fav_1unf | 0 | 9839 (100.0) | - | - |
| | 1 | 1213 (100.0) | 2.87 (2.56-3.22, p<0.001) | 1.47 (1.30-1.65, p<0.001) |
| PDW_0fav_1unf | 0 | 10079 (100.0) | - | - |
| | 1 | 973 (100.0) | 1.84 (1.60-2.12, p<0.001) | 1.23 (1.07-1.42, p=0.004) |

**Model Metrics:** Number in dataframe = 11052, Number in model = 11052, Missing = 0, Number of events = 1597, Concordance = 0.828 (SE = 0.004), R-squared = 0.185( Max possible = 0.929), Likelihood ratio test = 2261.731 (df = 11, p = 0.000)

Abbreviations: discretized value on favourable interval (0); discretized value on unfavourable interval (1); DIC = dichotomized by MSRS; 0fav_1unfav = discretized by OEHR

#### Fig S/6 - Hazards Regression Plot - Tailored BCDC, Gender, Age (without Inward/Discharged)

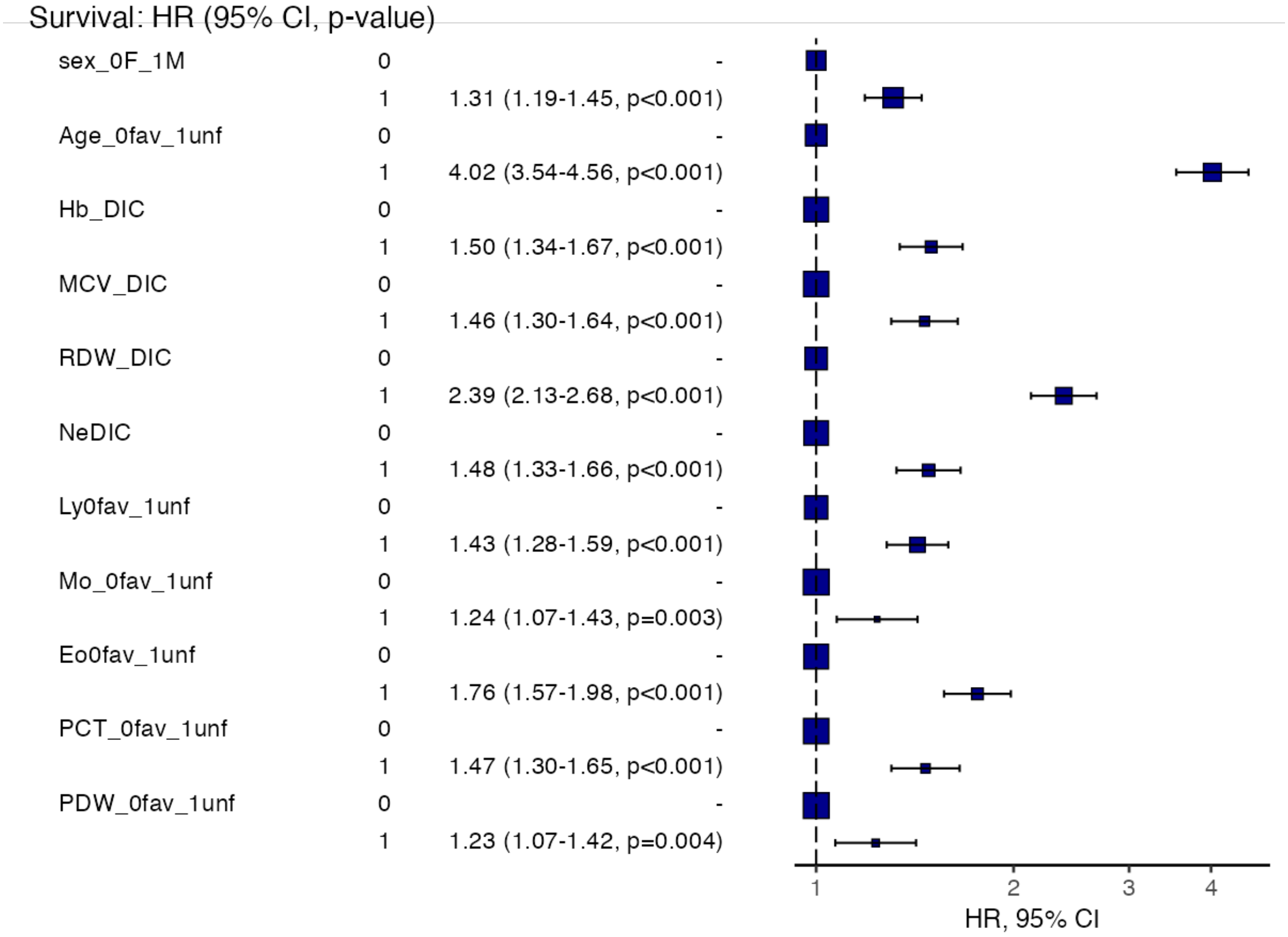

#### Tab S/7 - Score sum (count of unfavourable factors by quintiles) for each discretization method

Descriptives

| | Benchm_CBDC_sum | Tailored_CBDC_sum |
|---|---|---|
| N | 11052 | 11052 |
| Missing | 0 | 0 |
| Mean | 2.30 | 1.49 |
| Median | 2.00 | 1.00 |
| Standard deviation | 1.61 | 1.52 |
| Minimum | 0.00 | 0.00 |
| Maximum | 9.00 | 8.00 |
| 20th percentile | 1.00 | 0.00 |
| 40th percentile | 2.00 | 1.00 |
| 60th percentile | 3.00 | 2.00 |
| 80th percentile | 4.00 | 3.00 |

#### Fig S/7 Plots

**Fig. S/7a Benchm_CBDC_sum**

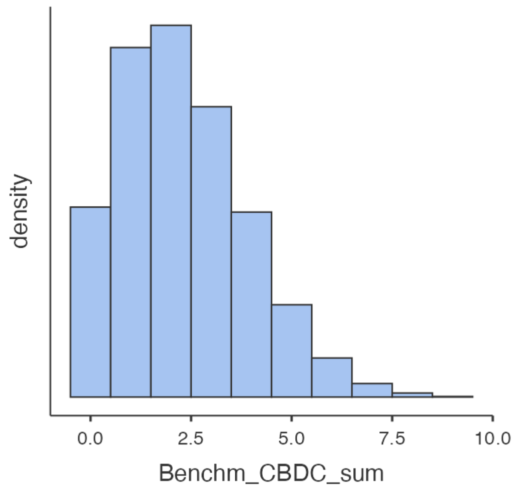


**Fig.S/7b Tailored_CBDC_sum**

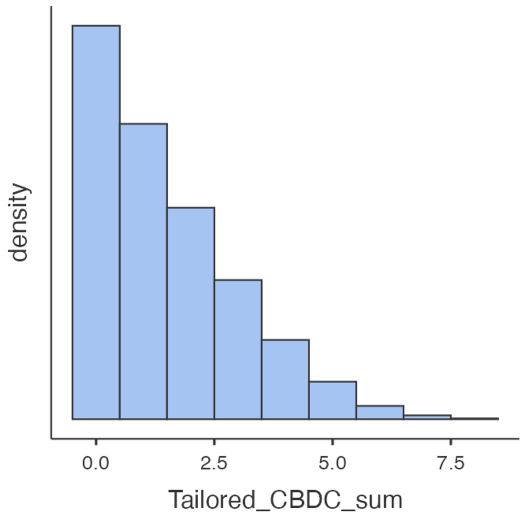

#### Tab S/8 Descriptive statistics of Score sum (count of unfavourable factors by quintiles) for each discretization method by Alive/Dead status

Descriptives

| | alive0_dead1 | Bench_CBDC_sum_Quintiles | Tail_CBDC_sum_Quintiles |
|---|---|---|---|
| N | 0 | 9455 | 9455 |
| | 1 | 1597 | 1597 |
| Missing | 0 | 0 | 0 |
| | 1 | 0 | 0 |
| Mean | 0 | 2.20 | 2.21 |
| | 1 | 3.38 | 3.70 |
| Median | 0 | 2 | 2 |
| | 1 | 3 | 4 |
| Standard deviation | 0 | 1.24 | 1.25 |
| | 1 | 1.35 | 1.27 |
| Minimum | 0 | 1 | 1 |
| | 1 | 1 | 1 |
| Maximum | 0 | 5 | 5 |
| | 1 | 5 | 5 |

#### Tab S/8a
#### Frequencies of Score sum (count of unfavourable factors by quintiles) for Benchmark discretization method

Frequencies of Bench_CBDC_sum_Quintiles

| | alive0_dead1 | |
|---|---|---|
| **Bench_CBDC_sum_Quintiles** | **0** | **1** |
| 1 | 3703 | 179 |
| 2 | 2393 | 282 |
| 3 | 1747 | 342 |
| 4 | 992 | 339 |
| 5 | 620 | 455 |

#### Tab S/8b Frequencies of Score sum (count of unfavourable factors by quintiles) for Tailored discretization method

Frequencies of Tail_CBDC_sum_Quintiles

| | alive0_dead1 | |
|---|---|---|
| **Tail_CBDC_sum_Quintiles** | **0** | **1** |
| 1 | 3598 | 106 |
| 2 | 2567 | 213 |
| 3 | 1672 | 319 |
| 4 | 938 | 372 |
| 5 | 680 | 587 |

#### Fig S/8 Plots

Fig S/8a Count of Alive or Dead pts by Benchmark Score Sum (count of off-interval BCDC values) quintiles

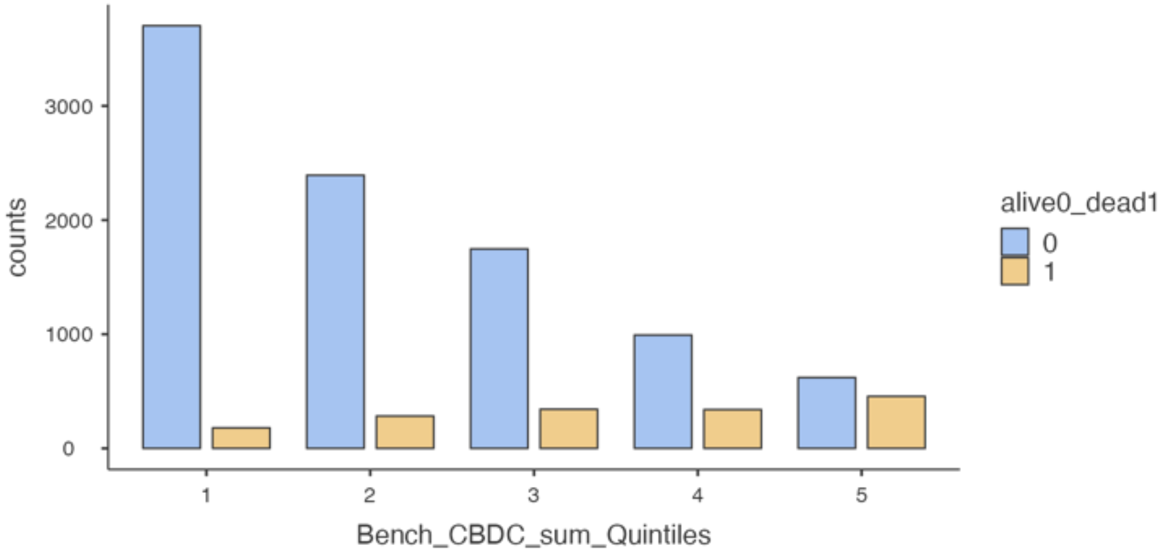

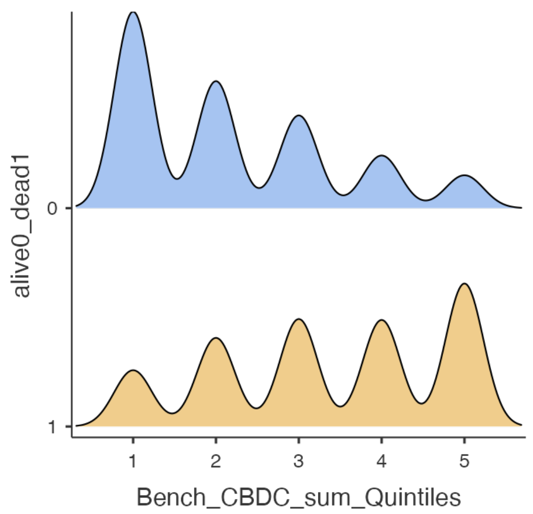


Fig. S/8b Count of Alive or Dead pts by Tailored Score Sum (count of unfavourable BCDC values) quintiles

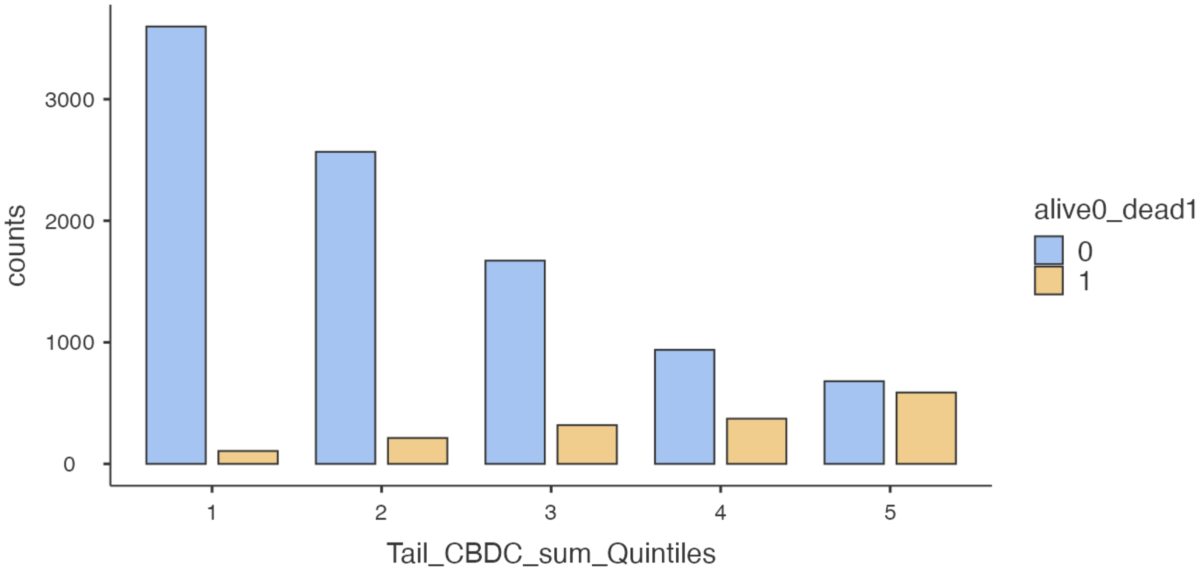


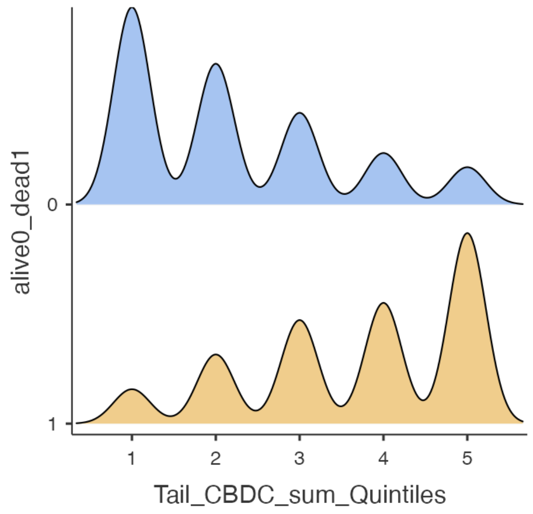

Tab S/9

#### Tab S/9 Survival Analysis -Benchmark Score Sum stratified by risk group

Events Summary

| | N | Censored | Observed Events | Expected Events |
|---|---|---|---|---|
| 1 | 3882 | 3703 | 179 | 598 |
| 2 | 2675 | 2393 | 282 | 397 |
| 3 | 2089 | 1747 | 342 | 296 |
| 4 | 1331 | 992 | 339 | 179 |
| 5 | 1075 | 620 | 455 | 128 |

S/9a Analysis of the differences

| Test | $\chi^2$ | df | p |
|---|---|---|---|
| Log-rank | 1319 | 4 | < .001 |

S/9b Pairwise Comparisons

| | | Test | ν | SE | z | p |
|---|---|---|---|---|---|---|
| 1 | 2 | Log-rank | 99.1 | 10.50 | 9.441 | < .001 |
| 1 | 3 | Log-rank | 170.2 | 10.72 | 15.870 | < .001 |
| 1 | 4 | Log-rank | 219.9 | 9.57 | 22.990 | < .001 |
| 1 | 5 | Log-rank | 342.7 | 9.59 | 35.747 | < .001 |
| 2 | 3 | Log-rank | 75.7 | 12.35 | 6.129 | < .001 |
| 2 | 4 | Log-rank | 146.1 | 11.52 | 12.688 | < .001 |
| 2 | 5 | Log-rank | 275.3 | 11.62 | 23.679 | < .001 |
| 3 | 4 | Log-rank | 81.8 | 12.63 | 6.477 | < .001 |
| 3 | 5 | Log-rank | 213.3 | 12.94 | 16.483 | < .001 |
| 4 | 5 | Log-rank | 123.7 | 13.85 | 8.931 | < .001 |

Note. P-values are Bonferroni-corrected.

#### Fig S/9a Survival Curve Benchmark Score Sum stratified by risk group

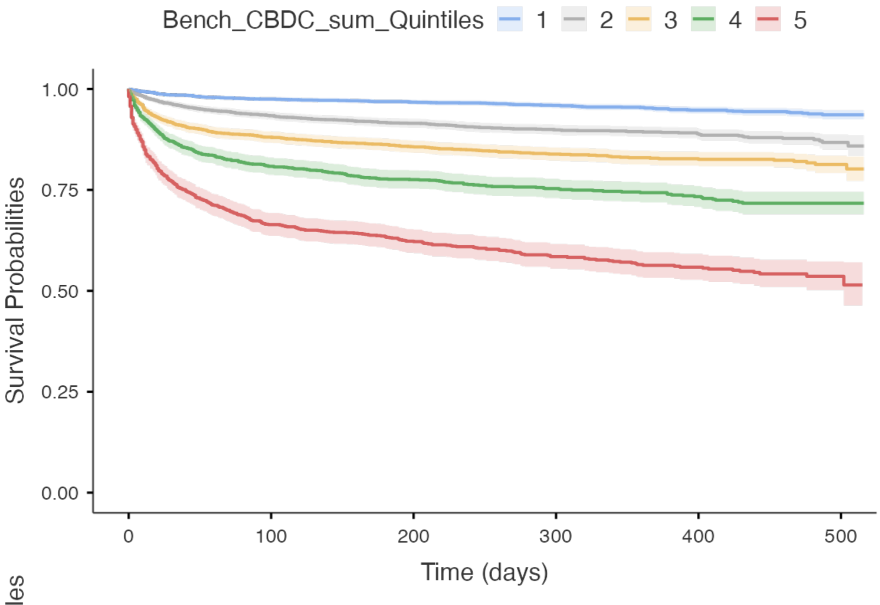


Number at risk

| Bench_CBDC_sum_Quintiles | 0 | 100 | 200 | 300 | 400 | 500 |
|---|---|---|---|---|---|---|
| 1 | 3882 | 3779 | 3345 | 2304 | 1195 | 271 |
| 2 | 2675 | 2496 | 2117 | 1422 | 752 | 142 |
| 3 | 2089 | 1837 | 1560 | 1018 | 555 | 93 |
| 4 | 1331 | 1074 | 903 | 584 | 344 | 58 |
| 5 | 1075 | 713 | 562 | 378 | 233 | 34 |

Time (days)

#### Fig S/9b Cumulative Hazard Function Benchmark Score Sum stratified by risk group

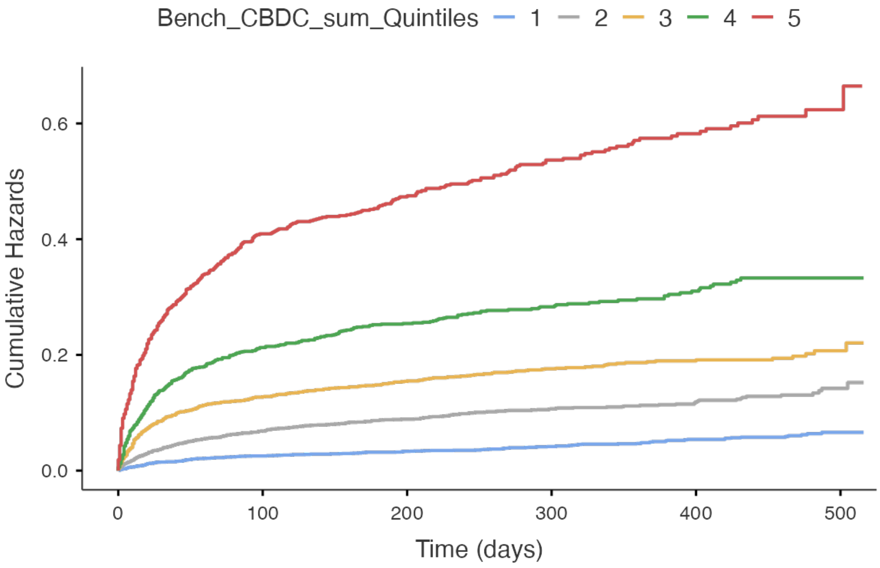

### Tab S/10

#### Tab S/10 Survival Analysis -Tailored Score Sum stratified by risk group

Events Summary

| | N | Censored | Observed Events | Expected Events |
|---|---|---|---|---|
| 1 | 3704 | 3598 | 106 | 577 |
| 2 | 2780 | 2567 | 213 | 418 |
| 3 | 1991 | 1672 | 319 | 286 |
| 4 | 1310 | 938 | 372 | 172 |
| 5 | 1267 | 680 | 587 | 144 |

### S/10a

Analysis of the differences

| Test | $\chi^2$ | df | p |
|---|---|---|---|
| Log-rank | 2091 | 4 | < .001 |

### S/10b

Pairwise Comparisons

| | | Test | ν | SE | z | p |
|---|---|---|---|---|---|---|
| 1 | 2 | Log-rank | 80.0 | 8.80 | 9.084 | < .001 |
| 1 | 3 | Log-rank | 179.5 | 9.67 | 18.563 | < .001 |
| 1 | 4 | Log-rank | 263.0 | 9.16 | 28.717 | < .001 |
| 1 | 5 | Log-rank | 446.1 | 10.55 | 42.279 | < .001 |
| 2 | 3 | Log-rank | 103.5 | 11.32 | 9.143 | < .001 |
| 2 | 4 | Log-rank | 201.9 | 10.97 | 18.414 | < .001 |
| 2 | 5 | Log-rank | 379.7 | 12.34 | 30.764 | < .001 |
| 3 | 4 | Log-rank | 113.0 | 12.70 | 8.894 | < .001 |
| 3 | 5 | Log-rank | 281.9 | 14.17 | 19.894 | < .001 |
| 4 | 5 | Log-rank | 148.3 | 15.37 | 9.649 | < .001 |

#### Fig.S/10a

##### Survival Curve Tailored Score Sum stratified by risk group

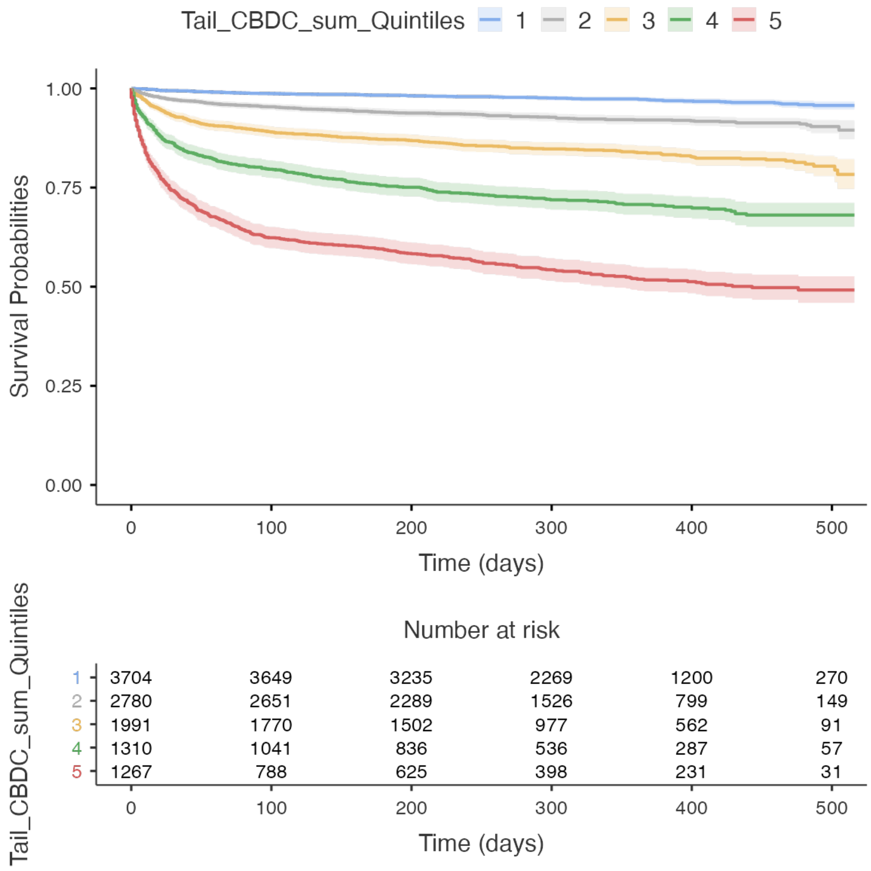


##### Cumulative Hazard Function - Tailored Score Sum stratified by risk group

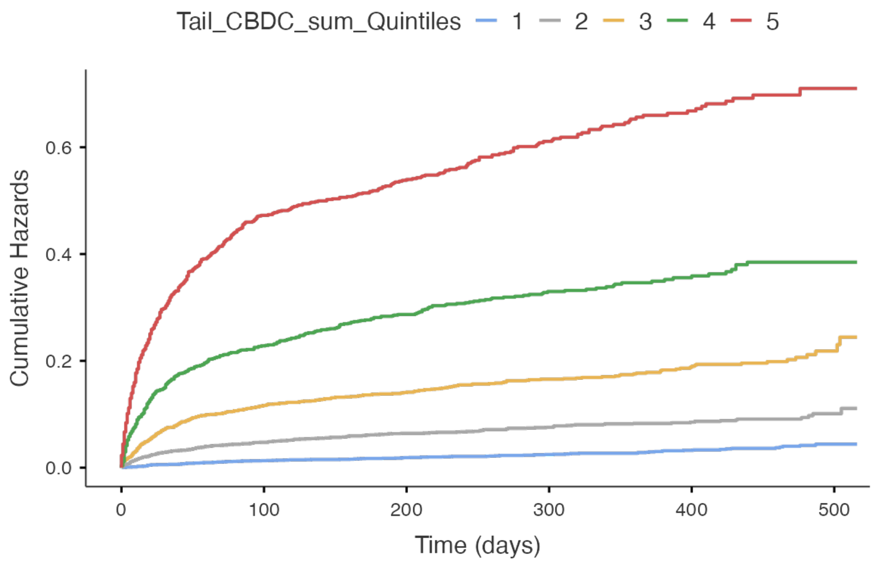

### Table S/11

#### TestROC - Benchmark BCDC, Tailored BCDC

Procedure Notes

The TestROC optimal cutpoint analysis has been completed using the following specifications:

Measure Variable(s): Benchm_CBDC_sum, Tailored_CBDC_sum

Class Variable: deadTRUE

Positive Class: TRUE

Method: maximize_metric

All Observed Cutpoints: TRUE

Metric: roc01

Direction (relative to cutpoint): >=

Tie Breakers: c

Metric Tolerance: 0.05

##### Results Table

Scale: Benchm_CBDC_sum

| Cutpoint | Sensitivity (%) | Specificity (%) | PPV (%) | NPV (%) | Youden's index | AUC | Metric Score |
|---|---|---|---|---|---|---|---|
| 0 | 100% | 0% | 14.45% | NaN% | 0.00000 | 0.739 | 1.000 |
| 1 | 98% | 14.12% | 16.16% | 97.66% | 0.12116 | 0.739 | 0.859 |
| 2 | 88.79% | 39.16% | 19.78% | 95.39% | 0.27956 | 0.739 | 0.619 |
| 3 | 71.13% | 64.47% | 25.27% | 92.97% | 0.35607 | 0.739 | 0.458 |
| 4 | 49.72% | 82.95% | 33% | 90.71% | 0.32669 | 0.739 | 0.531 |
| 5 | 28.49% | 93.44% | 42.33% | 88.55% | 0.21934 | 0.739 | 0.718 |
| 6 | 12.52% | 97.77% | 48.66% | 86.87% | 0.10292 | 0.739 | 0.875 |
| 7 | 4.13% | 99.32% | 50.77% | 85.98% | 0.03456 | 0.739 | 0.959 |
| 8 | 1.19% | 99.85% | 57.58% | 85.68% | 0.01042 | 0.739 | 0.988 |
| 9 | 0.13% | 99.98% | 50% | 85.56% | 0.00104 | 0.739 | 0.999 |

Scale: Tailored_CBDC_sum

| Cutpoint | Sensitivity (%) | Specificity (%) | PPV (%) | NPV (%) | Youden's index | AUC | Metric Score |
|---|---|---|---|---|---|---|---|
| 0 | 100% | 0% | 14.45% | NaN% | 0.00000 | 0.790 | 1.000 |
| 1 | 93.36% | 38.05% | 20.29% | 97.14% | 0.31416 | 0.790 | 0.623 |
| 2 | 80.03% | 65.2% | 27.98% | 95.08% | 0.45229 | 0.790 | 0.401 |
| 3 | 60.05% | 82.89% | 37.21% | 92.47% | 0.42937 | 0.790 | 0.435 |
| 4 | 36.76% | 92.81% | 46.33% | 89.68% | 0.29564 | 0.790 | 0.637 |
| 5 | 17.09% | 97.38% | 52.4% | 87.43% | 0.14472 | 0.790 | 0.829 |
| 6 | 5.32% | 99.12% | 50.6% | 86.11% | 0.04445 | 0.790 | 0.947 |
| 7 | 1.57% | 99.81% | 58.14% | 85.72% | 0.01375 | 0.790 | 0.984 |
| 8 | 0.31% | 99.98% | 71.43% | 85.59% | 0.00292 | 0.790 | 0.997 |

#### DeLong Test of Difference between AUCs (1=Benchmark; 2=Tailored)

```
Estimated AUC's:
    AUC SD(Hanley) P(H0: AUC=0.5) SD(DeLong) P(H0: AUC=0.5)
1 0.739      0.007          0.000      0.007          0.000
2 0.790      0.007          0.000      0.006          0.000

 Pairwise comparisons:
        AUC Difference CI(lower) CI(upper) P.Value Correlation
1 vs. 2         -0.051    -0.060    -0.042   0.000       0.728

 Overall test: p-value = <2e-16
```

#### Fig S/11

##### ROC Curves

ROC Curve: Combined

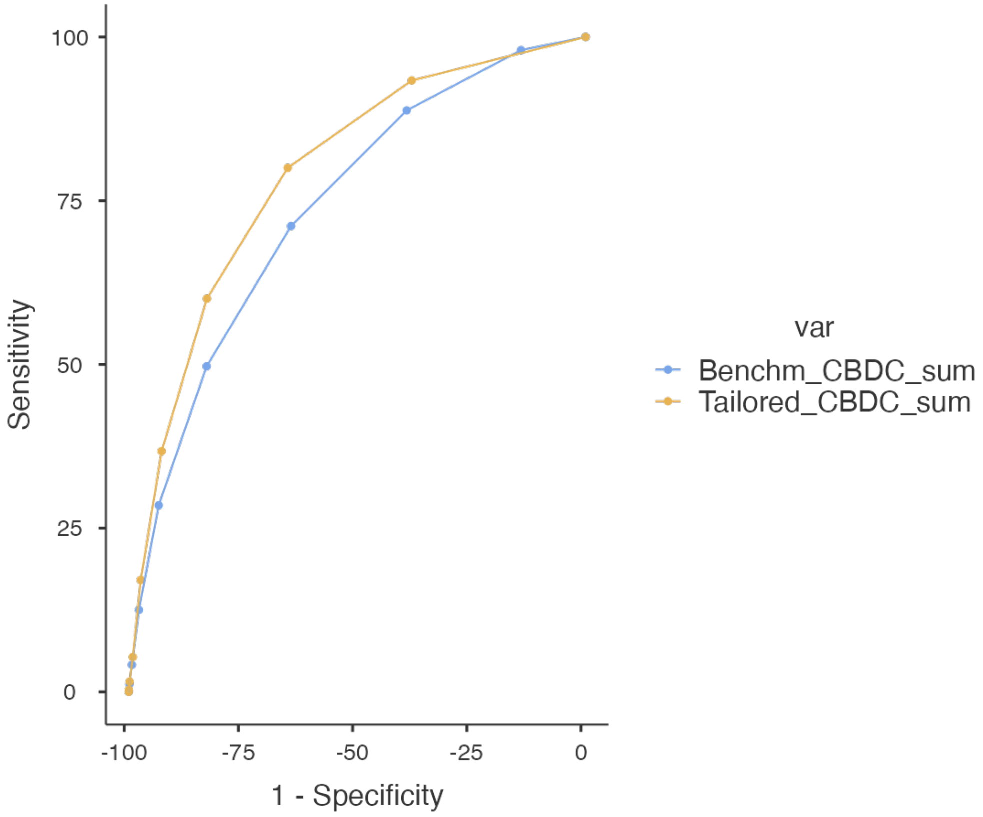


Abbreviations: Positive predictive value (PPV), negative predictive value (NPV), area under the curve (AUC), standard deviation (SD), blood cell differential count (BCDC), Benchmark method (Bench), Tailored method (Tailrd)

#### Table S/12

TestROC  Benchmark BCDC+age+sex, Tailored BCDC+age+sex, Age+Sex

##### curve 1 BCDC Benchmark age sex
##### curve 2 BCDC Tailored age sex
##### curve 3 Age Sex

Procedure Notes

The TestROC optimal cutpoint analysis has been completed using the following specifications:

Measure Variable(s): Benchm_SxAge_sum, Tailrd_SxAge_sum, SEX_AGE_sum

Class Variable: deadTRUE

Positive Class: TRUE

Method: maximize_metric

All Observed Cutpoints: TRUE

Metric: roc01

Direction (relative to cutpoint): >=

Tie Breakers: c

Metric Tolerance: 0.05

##### Results Table

Scale: Benchm_SxAge_sum

| Cutpoint | Sensitivity (%) | Specificity (%) | PPV (%) | NPV (%) | Youden's index | AUC | Metric Score |
|---|---|---|---|---|---|---|---|
| 0 | 100% | 0% | 14.45% | NaN% | 0.00000 | 0.787 | 1.000 |
| 1 | 99.75% | 3.82% | 14.91% | 98.9% | 0.03568 | 0.787 | 0.962 |
| 2 | 98.5% | 21.37% | 17.46% | 98.83% | 0.19872 | 0.787 | 0.786 |
| 3 | 91.86% | 44.44% | 21.83% | 97% | 0.36302 | 0.787 | 0.562 |
| 4 | 77.46% | 66.36% | 28% | 94.57% | 0.43814 | 0.787 | 0.405 |
| 5 | 56.86% | 82.54% | 35.48% | 91.89% | 0.39395 | 0.787 | 0.465 |
| 6 | 35.88% | 92.23% | 43.81% | 89.49% | 0.28106 | 0.787 | 0.646 |
| 7 | 17.85% | 97.11% | 51.08% | 87.5% | 0.14959 | 0.787 | 0.822 |
| 8 | 7.26% | 99.1% | 57.71% | 86.35% | 0.06365 | 0.787 | 0.927 |

Scale: Benchm_SxAge_sum

| Cutpoint | Sensitivity (%) | Specificity (%) | PPV (%) | NPV (%) | Youden's index | AUC | Metric Score |
|---|---|---|---|---|---|---|---|
| 9 | 2.57% | 99.77% | 65.08% | 85.84% | 0.02335 | 0.787 | 0.974 |
| 10 | 0.63% | 99.97% | 76.92% | 85.62% | 0.00594 | 0.787 | 0.994 |
| 11 | 0.06% | 99.99% | 50% | 85.56% | 5.20e-4 | 0.787 | 0.999 |

Scale: Tailrd_SxAge_sum

| Cutpoint | Sensitivity (%) | Specificity (%) | PPV (%) | NPV (%) | Youden's index | AUC | Metric Score |
|---|---|---|---|---|---|---|---|
| 0 | 100% | 0% | 14.45% | NaN% | 0.00000 | 0.823 | 1.000 |
| 1 | 99.12% | 14.19% | 16.33% | 98.97% | 0.13317 | 0.823 | 0.858 |
| 2 | 95.12% | 43.23% | 22.06% | 98.13% | 0.38342 | 0.823 | 0.570 |
| 3 | 84.78% | 65.59% | 29.39% | 96.23% | 0.50379 | 0.823 | 0.376 |
| 4 | 66.94% | 81.69% | 38.18% | 93.6% | 0.48630 | 0.823 | 0.378 |
| 5 | 44.71% | 91.35% | 46.61% | 90.72% | 0.36057 | 0.823 | 0.560 |
| 6 | 24.73% | 96.44% | 53.96% | 88.35% | 0.21170 | 0.823 | 0.754 |
| 7 | 9.39% | 98.79% | 56.82% | 86.59% | 0.08187 | 0.823 | 0.906 |
| 8 | 3.32% | 99.64% | 60.92% | 85.92% | 0.02959 | 0.823 | 0.967 |
| 9 | 0.88% | 99.94% | 70% | 85.65% | 0.00813 | 0.823 | 0.991 |
| 10 | 0.06% | 100% | 100% | 85.56% | 6.26e-4 | 0.823 | 0.999 |

Scale: SEX_AGE_sum

| Cutpoint | Sensitivity (%) | Specificity (%) | PPV (%) | NPV (%) | Youden's index | AUC | Metric Score |
|---|---|---|---|---|---|---|---|
| 0 | 100% | 0% | 14.45% | NaN% | 0.000 | 0.691 | 1.000 |
| 1 | 92.36% | 31.11% | 18.46% | 96.02% | 0.235 | 0.691 | 0.693 |
| 2 | 42.2% | 84.52% | 31.52% | 89.65% | 0.267 | 0.691 | 0.598 |

#### **DeLong Test of Difference between AUCs** (curve 1=BCDC Benchmark age sex; curve 2 =BCDC Tailored age sex; curve 3= Age Sex)

```
Estimated AUC's:
    AUC  SD(Hanley)  P(H0: AUC=0.5)  SD(DeLong)  P(H0: AUC=0.5)
1  0.787      0.007           0.000       0.006           0.000
2  0.823      0.007           0.000       0.005           0.000
3  0.691      0.008           0.000       0.006           0.000

 Pairwise comparisons:
```

|  | AUC Difference | CI(lower) | CI(upper) | P.Value | Correlation |
|---|---|---|---|---|---|
| 1 vs. 2 | -0.036 | -0.043 | -0.029 | 0.000 | 0.766 |
| 1 vs. 3 | 0.096 | 0.082 | 0.109 | 0.000 | 0.329 |
| 2 vs. 3 | 0.132 | 0.119 | 0.144 | 0.000 | 0.383 |

Overall test:
p-value =

#### Fig. S/12 ROC Curves

**ROC Curve: Combined**

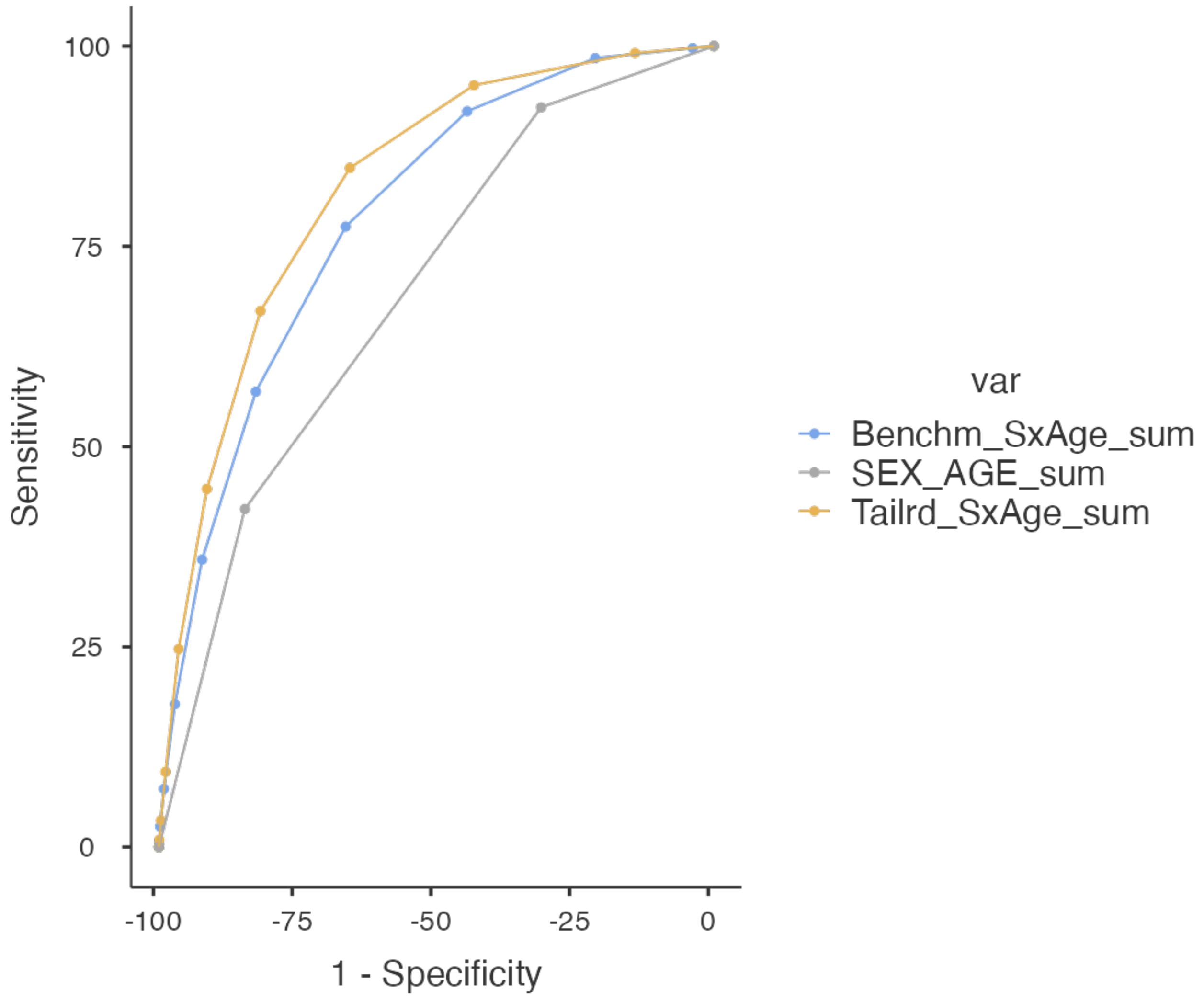

### Section TECHNIQUES OF BLOOD CELL DIFFERENTIAL COUNT DISCRETIZATION

#### Penalized B-splines (psplines) plotting

#### OEHR (optimal-equal hazard ratio method) discretization for U-shaped distribution variables

```
# EMOGLOBINA Waves120

#Call:
#  coxph(formula = Surv(ddInwFUp, deadTRUE, type = "right") ~ pspline(EMOGLOBINA.x,
#                                        df = 3, caic = TRUE), data = cutpoints99_Wave1_2_0_CSV)
#
#coef se(coef)     se2    Chisq  DF     p
#pspline(EMOGLOBINA.x, df   -0.2616   0.0105   0.0105 620.1145 1.0 <2e-16
#       pspline(EMOGLOBINA.x, df                        87.5145 2.1 <2e-16
#
#            Iterations: 5 outer, 17 Newton-Raphson
#            Theta= 0.956
#            Degrees of freedom for terms= 3.1
#            Likelihood ratio test=679  on 3.1 df, p=<2e-16
#            n= 11052, number of events= 1597
```

```
termplot(HbWav120Cox, term=1, se=TRUE, col.term=1, col.se=1)
```

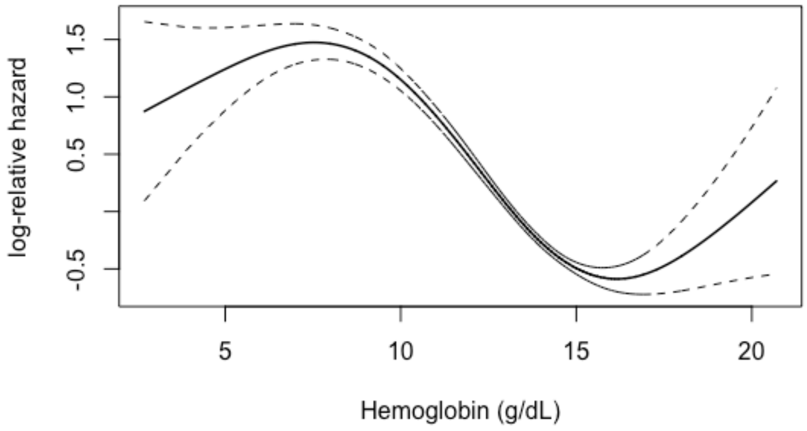

#GLOBULI ROSSI Waves120

#Call:
#coxph(formula = Surv(ddInwFUp, deadTRUE, type = "right") ~ pspline(GLOBULI_ROSSI.x,
### df = 3, caic = TRUE), data = cutpoints99_Wave1_2_0_CSV)
#
#coef se(coef) se2 Chisq DF p
#pspline(GLOBULI_ROSSI.x, -0.8008 0.0314 0.0314 651.3075 1.00 < 2e-16
### pspline(GLOBULI_ROSSI.x, 64.5139 2.05 1.1e-14
#
### Iterations: 5 outer, 17 Newton-Raphson
### Theta= 0.925
### Degrees of freedom for terms= 3.1
### Likelihood ratio test=660 on 3.05 df, p=<2e-16
### n= 11051, number of events= 1597
### (1 observation deleted due to missingness)

termplot(RBCWav120Cox, term=1, se=TRUE, col.term=1, col.se=1)

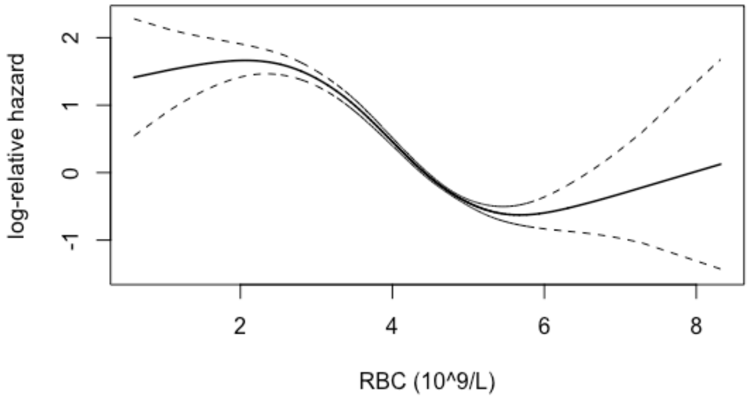


### RDW waves 120

#Call:
### coxph(formula = Surv(ddInwFUp, deadTRUE, type = "right") ~ pspline(RDW.x,
### df = 3, caic = TRUE), data = cutpoints99_Wave1_2_0_CSV)
#
#coef se(coef) se2 Chisq DF p
#pspline(RDW.x, df = 3, ca 0.2941 0.0112 0.0112 689.2405 1.00 <2e-16
### pspline(RDW.x, df = 3, ca 356.3978 2.04 <2e-16
#
### Iterations: 6 outer, 18 Newton-Raphson
### Theta= 0.949
### Degrees of freedom for terms= 3

#               Likelihood ratio test=1165  on 3.04 df, p=<2e-16
#               n= 11052, number of events= 1597

termplot(RDWWav120Cox, term=1, se=TRUE, col.term=1, col.se=1)

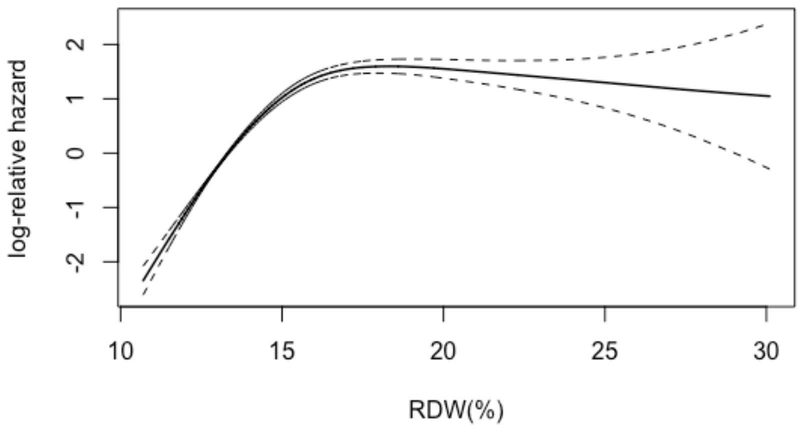


### MCV Waves120

#Call:
###  coxph(formula = Surv(ddInwFUp, deadTRUE, type = "right") ~ pspline(MCV.x,
#                                          df = 3, caic = TRUE), data = cutpoints99_Wave1_2_0_CSV)
#
#coef se(coef)     se2    Chisq   DF      p
#pspline(MCV.x, df = 3, ca 4.66e-02 3.24e-03 3.24e-03 2.07e+02 1.00 <2e-16
#       pspline(MCV.x, df = 3, ca                        9.58e+01 2.07 <2e-16
#
#               Iterations: 5 outer, 19 Newton-Raphson
#               Theta= 0.934
#               Degrees of freedom for terms= 3.1
#               Likelihood ratio test=288  on 3.07 df, p=<2e-16
#               n= 11052, number of events= 1597

termplot(MCVWav120Cox, term=1, se=TRUE, col.term=1, col.se=1)

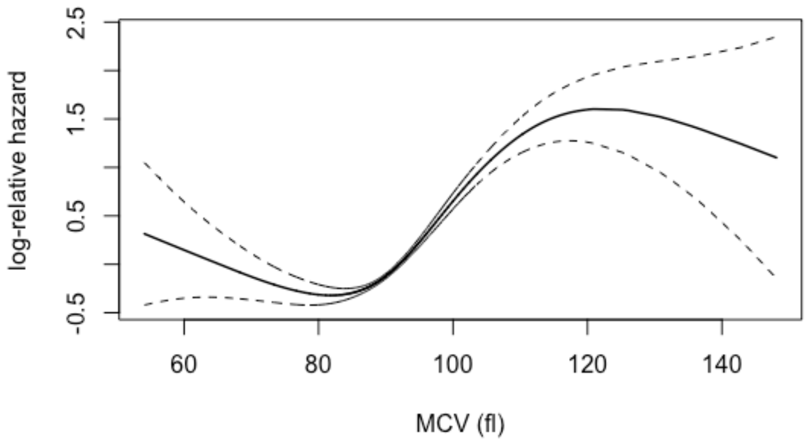


```
# LEUCOCITI Waves 120

#Call:
#  coxph(formula = Surv(ddInwFUp, deadTRUE, type = "right") ~ pspline(LEUCOCITI.x,
#                                                    df = 3, caic = TRUE), data = cutpoints99_Wave1_2_0_CSV)
#
#coef se(coef)     se2    Chisq   DF      p
#pspline(LEUCOCITI.x, df = 6.42e-02 4.97e-03 4.97e-03 1.67e+02 1.00 <2e-16
#       pspline(LEUCOCITI.x, df =                         5.55e+01 2.07  1e-12
#
#             Iterations: 5 outer, 18 Newton-Raphson
#             Theta= 0.94
#             Degrees of freedom for terms= 3.1
#             Likelihood ratio test=184  on 3.07 df, p=<2e-16
#             n= 11052, number of events= 1597

termplot(WBCWaves120Cox, term=1, se=TRUE, col.term=1, col.se=1)
```

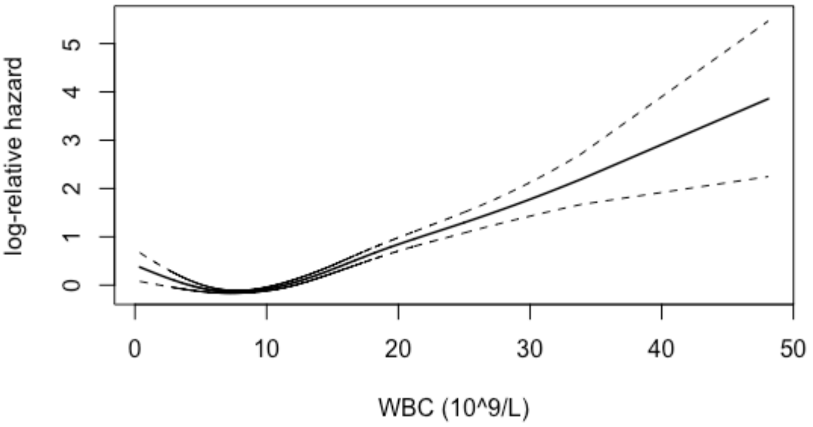


```
# Ne Waves120

#Call:
#  coxph(formula = Surv(ddInwFUp, deadTRUE, type = "right") ~ pspline(neutrofili.x,
#      df = 3, caic = TRUE), data = cutpoints99_Wave1_2_0_CSV)
#
#                          coef se(coef)      se2    Chisq   DF      p
#pspline(neutrofili.x, df  8.74e-02 5.38e-03 5.38e-03 2.64e+02 1.00 <2e-16
#pspline(neutrofili.x, df                       2.35e+00 2.07   0.32


#Iterations: 6 outer, 18 Newton-Raphson
#Theta= 0.975
#Degrees of freedom for terms= 3.1
#Likelihood ratio test=228  on 3.07 df, p=<2e-16
#n= 11052, number of events= 1597

termplot(NeWav120Cox, term=1, se=TRUE, col.term=1, col.se=1)
```

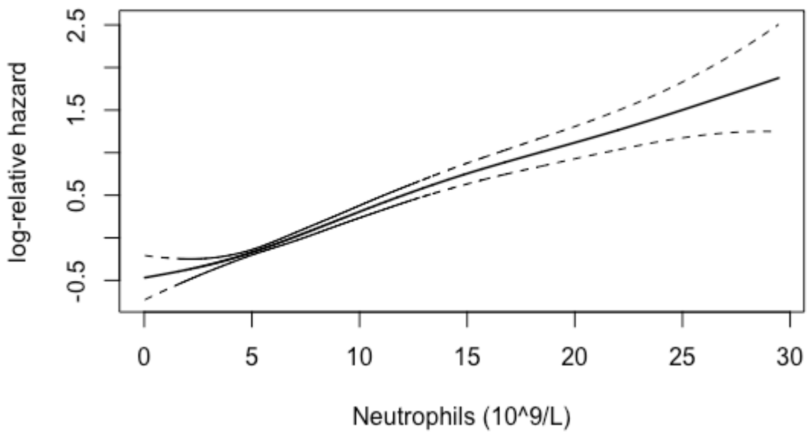


#LY Waves 120

#Call:
### coxph(formula = Surv(ddInwFUp, deadTRUE, type = "right") ~ pspline(linfociti.x,
### df = 3, caic = TRUE), data = cutpoints99_Wave1_2_0_CSV)
#
#coef se(coef) se2 Chisq DF p
#pspline(linfociti.x, df = -0.4878 0.0276 0.0274 311.7381 1.00 <2e-16
### pspline(linfociti.x, df = 311.7628 2.09 <2e-16
#
### Iterations: 5 outer, 16 Newton-Raphson
### Theta= 0.945
### Degrees of freedom for terms= 3.1
### Likelihood ratio test=646 on 3.09 df, p=<2e-16
### n= 11052, number of events= 1597

termplot(LyWav120Cox, term=1, se=TRUE, col.term=1, col.se=1)

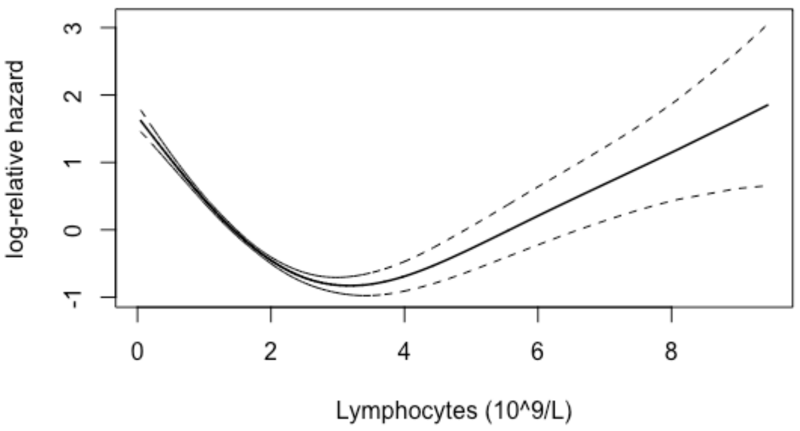


```
LyWav120cutpts <-findcutpoints(cox_pspline_fit=LyWav120Cox, data.frame(
cutpoints99_Wave1_2_0_CSV), nquantile = 100, exclude = 0.05, eps = 0.01, shape = "U")

LyWav120cutpts$optimal
#Cutpoint_L Cutpoint_R
# 1.04      6.42


# Mo Waves 120

#Call:
#coxph(formula = Surv(ddInwFUp, deadTRUE, type = "right") ~ pspline(monociti.x,
#                                        df = 3, caic = TRUE), data = cutpoints99_Wave1_2_0_CSV)
#
#coef se(coef)     se2    Chisq   DF      p
#pspline(monociti.x, df =    0.1270   0.0688   0.0687   3.4121 1.00  0.065
#      pspline(monociti.x, df =                       145.3521 2.06 <2e-16
#
#            Iterations: 6 outer, 19 Newton-Raphson
#            Theta= 0.976
#            Degrees of freedom for terms= 3.1
#            Likelihood ratio test=163  on 3.06 df, p=<2e-16
#            n= 11052, number of events= 1597

termplot(MoWav120Cox, term=1, se=TRUE, col.term=1, col.se=1)
```

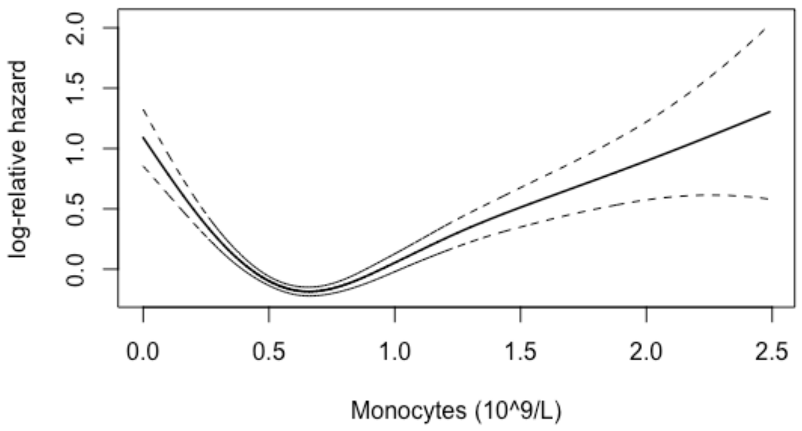


```
MoWav120cutpts <-findcutpoints(cox_pspline_fit=MoWav120Cox, data.frame(
cutpoints99_Wave1_2_0_CSV), nquantile = 100, exclude = 0.05, eps = 0.01, shape = "U")

MoWav120cutpts$optimal
#Cutpoint_L Cutpoint_R
#0.25      1.32


# Eos Waves120

#Call:
#coxph(formula = Surv(ddInwFUp, deadTRUE, type = "right") ~ pspline(eosinofili.x,
#                                                df = 3, caic = TRUE), data = cutpoints99_Wave1_2_0_CSV)
#
#coef se(coef)    se2   Chisq   DF      p
#pspline(eosinofili.x, df   -0.908    0.207   0.196  19.151 1.00 1.2e-05
#      pspline(eosinofili.x, df                     203.530 2.08 < 2e-16
#
#            Iterations: 5 outer, 15 Newton-Raphson
#            Theta= 0.929
#            Degrees of freedom for terms= 3.1
#            Likelihood ratio test=266  on 3.08 df, p=<2e-16
#            n= 11052, number of events= 1597

termplot(EoWav120Cox, term=1, se=TRUE, col.term=1, col.se=1)
```

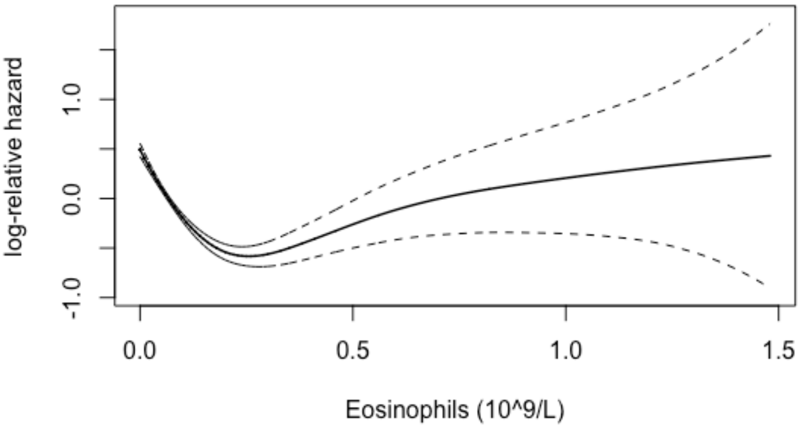


```
EoWav120cutpts <-findcutpoints(cox_pspline_fit=EoWav120Cox, data.frame(
cutpoints99_Wave1_2_0_CSV), nquantile = 100, exclude = 0.05, eps = 0.01, shape = "U")

EoWav120cutpts$optimal
#Cutpoint_L Cutpoint_R
#0.01      1.41


# BASOFILI Waves120


#Call:
#coxph(formula = Surv(ddInwFUp, deadTRUE, type = "right") ~ pspline(basofili.x,
#                                        df = 3, caic = TRUE), data = cutpoints99_Wave1_2_0_CSV)
#
#coef se(coef)    se2   Chisq   DF      p
#pspline(basofili.x, df =   -7.261    1.005   0.984  52.169 1.00 5.1e-13
#       pspline(basofili.x, df =                     187.905 2.04 < 2e-16
#
#             Iterations: 6 outer, 17 Newton-Raphson
#             Theta= 0.955
#             Degrees of freedom for terms= 3
#             Likelihood ratio test=246  on 3.04 df, p=<2e-16
#             n= 11052, number of events= 1597

termplot(BaWav120Cox, term=1, se=TRUE, col.term=1, col.se=1)
```

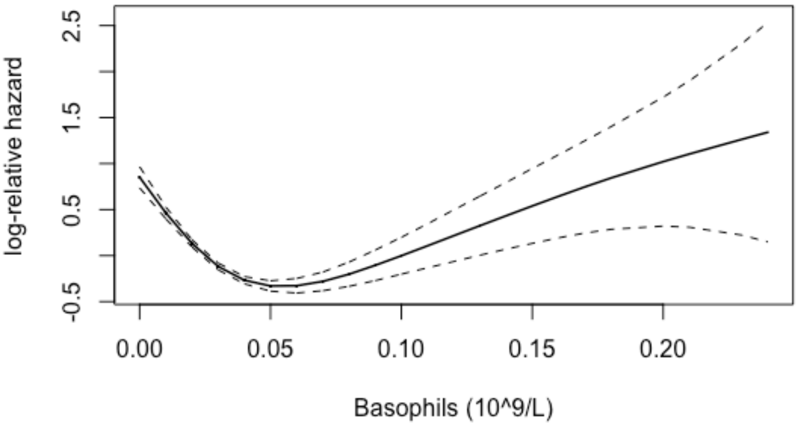


```
BaWav120cutpts <-findcutpoints(cox_pspline_fit=BaWav120Cox, data.frame(
cutpoints99_Wave1_2_0_CSV), nquantile = 100, exclude = 0.05, eps = 0.01, shape = "U")

BaWav120cutpts$optimal
#Cutpoint_L Cutpoint_R
#0.0177     0.1192


# PDW waves120

#Call:
#  coxph(formula = Surv(ddInwFUp, deadTRUE, type = "right") ~ pspline(PDW.x,
#                                              df = 3, caic = TRUE), data = cutpoints99_Wave1_2_0_CSV)
#
#coef se(coef)      se2    Chisq   DF       p
#pspline(PDW.x, df = 3, ca  0.05234  0.00936  0.00936 31.24807 1.00 2.3e-08
#       pspline(PDW.x, df = 3, ca                        60.28198 2.04 8.8e-14
#
#             Iterations: 6 outer, 17 Newton-Raphson
#             Theta= 0.949
#             Degrees of freedom for terms= 3
#             Likelihood ratio test=88.8  on 3.04 df, p=<2e-16
#             n= 11052, number of events= 1597

termplot(PDWWav120Cox, term=1, se=TRUE, col.term=1, col.se=1)
```

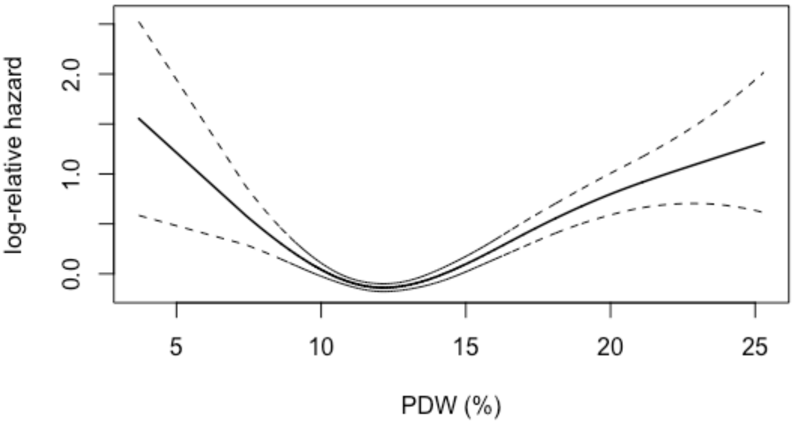


PDWWav120cutpts <-findcutpoints(cox_pspline_fit=PDWWav120Cox, data.frame(
cutpoints99_Wave1_2_0_CSV), nquantile = 100, exclude = 0.05, eps = 0.01, shape = "U")

PDWWav120cutpts$optimal
#Cutpoint_L Cutpoint_R
#8.9 16.0

### PIASTRINE wave 120

```
#Call:
#coxph(formula = Surv(ddInwFUp, deadTRUE, type = "right") ~ pspline(PIASTRINE.x,
#                                  df = 3, caic = TRUE), data = cutpoints99_Wave1_2_0_CSV)
#
#coef  se(coef)      se2     Chisq  DF     p
#pspline(PIASTRINE.x, df = -7.15e-04  2.49e-04  2.49e-04  8.27e+00 1.0  0.004
#     pspline(PIASTRINE.x, df =                         2.74e+02 2.1 <2e-16
#
#           Iterations: 5 outer, 18 Newton-Raphson
#           Theta= 0.944
#           Degrees of freedom for terms= 3.1
#           Likelihood ratio test=301  on 3.1 df, p=<2e-16
#           n= 11052, number of events= 1597
```

termplot(PLTWav120Cox, term=1, se=TRUE, col.term=1, col.se=1)

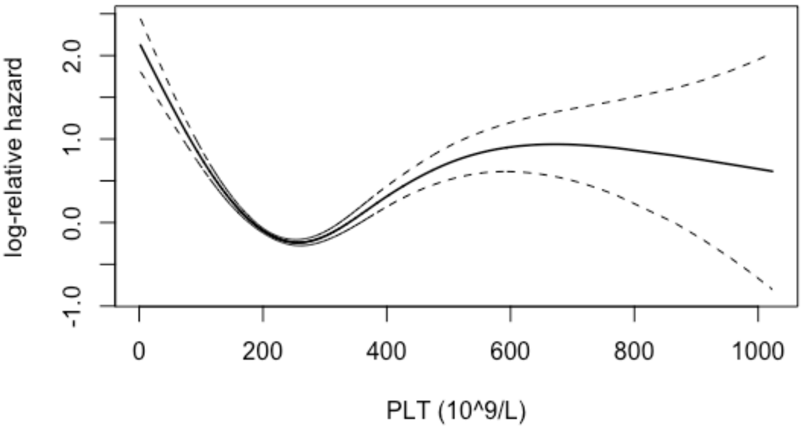


PLTWav120cutpts <-findcutpoints(cox_pspline_fit=PLTWav120Cox, data.frame(
cutpoints99_Wave1_2_0_CSV), nquantile = 100, exclude = 0.05, eps = 0.01, shape = "U")

PLTWav120cutpts$optimal
#Cutpoint_L Cutpoint_R
#131 426

### PCT wave 120

#Call:
#coxph(formula = Surv(ddInwFUp, deadTRUE, type = "right") ~ pspline(PCT.x,
### df = 3, caic = TRUE), data = cutpoints99_Wave1_2_0_CSV)
#
#coef se(coef) se2 Chisq DF p
#pspline(PCT.x, df = 3, ca -0.318 0.248 0.248 1.634 1.00 0.2
### pspline(PCT.x, df = 3, ca 296.672 2.09 <2e-16
#
### Iterations: 5 outer, 18 Newton-Raphson
### Theta= 0.945
### Degrees of freedom for terms= 3.1
### Likelihood ratio test=319 on 3.09 df, p=<2e-16
### n= 11052, number of events= 1597

termplot(PCTWav120Cox, term=1, se=TRUE, col.term=1, col.se=1)

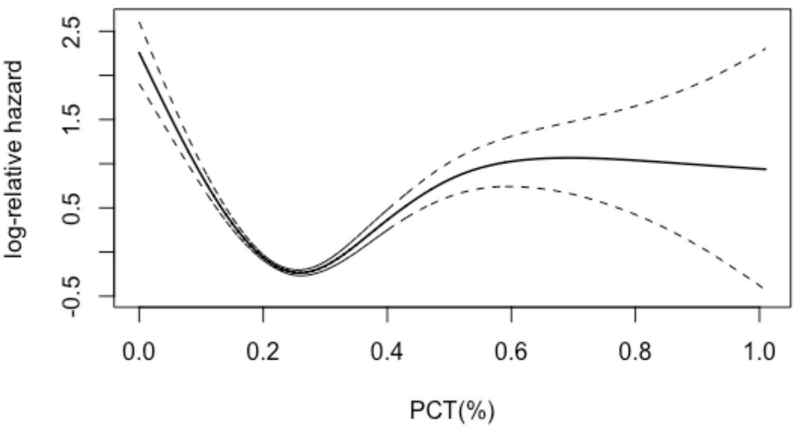


PCTWav120cutpts <-findcutpoints(cox_pspline_fit=PCTWav120Cox, data.frame(
cutpoints99_Wave1_2_0_CSV), nquantile = 100, exclude = 0.05, eps = 0.01, shape = "U")

PCTWav120cutpts$optimal
#Cutpoint_L Cutpoint_R
#0.1476 0.3972

### Eta.x Waves 120

#Call:
### coxph(formula = Surv(ddInwFUp, deadTRUE, type = "right") ~ pspline(Eta.x,
### df = 3, caic = TRUE), data = cutpoints99_Wave1_2_0_CSV)
#
#coef se(coef) se2 Chisq DF p
#pspline(Eta.x, df = 3, ca 7.18e-02 2.17e-03 2.17e-03 1.09e+03 1.00 <2e-16
### pspline(Eta.x, df = 3, ca 1.38e+00 2.06 0.51
#
### Iterations: 6 outer, 17 Newton-Raphson
### Theta= 0.971
### Degrees of freedom for terms= 3.1
### Likelihood ratio test=1675 on 3.06 df, p=<2e-16
### n= 11052, number of events= 1597

termplot(EtaWav120Cox, term=1, se=TRUE, col.term=1, col.se=1)

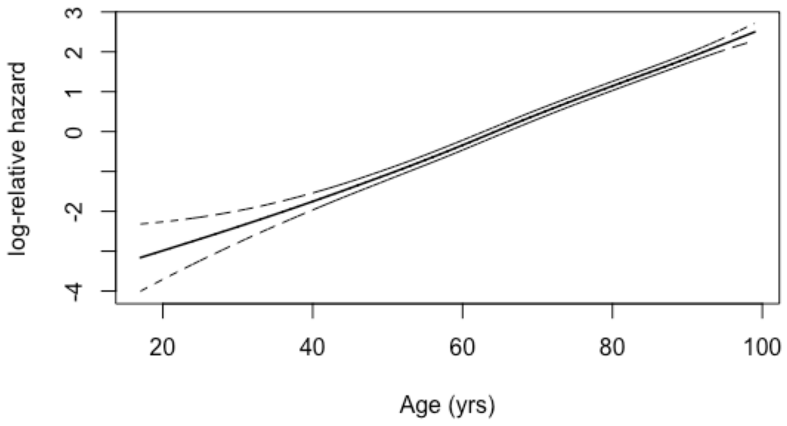
log-relative hazard
3
2
1
0
-2
-4
20
40
60
80
100
Age (yrs)

#### MSRS (maximally selected rank statistic method) techniques for linear distribution variables (Age, Hemoglobin, Neutrophils)

.

### Cox Regression Summary and Table - Age

Cox Table- Eta.x

| Explanatory | Levels | all | HR (Univariable) | HR (Multivariable) |
|---|---|---|---|---|
| Eta.x | Mean (SD) | 64.2 (19.7) | 1.07 (1.07-1.08, p<0.001) | 1.07 (1.07-1.08, p<0.001) |

**Model Metrics:** Number in dataframe = 11052, Number in model = 11052, Missing = 0, Number of events = 1597, Concordance = 0.778 (SE = 0.005), R-squared = 0.140( Max possible = 0.929), Likelihood ratio test = 1672.822 (df = 1, p = 0.000)

Eta.x

| Cut Point | Statistic |
|---|---|
| 73.0 | 35.2 |

#### Cutpoint Plot

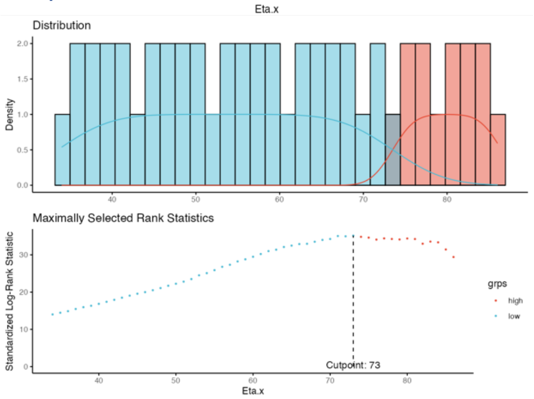


#### Survival Plot with new Cut-off

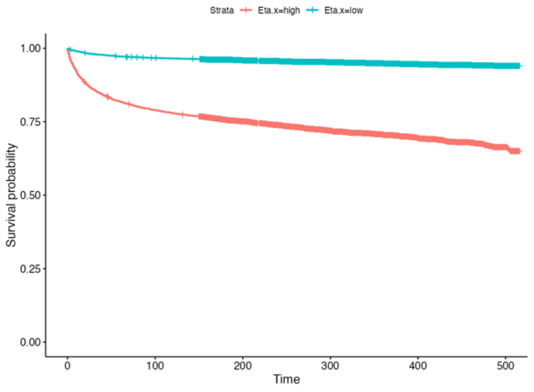

#### Median Survival Summary and Table - Eta.x

```
When Eta.x is high, median survival is NA [NA - NA, 95% CI] months.
When Eta.x is low, median survival is NA [NA - NA, 95% CI] months.
```

Median Survival Table: Levels for Eta.x

| | | | | | | 95% Confidence Interval | |
|---|---|---|---|---|---|---|---|
| Levels | Records | Events | rmean | se_rmean | Median | Lower | Upper |
| Eta.x=high | 4359 | 1263 | 384 | 3.17 | NaN | NaN | NaN |
| Eta.x=low | 6693 | 334 | 494 | 1.20 | NaN | NaN | NaN |

#### 1, 3, 5-yr Survival Summary and Table - Eta.x

```
When Eta.x=high, 12 month survival is 90.78% [89.92%-91.64%, 95% CI].
When Eta.x=high, 36 month survival is 84.86% [83.80%-85.93%, 95% CI].
When Eta.x=high, 60 month survival is 81.83% [80.69%-82.98%, 95% CI].
When Eta.x=low, 12 month survival is 98.89% [98.64%-99.15%, 95% CI].
When Eta.x=low, 36 month survival is 97.82% [97.47%-98.17%, 95% CI].
When Eta.x=low, 60 month survival is 97.21% [96.81%-97.60%, 95% CI].
```

1, 3, 5 year Survival - Eta.x

| | | | | | 95% Confidence Interval | |
|---|---|---|---|---|---|---|
| Levels | time | Number at Risk | Number of Events | Survival | Lower | Upper |
| Eta.x=high | 12 | 3978 | 402 | 90.8 % | 89.9 % | 91.6 % |
| Eta.x=high | 36 | 3701 | 258 | 84.9 % | 83.8 % | 85.9 % |
| Eta.x=high | 60 | 3568 | 132 | 81.8 % | 80.7 % | 83.0 % |
| Eta.x=low | 12 | 6623 | 74 | 98.9 % | 98.6 % | 99.1 % |
| Eta.x=low | 36 | 6545 | 72 | 97.8 % | 97.5 % | 98.2 % |
| Eta.x=low | 60 | 6503 | 41 | 97.2 % | 96.8 % | 97.6 % |

#### Cumulative Hazard - Eta.x

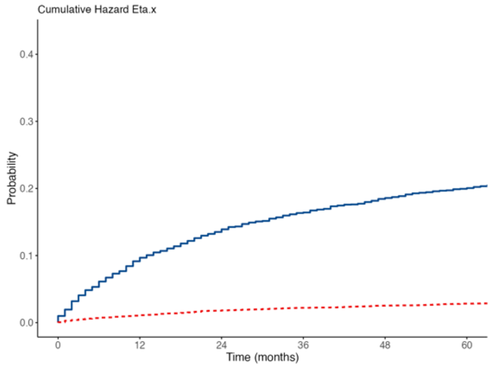

#### Cox Regression Summary and Table - Hemoglobin

Cox Table- EMOGLOBINA.x

| Explanatory | Levels | all | HR (Univariable) | HR (Multivariable) |
|---|---|---|---|---|
| EMOGLOBINA.x | Mean (SD) | 13.5 (2.0) | 0.78 (0.76-0.79, p<0.001) | 0.78 (0.76-0.79, p<0.001) |

**Model Metrics:** Number in dataframe = 11052, Number in model = 11052, Missing = 0, Number of events = 1597, Concordance = 0.665 (SE = 0.007), R-squared = 0.049( Max possible = 0.929), Likelihood ratio test = 556.119 (df = 1, p = 0.000)

EMOGLOBINA.x

| Cut Point | Statistic |
|---|---|
| 11.7 | 25.4 |

##### Cutpoint Plot

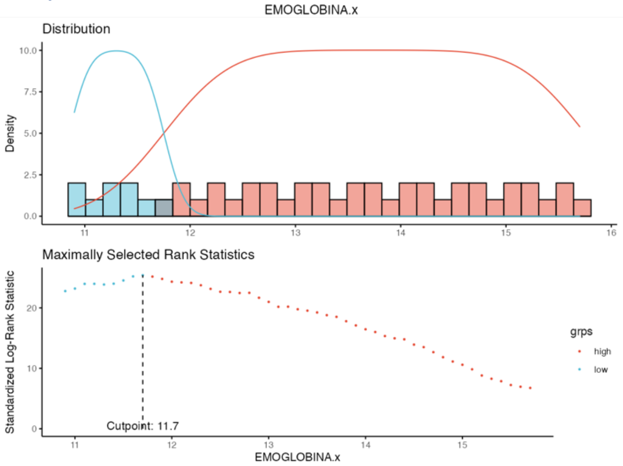


##### Survival Plot with new Cut-off

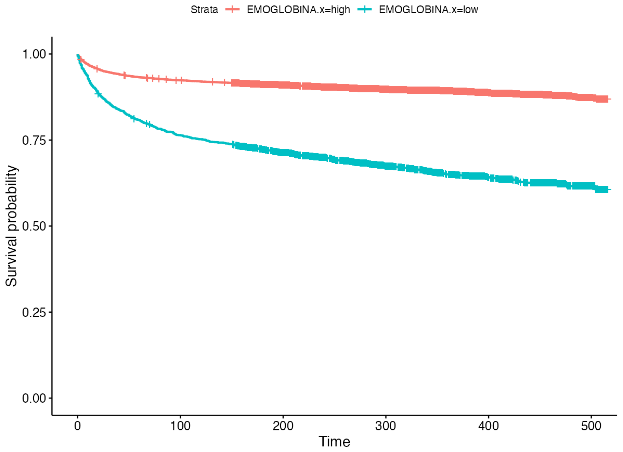

#### Median Survival Summary and Table - EMOGLOBINA.x

```
When EMOGLOBINA.x is high, median survival is NA [NA - NA, 95% CI] months.
When EMOGLOBINA.x is low, median survival is NA [NA - NA, 95% CI] months.
```

Median Survival Table: Levels for EMOGLOBINA.x

| | | | | | | 95% Confidence Interval | |
|---|---|---|---|---|---|---|---|
| **Levels** | **Records** | **Events** | **rmean** | **se_rmean** | **Median** | **Lower** | **Upper** |
| EMOGLOBINA.x=high | 9201 | 977 | 468 | 1.48 | NaN | NaN | NaN |
| EMOGLOBINA.x=low | 1851 | 620 | 365 | 5.00 | NaN | NaN | NaN |

#### 1, 3, 5-yr Survival Summary and Table - EMOGLOBINA.x

```
When EMOGLOBINA.x=high, 12 month survival is 96.5% [96.2%-96.91%, 95% CI].
When EMOGLOBINA.x=high, 36 month survival is 94.4% [93.9%-94.83%, 95% CI].
When EMOGLOBINA.x=high, 60 month survival is 93.2% [92.7%-93.74%, 95% CI].
When EMOGLOBINA.x=low, 12 month survival is 91.5% [90.3%-92.80%, 95% CI].
When EMOGLOBINA.x=low, 36 month survival is 84.5% [82.9%-86.16%, 95% CI].
When EMOGLOBINA.x=low, 60 month survival is 80.8% [79.0%-82.58%, 95% CI].
```

1, 3, 5 year Survival - EMOGLOBINA.x

| | | | | | 95% Confidence Interval | |
|---|---|---|---|---|---|---|
| **Levels** | **time** | **Number at Risk** | **Number of Events** | **Survival** | **Lower** | **Upper** |
| EMOGLOBINA.x=high | 12 | 8894 | 319 | 96.5 % | 96.2 % | 96.9 % |
| EMOGLOBINA.x=high | 36 | 8682 | 200 | 94.4 % | 93.9 % | 94.8 % |
| EMOGLOBINA.x=high | 60 | 8576 | 104 | 93.2 % | 92.7 % | 93.7 % |
| EMOGLOBINA.x=low | 12 | 1707 | 157 | 91.5 % | 90.3 % | 92.8 % |
| EMOGLOBINA.x=low | 36 | 1564 | 130 | 84.5 % | 82.9 % | 86.2 % |
| EMOGLOBINA.x=low | 60 | 1495 | 69 | 80.8 % | 79.0 % | 82.6 % |

#### Cumulative Hazard - EMOGLOBINA.x

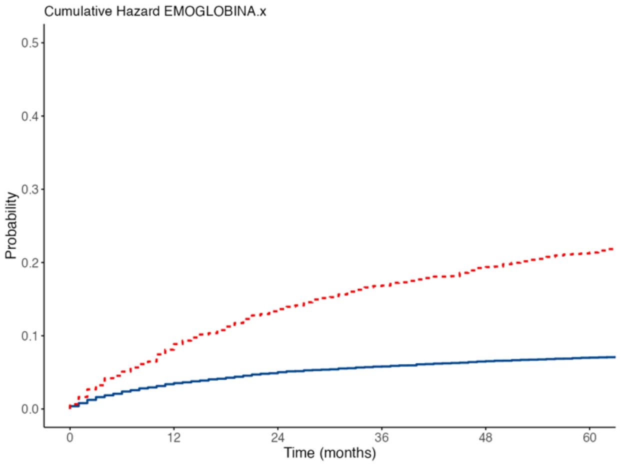

### Cox Regression Summary and Table - MCV

Cox Table- MCV.x

| Explanatory | Levels | all | HR (Univariable) | HR (Multivariable) |
|---|---|---|---|---|
| MCV.x | Mean (SD) | 90.2 (6.6) | 1.05 (1.04-1.06, p<0.001) | 1.05 (1.04-1.06, p<0.001) |

**Model Metrics:** Number in dataframe = 11052, Number in model = 11052, Missing = 0, Number of events = 1597, Concordance = 0.598 (SE = 0.008), R-squared = 0.016( Max possible = 0.929), Likelihood ratio test = 173.426 (df = 1, p = 0.000)

MCV.x

| Cut Point | Statistic |
|---|---|
| 97.1 | 17.4 |

#### Cutpoint Plot

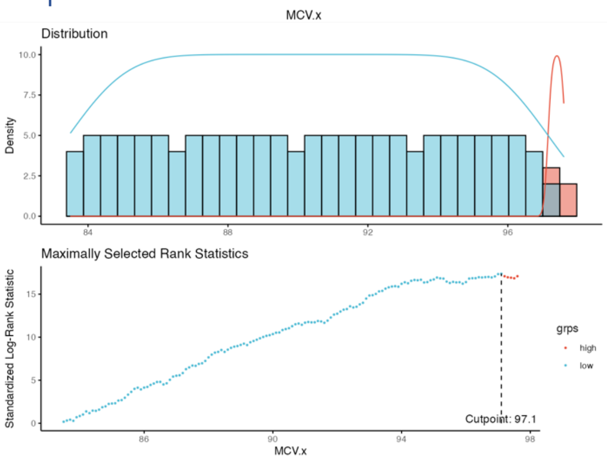


#### Survival Plot with new Cut-off

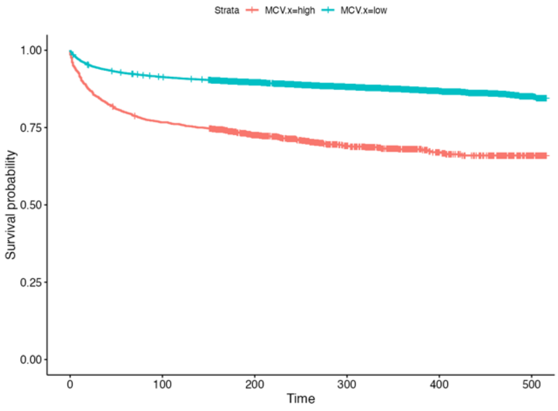

#### Median Survival Summary and Table - MCV.x

```
When MCV.x is high, median survival is NA [NA - NA, 95% CI] months.
When MCV.x is low, median survival is NA [NA - NA, 95% CI] months.
```

Median Survival Table: Levels for MCV.x

| | | | | | | 95% Confidence Interval | |
|---|---|---|---|---|---|---|---|
| Levels | Records | Events | rmean | se_rmean | Median | Lower | Upper |
| MCV.x=high | 1249 | 377 | 373 | 6.16 | NaN | NaN | NaN |
| MCV.x=low | 9803 | 1220 | 460 | 1.52 | NaN | NaN | NaN |

#### 1, 3, 5-yr Survival Summary and Table - MCV.x

```
When MCV.x=high, 12 month survival is 90.2% [88.6%-91.9%, 95% CI].
When MCV.x=high, 36 month survival is 83.5% [81.5%-85.6%, 95% CI].
When MCV.x=high, 60 month survival is 79.9% [77.7%-82.2%, 95% CI].
When MCV.x=low, 12 month survival is 96.4% [96.0%-96.8%, 95% CI].
When MCV.x=low, 36 month survival is 93.9% [93.4%-94.4%, 95% CI].
When MCV.x=low, 60 month survival is 92.6% [92.1%-93.1%, 95% CI].
```

1, 3, 5 year Survival - MCV.x

| | | | | | 95% Confidence Interval | |
|---|---|---|---|---|---|---|
| Levels | time | Number at Risk | Number of Events | Survival | Lower | Upper |
| MCV.x=high | 12 | 1138 | 122 | 90.2 % | 88.6 % | 91.9 % |
| MCV.x=high | 36 | 1045 | 84 | 83.5 % | 81.5 % | 85.6 % |
| MCV.x=high | 60 | 998 | 45 | 79.9 % | 77.7 % | 82.2 % |
| MCV.x=low | 12 | 9463 | 354 | 96.4 % | 96.0 % | 96.8 % |
| MCV.x=low | 36 | 9201 | 246 | 93.9 % | 93.4 % | 94.4 % |
| MCV.x=low | 60 | 9073 | 128 | 92.6 % | 92.1 % | 93.1 % |

#### Cumulative Hazard - MCV.x

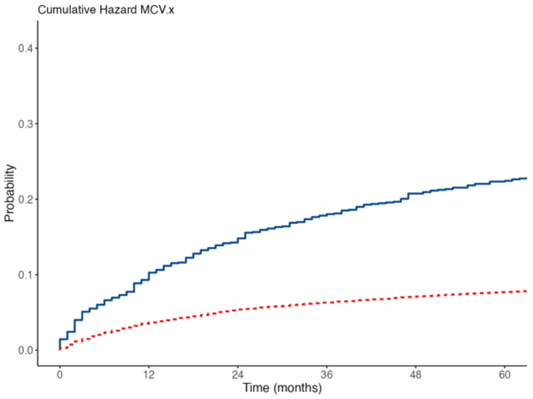

### Cox Regression Summary and Table - RDW

Cox Table- RDW.x

| Explanatory | Levels | all | HR (Univariable) | HR (Multivariable) |
|---|---|---|---|---|
| RDW.x | Mean (SD) | 13.7 (1.8) | 1.28 (1.26-1.29, p<0.001) | 1.28 (1.26-1.29, p<0.001) |

**Model Metrics:** Number in dataframe = 11052, Number in model = 11052, Missing = 0, Number of events = 1597, Concordance = 0.739 (SE = 0.006), R-squared = 0.059( Max possible = 0.929), Likelihood ratio test = 673.081 (df = 1, p = 0.000)

RDW.x

| Cut Point | Statistic |
|---|---|
| 13.8 | 32.2 |

#### Cutpoint Plot

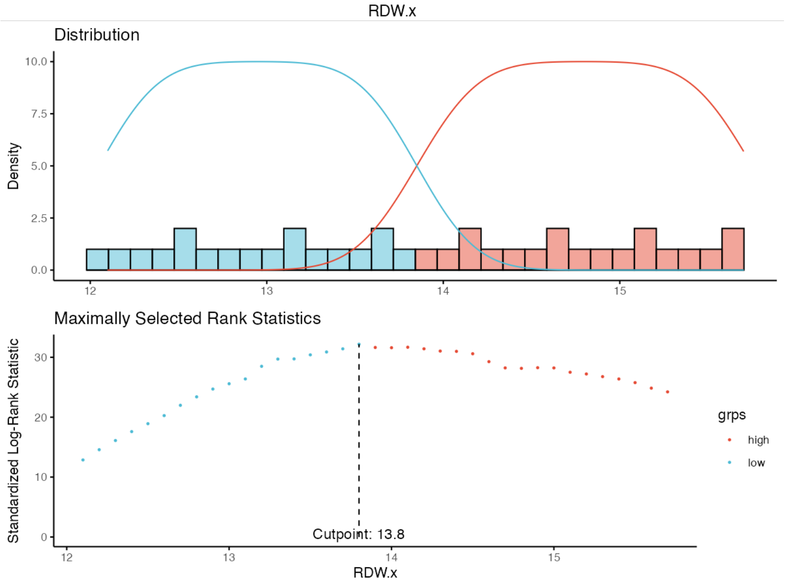


#### Survival Plot with new Cut-off

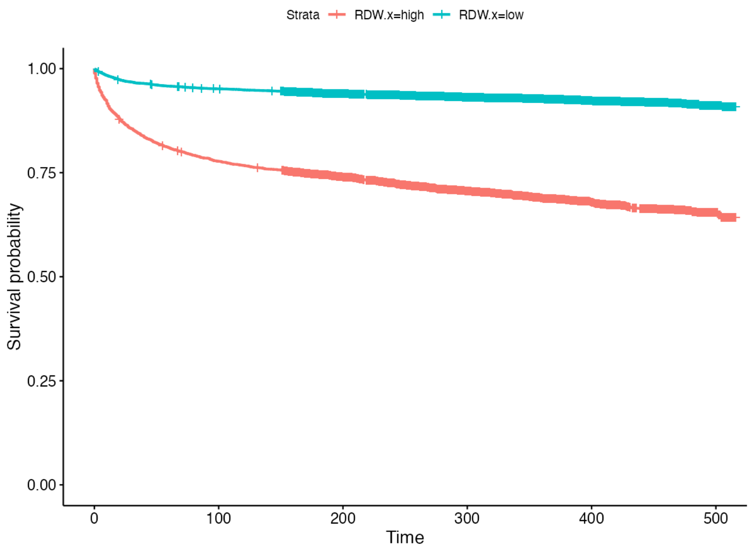


#### Median Survival Summary and Table - RDW.x

```
When RDW.x is high, median survival is NA [NA - NA, 95% CI] months.
When RDW.x is low, median survival is NA [NA - NA, 95% CI] months.
```

Median Survival Table: Levels for RDW.x

| | | | | | | 95% Confidence Interval | |
|---|---|---|---|---|---|---|---|
| **Levels** | **Records** | **Events** | **rmean** | **se_rmean** | **Median** | **Lower** | **Upper** |
| RDW.x=high | 3467 | 1051 | 378 | 3.60 | NaN | NaN | NaN |
| RDW.x=low | 7585 | 546 | 484 | 1.34 | NaN | NaN | NaN |

#### 1, 3, 5-yr Survival Summary and Table - RDW.x

```
When RDW.x=high, 12 month survival is 90.40% [89.42%-91.38%, 95% CI].
When RDW.x=high, 36 month survival is 84.39% [83.20%-85.61%, 95% CI].
When RDW.x=high, 60 month survival is 80.96% [79.66%-82.28%, 95% CI].
When RDW.x=low, 12 month survival is 98.11% [97.81%-98.42%, 95% CI].
When RDW.x=low, 36 month survival is 96.51% [96.09%-96.92%, 95% CI].
When RDW.x=low, 60 month survival is 95.79% [95.34%-96.25%, 95% CI].
```

1, 3, 5 year Survival - RDW.x

| | | | | | 95% Confidence Interval | |
|---|---|---|---|---|---|---|
| **Levels** | **time** | **Number at Risk** | **Number of Events** | **Survival** | **Lower** | **Upper** |
| RDW.x=high | 12 | 3154 | 333 | 90.4 % | 89.4 % | 91.4 % |
| RDW.x=high | 36 | 2927 | 208 | 84.4 % | 83.2 % | 85.6 % |
| RDW.x=high | 60 | 2809 | 119 | 81.0 % | 79.7 % | 82.3 % |
| RDW.x=low | 12 | 7447 | 143 | 98.1 % | 97.8 % | 98.4 % |
| RDW.x=low | 36 | 7319 | 122 | 96.5 % | 96.1 % | 96.9 % |
| RDW.x=low | 60 | 7262 | 54 | 95.8 % | 95.3 % | 96.2 % |

#### Cumulative Hazard - RDW.x

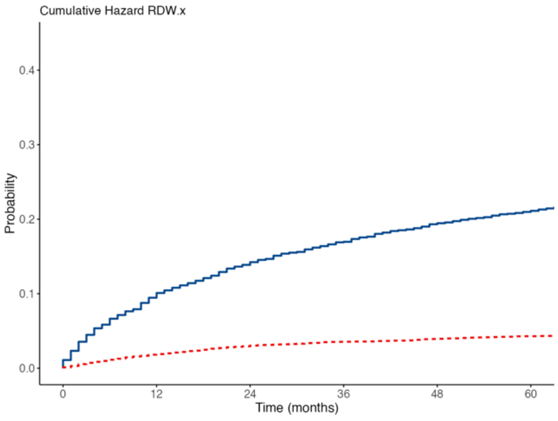


### Cox Regression Summary and Table - Neutrophils

```
When neutrofili.x increases 1 unit, the hazard increases 1.09 (1.08-1.10, p
```

Cox Table- neutrofili.x

| Explanatory | Levels | all | HR (Univariable) | HR (Multivariable) |
|---|---|---|---|---|
| neutrofili.x | Mean (SD) | 6.7 (3.7) | 1.09 (1.08-1.10, p<0.001) | 1.09 (1.08-1.10, p<0.001) |

**Model Metrics:** Number in dataframe = 11052, Number in model = 11052, Missing = 0, Number of events = 1597, Concordance = 0.598 (SE = 0.008), R-squared = 0.020( Max possible = 0.929), Likelihood ratio test = 224.707 (df = 1, p = 0.000)

neutrofili.x

| Cut Point | Statistic |
|---|---|
| 9.65 | 13.4 |

#### Cutpoint Plot

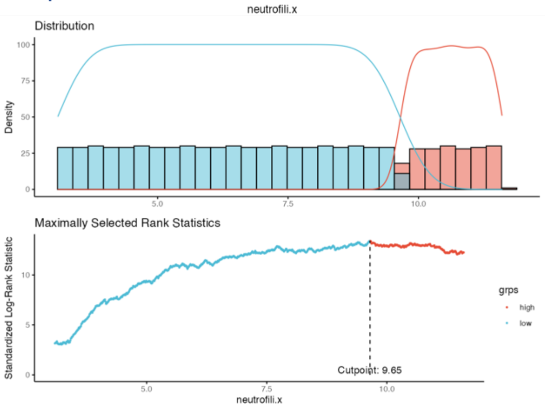


#### Survival Plot with new Cut-off

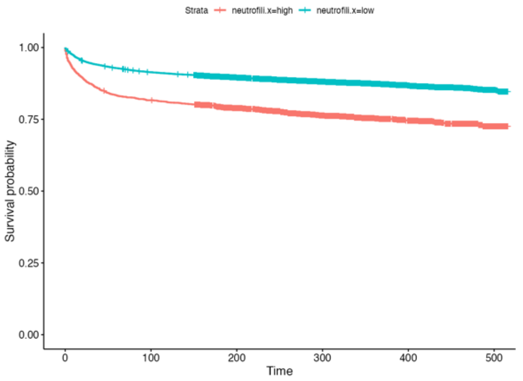


#### Median Survival Summary and Table - neutrofili.x

```
When neutrofili.x is high, median survival is NA [NA - NA, 95% CI] months.
When neutrofili.x is low, median survival is NA [NA - NA, 95% CI] months.
```

Median Survival Table: Levels for neutrofili.x

| | | | | | | 95% Confidence Interval | |
|---|---|---|---|---|---|---|---|
| Levels | Records | Events | rmean | se_rmean | Median | Lower | Upper |
| neutrofili.x=high | 1934 | 462 | 405 | 4.56 | NaN | NaN | NaN |
| neutrofili.x=low | 9118 | 1135 | 460 | 1.57 | NaN | NaN | NaN |

#### 1, 3, 5-yr Survival Summary and Table - neutrofili.x

```
When neutrofili.x=high, 12 month survival is 91.5% [90.2%-92.72%, 95% CI].
When neutrofili.x=high, 36 month survival is 86.1% [84.6%-87.70%, 95% CI].
When neutrofili.x=high, 60 month survival is 83.2% [81.6%-84.93%, 95% CI].
When neutrofili.x=low, 12 month survival is 96.6% [96.2%-96.96%, 95% CI].
When neutrofili.x=low, 36 month survival is 94.1% [93.6%-94.58%, 95% CI].
When neutrofili.x=low, 60 month survival is 92.8% [92.3%-93.35%, 95% CI].
```

1, 3, 5 year Survival - neutrofili.x

| | | | | | 95% Confidence Interval | |
|---|---|---|---|---|---|---|
| Levels | time | Number at Risk | Number of Events | Survival | Lower | Upper |
| neutrofili.x=high | 12 | 1775 | 165 | 91.5 % | 90.2 % | 92.7 % |
| neutrofili.x=high | 36 | 1667 | 103 | 86.1 % | 84.6 % | 87.7 % |

1, 3, 5 year Survival - neutrofili.x

| | | | | | 95% Confidence Interval | |
|---|---|---|---|---|---|---|
| **Levels** | **time** | **Number at Risk** | **Number of Events** | **Survival** | **Lower** | **Upper** |
| neutrofili.x=high | 60 | 1612 | 56 | 83.2 % | 81.6 % | 84.9 % |
| neutrofili.x=low | 12 | 8826 | 311 | 96.6 % | 96.2 % | 97.0 % |
| neutrofili.x=low | 36 | 8579 | 227 | 94.1 % | 93.6 % | 94.6 % |
| neutrofili.x=low | 60 | 8459 | 117 | 92.8 % | 92.3 % | 93.3 % |

#### Cumulative Hazard - neutrofili.x

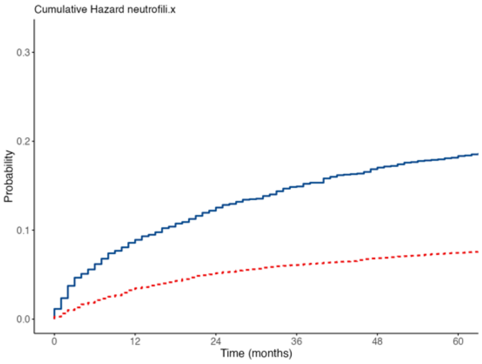
